# Integrated Analysis of Preterm Birth and Socioeconomic Status with Neonatal Brain Structure

**DOI:** 10.1101/2022.12.19.22283518

**Authors:** Katie Mckinnon, Paola Galdi, Manuel Blesa-Cábez, Gemma Sullivan, Kadi Vaher, Amy Corrigan, Jill Hall, Lorena Jiménez-Sánchez, Michael Thrippleton, Mark E Bastin, Alan J. Quigley, Evdoxia Valavani, Athanasios Tsanas, Hilary Richardson, James P. Boardman

**Author notes:** **Corresponding author** Professor James Boardman, FRCPCH, PhD, Medical Research Council Centre for Reproductive Health at the University of Edinburgh, The Queen’s Medical Research Institute, Edinburgh BioQuarter, 47 Little France Crescent, Edinburgh, EH16 4TJ, United Kingdom.

## Abstract

**Importance:** Preterm birth and socioeconomic status (SES) are associated with brain structure in childhood, but the relative contributions of each during the neonatal period are unknown.

**Objective:** To investigate associations of gestational age (GA) and SES with neonatal brain morphology, by testing 3 hypotheses: GA and SES are associated with brain morphology; associations between SES and brain morphology vary across the GA range, and; associations between SES and brain structure/morphology depend on how SES is operationalized.

**Design:** Cohort study, recruited 2016-2021.

**Setting:** Single center, UK.

**Participants:** 170 preterm infants and 91 term infants with median (range) birth GA 30^+0^ (22^+1^-32^+6^) and 39^+4^ (36^+3^-42^+1^) weeks, respectively. Exclusion criteria: major congenital malformation, chromosomal abnormality, congenital infection, cystic periventricular leukomalacia, hemorrhagic parenchymal infarction, post-hemorrhagic ventricular dilatation.

**Exposures:** Using linear ridge regression models, we investigated associations of GA and SES, operationalized at the neighborhood-level (Scottish Index of Multiple Deprivation), family-level (parental education and occupation) and subjectively (WHO Quality of Life), with regional brain volumes and cortical morphology.

**Main outcomes/measures:** Brain volume (85 parcels) and 5 whole-brain cortical morphology measures (gyrification index, thickness, sulcal depth, curvature, surface area) at term-equivalent age.

**Results:** In fully adjusted models, GA associated with a higher proportion of brain volumes (22/85 [26%], β range |-0.13| to |0.22|) than neighborhood SES (1/85 [1%], β=0.17). GA associated with cortical surface area (β=0.10 [95% confidence interval (CI) 0.02-0.18]) and gyrification index (β=0.16 [95% CI 0.07-0.25]); neighborhood SES did not. Family-level SES associated with the volumes of more parcels than neighborhood SES, but it did not have as extensive associations with brain structure as GA. There were interactions between GA and both family- and subjective-level SES measures on brain structure.

**Conclusions/relevance:** In a UK cohort, GA and SES impact neonatal brain morphology, but low GA has more widely distributed effects on neonatal brain structure than neighborhood-level, family-level and subjective measures of SES. Further work is warranted to elucidate the mechanisms embedding GA and level-specific SES in early brain development.

**Three key points:** *Question:* What are the impacts of preterm birth and socioeconomic status (SES), operationalized at neighborhood, family, and subjective levels, on neonatal brain structure?

*Finding:* After mutual adjustment, both low gestational age (GA) and SES associate with brain structure. The nature of SES-brain structure associations varies depending how SES is operationalized; there are interactions between GA and measures of family- and subjective-level SES on brain structure.

*Meaning:* Low GA, and to a lesser extent SES, are associated with neonatal brain structure. Further work is required to elucidate the mechanisms that embed preterm birth and level-specific SES in the developing brain.

## Introduction

Preterm birth, defined as birth <37 weeks of gestation, affects 11% of births globally and is a leading cause of atypical brain development underpinning long-term motor, cognitive, and behavioral problems^1,2^. While there is variability in developmental trajectories and outcomes of people born preterm, there is increased likelihood of cerebral palsy, cognitive impairment, lower educational attainment, visual/hearing impairment, attention deficit/hyperactivity disorder, autism spectrum disorder, and mental health diagnoses across the lifecourse^2–4^.

Preterm birth is associated with structural brain changes apparent by term-equivalent age including global and regional tissue volume reduction, altered cortical configuration, and enlargement of cerebrospinal fluid (CSF) spaces, although tissue loss is not inevitable^5–8^. Changes in neonatal morphology are associated with later functional impairment, highlighting the importance of elucidating factors contributing to early brain development^9–12^.

In the general population, brain structure, cognition, educational attainment, and adult income are patterned by socioeconomic status (SES) in childhood^13^. Children living in poverty are more likely to experience difficulties with memory, language, self-regulation and socioemotional processing, and are more likely to receive behavioral and mental health diagnoses^14–21^. Indeed, mediation analyses suggest a causal pathway between SES and cognitive development in childhood via changes in brain anatomy and function^13,22–25^. SES is inherently a multifaceted construct and can be operationalized using neighborhood-, family-, or subjective-level measures^14,21^. These capture different phenomena and the extent to which they correlate with one another depends on setting and population. Understanding the relative contributions of socioeconomic disadvantage, operationalized in different ways, and low GA to neonatal brain development is important for designing rational therapies and support strategies for children born preterm.

To investigate relationships between GA and SES with neonatal brain development, we combined high-resolution brain magnetic resonance imaging (MRI) from infants born across the GA range with neighborhood-, family-, and subjective-level SES measures. We investigated contributions of GA and SES to brain structure by testing hypotheses that: GA and SES are associated with neonatal brain structure in mutually adjusted models; associations between SES and brain structure vary across the GA range; and associations between SES and brain morphology depend how SES is operationalized.

## Methods

### Study participants

Participants were preterm (birth <33 weeks’ gestation [n=170]) and term control infants (n=91), recruited to a longitudinal study investigating the effect of preterm birth on brain development and outcomes^26^ (participant flow diagram – eFigure 1). Recruitment was at the Royal Infirmary of Edinburgh, UK, between 2016-2021. Ethical approval was obtained from the UK National Research Ethics Service and parents provided written consent (South East Scotland Research Ethic Committee 16/SS/0154).

Exclusion criteria were major congenital malformation, chromosomal abnormality, congenital infection, cystic periventricular leukomalacia, hemorrhagic parenchymal infarction, and post-hemorrhagic ventricular dilatation.

### Demographic variables

We assigned preterm infants into four categories with similar numbers of participants based on GA: 22^+0^-26^+6^, 27^+0^-28^+6^, 29^+0^-30^+6^, and 31^+0^-32^+6^ weeks’ gestation.

SES was measured in three ways: neighborhood-level, defined by the Scottish Index of Multiple Deprivation 2016 (SIMD) derived from the family’s postal code at birth^27^; family-level, defined as parental education (highest educational qualification and age leaving education) and parental occupation (current/most recent job); and subjective SES, defined using the environment domain of the World Health Organization Quality of Life (WHO QoL) assessment^28^. SIMD rank was chosen as the primary measure of SES because it correlates with child development^29^ and is a tractable tool for policy makers^27^. Other SES measures were investigated in exploratory analyses, as specified in our preregistered statistical plan^30^: maternal and paternal education, maternal and paternal occupation, and subjective SES (eMethods).

### Selection of covariates

Covariate selection was based on associations with brain structure in prior research: birth weight z-score^31,32^, birth head circumference z-score^33^, age at MRI, infant sex^34^, smoking in pregnancy^35,36^, and any breast milk at discharge^37,38^ (see eMethods for definitions).

### MRI Data Acquisition

MRI scans were performed at term-corrected gestation according to a published protocol^26^. In summary, a Siemens MAGNETOM Prisma 3T MRI clinical scanner (Siemens Healthcare, Erlangen, Germany) and 16-channel phased-array pediatric head receive coil were used to acquire: a 3D T1-weighted magnetization-prepared rapid acquisition with gradient echo structural volume scan (voxel size=1mm isotropic); and a 3D T2-weighted (T2w) sampling scheme with application-optimized contrasts using flip angle evolution structural scan (voxel size=1mm isotropic).

Infants were fed, wrapped, and slept naturally. Flexible earplugs and neonatal earmuffs (MiniMuffs, Natus) were used for acoustic protection. Infants were monitored throughout, and scans were supervised by a doctor or nurse trained in neonatal resuscitation.

### MRI Data Analysis

Structural images were reported by a radiologist with experience in neonatal MRI (A.J.Q.). The developing Human Connectome Project (dHCP) minimal processing pipeline for neonatal data^39^ was used to preprocess T2w images, allowing surface reconstruction from tissue segmentation. We obtained bias field corrected T2w, brain masks, tissue segmentation, label parcellation and surface reconstruction. We then calculated tissue volumes, gyrification index, cortical thickness, sulcal depth, cortical curvature and cortical surface area^39^.

### Selection of image features

We included 85 individual regional brain parcels (excluding the two background parcels) and 5 whole-cortex measures (gyrification index, thickness, sulcal depth, curvature, surface area), as defined by the dHCP^39^.

### Statistical analyses

Statistical analyses were preregistered^30^ and conducted in R (version 4.2.1)^40^.

We compared demographic data and SES measures across preterm and term groups. We compared categorical variables using Chi-squared tests (significance threshold *p*<.05), and Cramer’s V for estimating effect sizes. For continuous variables, we used Mann-Whitney U-tests and Cohen’s D. We assessed statistical relationships between SES measures using Spearman’s correlation, with strength of correlation classified as very weak (r=0-0.19), weak (r=0.2-0.39), moderate (r=0.40-0.59), strong (r=0.6-0.79), or very strong (r=0.8-1).

Because the sample included twins (25 pairs) and siblings (5 groups), which violates the assumption of non-independence among data-points, we repeated analyses after random removal of one sibling or twin per family.

To investigate associations between SES and GA with regional brain volumes and cortical measures, we developed regression models. Model 1 was a baseline linear regression model, including GA at birth, SIMD rank, gestational age at MRI, and a product interaction term (GA at birth x SIMD, if significant). Model 2 was a ridge regression (L2 penalization linear regression) model^41^, including all covariates; GA at birth, SES, gestation at MRI, the product interaction term (if significant), birth weight z-score, birth head circumference z-score, sex, smoking in pregnancy, and breast milk at discharge. Ridge regression aims to partially mitigate potential multicollinearity among predictors. We ran 85 regression models testing the relationship between each SES measure and GA and each regional brain volume, and 5 additional regression models testing the relationship between each SES measure and GA and each whole-brain cortical measure. Results are reported as standardized beta values with 95% confidence intervals (CI). To correct for multiple comparisons for each measure, we used Benjamini-Hochberg correction^42^, with *p*-value threshold *p*<.05. We compared the frequency of brain volume associations with GA and SES variables using McNemar’s tests.

As described in the statistical analysis plan^30^, SIMD was the primary SES measure in analyses, and maternal and paternal education, maternal and paternal occupation, and subjective SES were investigated in exploratory analyses. For these, the same statistical thresholds used in the primary analyses were applied.

## Results

### Participant characteristics

Participants were 170 preterm and 91 term born infants; their demographic characteristics are shown in Table 1. There was no difference in sex distribution across groups. Ethnicity did not differ between groups and is representative of Edinburgh^43^. All SES measures, smoking prevalence and multiple pregnancy differed between groups.

**Table 1.**
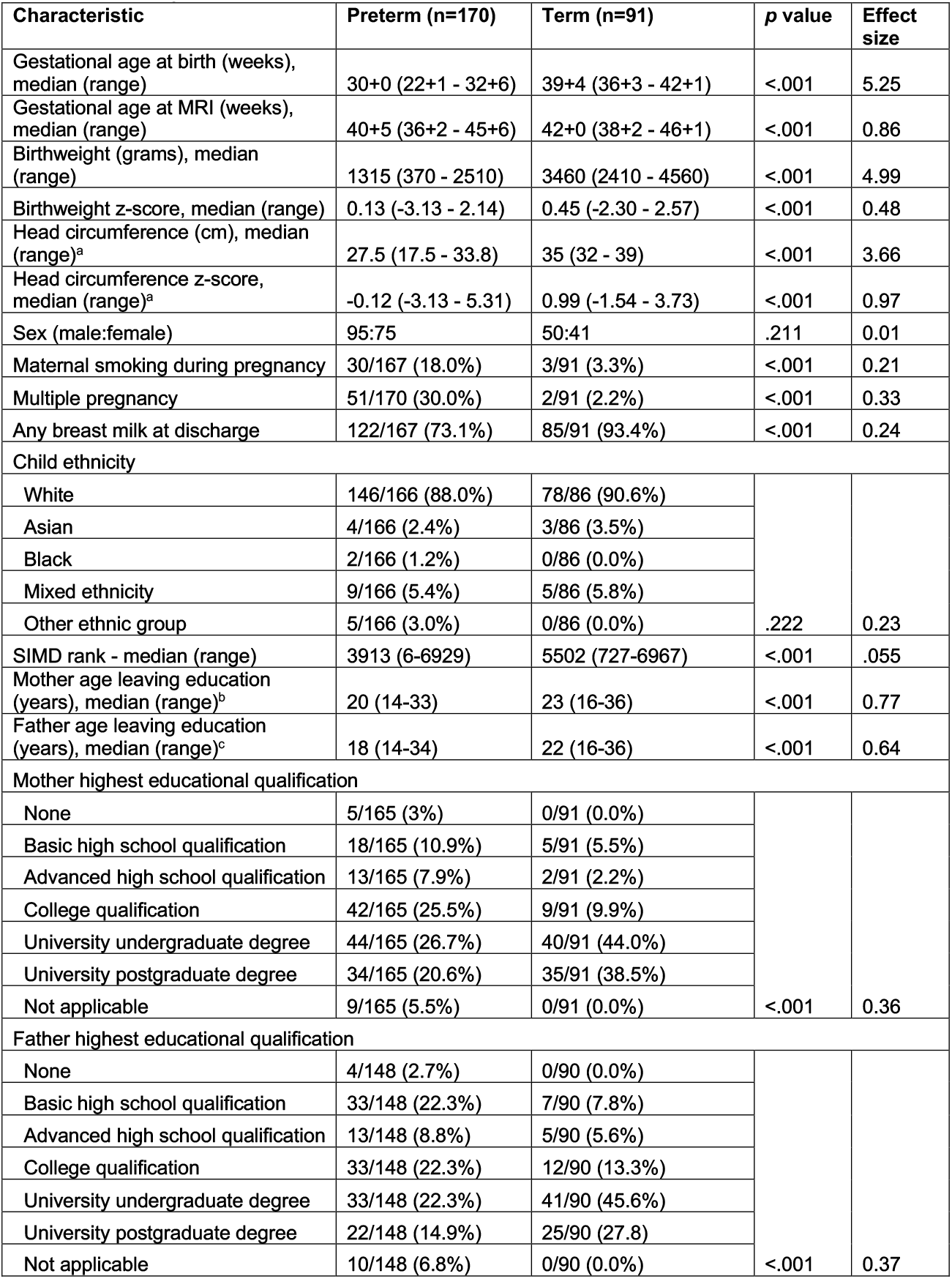
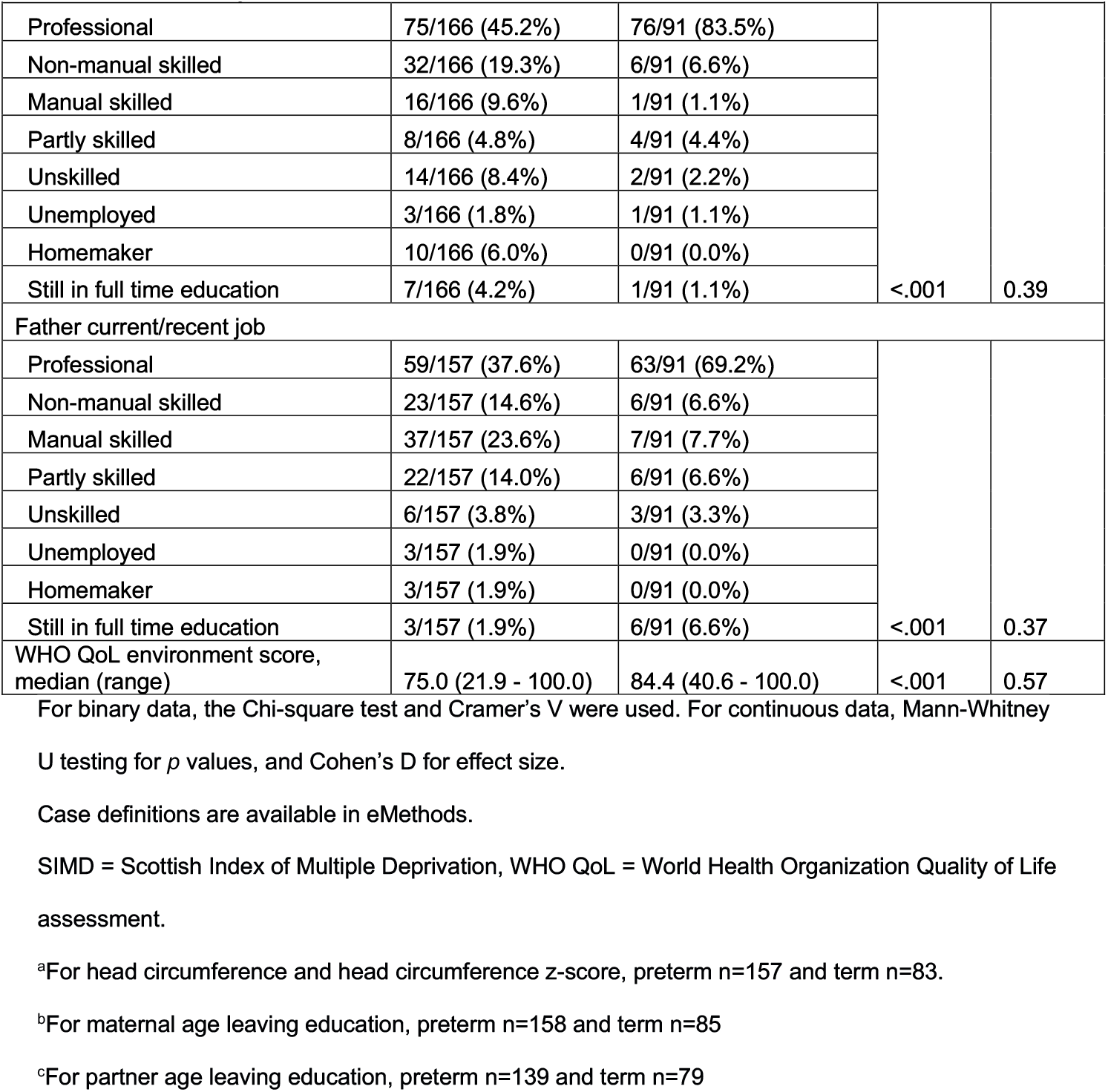
Participant characteristics.

| Characteristic | Preterm (n=170) | Term (n=91) | p value | Effect size |
| --- | --- | --- | --- | --- |
| Gestational age at birth (weeks), median (range) | 30+0 (22+1 - 32+6) | 39+4 (36+3 - 42+1) | <.001 | 5.25 |
| Gestational age at MRI (weeks), median (range) | 40+5 (36+2 - 45+6) | 42+0 (38+2 - 46+1) | <.001 | 0.86 |
| Birthweight (grams), median (range) | 1315 (370 - 2510) | 3460 (2410 - 4560) | <.001 | 4.99 |
| Birthweight z-score, median (range) | 0.13 (-3.13 - 2.14) | 0.45 (-2.30 - 2.57) | <.001 | 0.48 |
| Head circumference (cm), median (range)<sup>a</sup> | 27.5 (17.5 - 33.8) | 35 (32 - 39) | <.001 | 3.66 |
| Head circumference z-score, median (range)<sup>a</sup> | -0.12 (-3.13 - 5.31) | 0.99 (-1.54 - 3.73) | <.001 | 0.97 |
| Sex (male:female) | 95:75 | 50:41 | .211 | 0.01 |
| Maternal smoking during pregnancy | 30/167 (18.0%) | 3/91 (3.3%) | <.001 | 0.21 |
| Multiple pregnancy | 51/170 (30.0%) | 2/91 (2.2%) | <.001 | 0.33 |
| Any breast milk at discharge | 122/167 (73.1%) | 85/91 (93.4%) | <.001 | 0.24 |
| Child ethnicity |  |  |  |  |
| White | 146/166 (88.0%) | 78/86 (90.6%) | .222 | 0.23 |
| Asian | 4/166 (2.4%) | 3/86 (3.5%) |  |  |
| Black | 2/166 (1.2%) | 0/86 (0.0%) |  |  |
| Mixed ethnicity | 9/166 (5.4%) | 5/86 (5.8%) |  |  |
| Other ethnic group | 5/166 (3.0%) | 0/86 (0.0%) |  |  |
| SIMD rank - median (range) | 3913 (6-6929) | 5502 (727-6967) | <.001 | .055 |
| Mother age leaving education (years), median (range)<sup>b</sup> | 20 (14-33) | 23 (16-36) | <.001 | 0.77 |
| Father age leaving education (years), median (range)<sup>c</sup> | 18 (14-34) | 22 (16-36) | <.001 | 0.64 |
| Mother highest educational qualification |  |  |  |  |
| None | 5/165 (3%) | 0/91 (0.0%) | <.001 | 0.36 |
| Basic high school qualification | 18/165 (10.9%) | 5/91 (5.5%) |  |  |
| Advanced high school qualification | 13/165 (7.9%) | 2/91 (2.2%) |  |  |
| College qualification | 42/165 (25.5%) | 9/91 (9.9%) |  |  |
| University undergraduate degree | 44/165 (26.7%) | 40/91 (44.0%) |  |  |
| University postgraduate degree | 34/165 (20.6%) | 35/91 (38.5%) |  |  |
| Not applicable | 9/165 (5.5%) | 0/91 (0.0%) |  |  |
| Father highest educational qualification |  |  |  |  |
| None | 4/148 (2.7%) | 0/90 (0.0%) | <.001 | 0.37 |
| Basic high school qualification | 33/148 (22.3%) | 7/90 (7.8%) |  |  |
| Advanced high school qualification | 13/148 (8.8%) | 5/90 (5.6%) |  |  |
| College qualification | 33/148 (22.3%) | 12/90 (13.3%) |  |  |
| University undergraduate degree | 33/148 (22.3%) | 41/90 (45.6%) |  |  |
| University postgraduate degree | 22/148 (14.9%) | 25/90 (27.8) |  |  |
| Not applicable | 10/148 (6.8%) | 0/90 (0.0%) |  |  |
| Mother current/recent job |  |  |  |  |
| Professional | 75/166 (45.2%) | 76/91 (83.5%) | <.001 | 0.39 |
| Non-manual skilled | 32/166 (19.3%) | 6/91 (6.6%) |  |  |
| Manual skilled | 16/166 (9.6%) | 1/91 (1.1%) |  |  |
| Partly skilled | 8/166 (4.8%) | 4/91 (4.4%) |  |  |
| Unskilled | 14/166 (8.4%) | 2/91 (2.2%) |  |  |
| Unemployed | 3/166 (1.8%) | 1/91 (1.1%) |  |  |
| Homemaker | 10/166 (6.0%) | 0/91 (0.0%) |  |  |
| Still in full time education | 7/166 (4.2%) | 1/91 (1.1%) |  |  |
| Father current/recent job |  |  |  |  |
| Professional | 59/157 (37.6%) | 63/91 (69.2%) | <.001 | 0.37 |
| Non-manual skilled | 23/157 (14.6%) | 6/91 (6.6%) |  |  |
| Manual skilled | 37/157 (23.6%) | 7/91 (7.7%) |  |  |
| Partly skilled | 22/157 (14.0%) | 6/91 (6.6%) |  |  |
| Unskilled | 6/157 (3.8%) | 3/91 (3.3%) |  |  |
| Unemployed | 3/157 (1.9%) | 0/91 (0.0%) |  |  |
| Homemaker | 3/157 (1.9%) | 0/91 (0.0%) |  |  |
| Still in full time education | 3/157 (1.9%) | 6/91 (6.6%) |  |  |
| WHO QoL environment score, median (range) | 75.0 (21.9 - 100.0) | 84.4 (40.6 - 100.0) | <.001 | 0.57 |

For binary data, the Chi-square test and Cramer's V were used. For continuous data, Mann-Whitney
U testing for *p* values, and Cohen's D for effect size.
Case definitions are available in eMethods.
SIMD = Scottish Index of Multiple Deprivation, WHO QoL = World Health Organization Quality of Life assessment.
<sup>a</sup>For head circumference and head circumference z-score, preterm n=157 and term n=83.
<sup>b</sup>For maternal age leaving education, preterm n=158 and term n=85
<sup>c</sup>For partner age leaving education, preterm n=139 and term n=79

There were no significant demographic differences between included and excluded infants, eTable1. The major exposures and comorbidities of the preterm group are shown in eTable 2. Removal of all but one sibling per family from the sample and re-running analyses yielded a similar pattern of results, so the whole sample is included in reported analyses.

### Associations between regional brain volumes, prematurity, and neighborhood deprivation

GA associated with more regional brain volumes than SIMD (McNemar’s test comparing frequency of associations *p*=<.001, Table 2 and eTable 3). After Benjamini-Hochberg correction, GA correlated with the volume of 22/85 (26%) parcels (β range |-0.13| to |0.22|) compared to 1/85 (1.2%) parcel for SIMD (β=0.17). GA-associated parcels were within gray and white matter, often bilaterally, and predominantly had a positive association (17/19 tissue (non-CSF) regions, 89.5%) meaning that higher GA at birth was associated with increased tissue volume. There were negative associations between GA and volumes of CSF spaces (bilateral lateral ventricles and extracerebral CSF). The single parcel associated with SIMD was the right anterior medial and inferior temporal gyri (white matter) (β=0.17, *p*=.03). There were no interactions between GA and SIMD for any brain region.

**Table 2.**
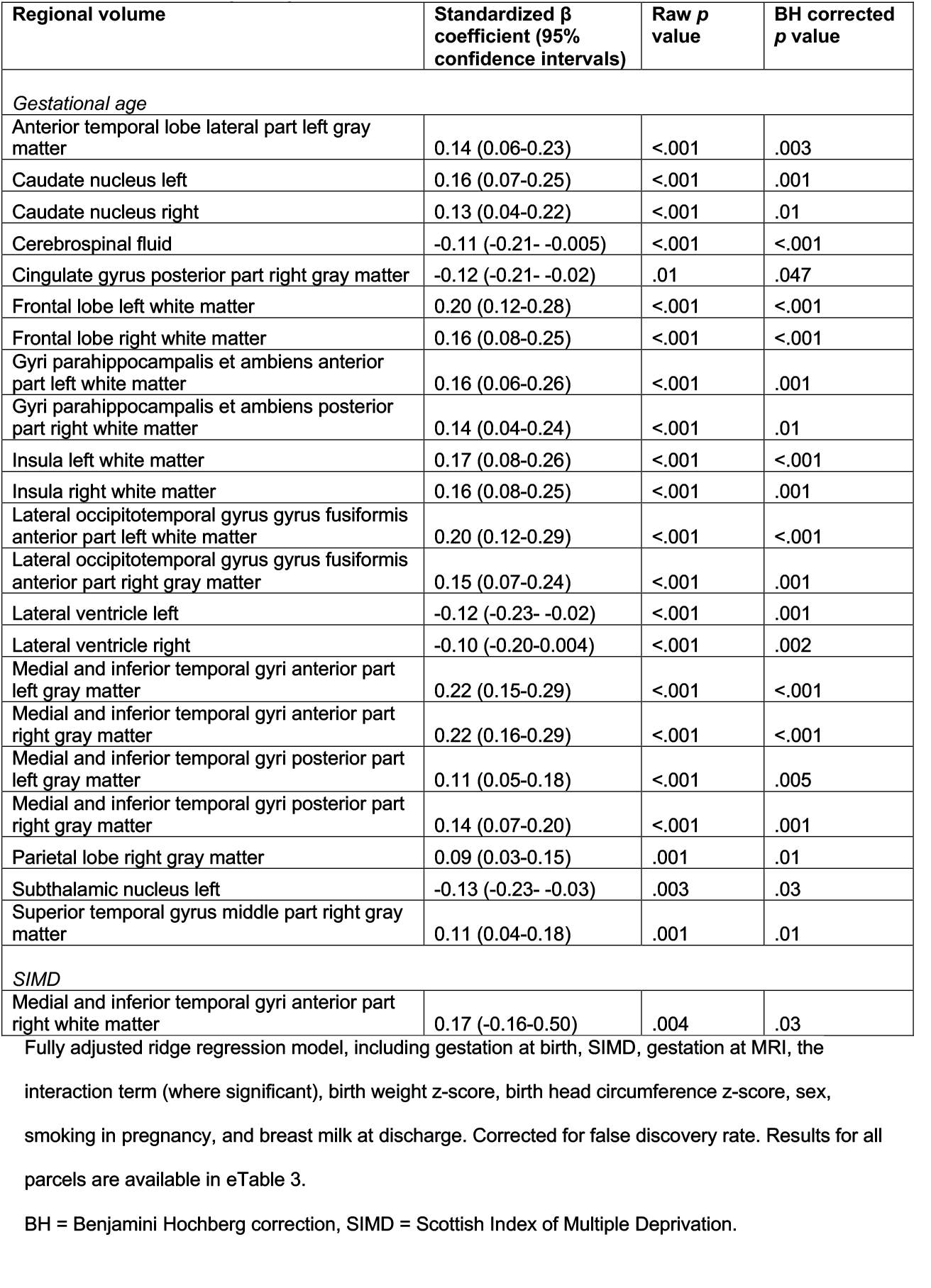
Regional brain volumes with significant relationship with either gestational age or the Scottish Index of Multiple Deprivation.

Fully adjusted models showed a similar profile of results to baseline models (eTable 4), with GA-parcel volume associations more widely distributed than SIMD-parcel volume associations; the number of associations was higher for GA than SIMD: 50/85 (58.8%) and 5/85 (5.9%), respectively.

### Associations between global cortical measures, prematurity, and neighborhood deprivation

GA was associated with cortical surface area (β=0.10 [95% CI 0.02-0.18], *p*=.03) and gyrification index (β=0.16 [95% CI 0.07-0.25], *p*=<.001), whereas SIMD was not associated with any measure of cortical morphology (Table 3). There were no interaction effects between GA and SIMD for any global cortical measure.

**Table 3.**
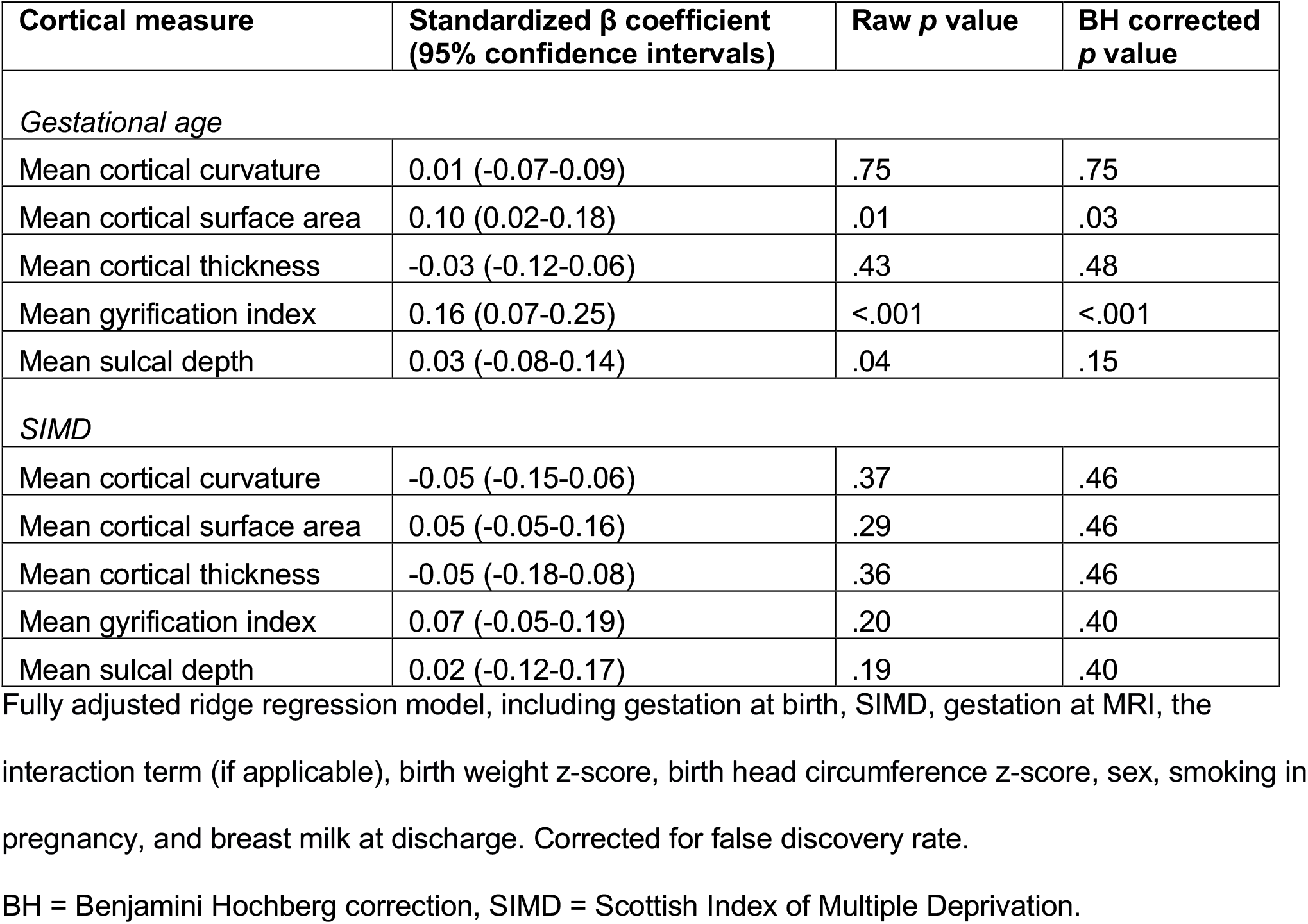
Cortical measures – relationship with gestational age and the Scottish Index of Multiple Deprivation.

| Cortical measure | Standardized $\beta$ coefficient<br>(95% confidence intervals) | Raw $p$ value | BH corrected<br>$p$ value |
| --- | --- | --- | --- |
| *Gestational age* |  |  |  |
| Mean cortical curvature | 0.01 (-0.07-0.09) | .75 | .75 |
| Mean cortical surface area | 0.10 (0.02-0.18) | .01 | .03 |
| Mean cortical thickness | -0.03 (-0.12-0.06) | .43 | .48 |
| Mean gyrification index | 0.16 (0.07-0.25) | <.001 | <.001 |
| Mean sulcal depth | 0.03 (-0.08-0.14) | .04 | .15 |
| *SIMD* |  |  |  |
| Mean cortical curvature | -0.05 (-0.15-0.06) | .37 | .46 |
| Mean cortical surface area | 0.05 (-0.05-0.16) | .29 | .46 |
| Mean cortical thickness | -0.05 (-0.18-0.08) | .36 | .46 |
| Mean gyrification index | 0.07 (-0.05-0.19) | .20 | .40 |
| Mean sulcal depth | 0.02 (-0.12-0.17) | .19 | .40 |

Fully adjusted ridge regression model, including gestation at birth, SIMD, gestation at MRI, the
interaction term (if applicable), birth weight z-score, birth head circumference z-score, sex, smoking in pregnancy, and breast milk at discharge. Corrected for false discovery rate.
BH = Benjamini Hochberg correction, SIMD = Scottish Index of Multiple Deprivation.

Fully adjusted models revealed a similar profile of results to the baseline linear regression models, with GA associations in 2/25 (40%) of cortical measures and no associations with SIMD (eTable 5).

### Correlations between neighborhood-, family-, and subjective-level SES measures

Figure 1A shows that correlations between SES measures ranged from very weak (e.g., between age of father leaving education and the WHO QoL environment score, r=0.07) to strong (e.g., age of father and mother leaving education and final education qualification r=0.7 and 0.64, respectively). Correlations between SIMD rank and parental education/occupation were weak to moderate (r=0.22-0.40).

**Figure 1.**
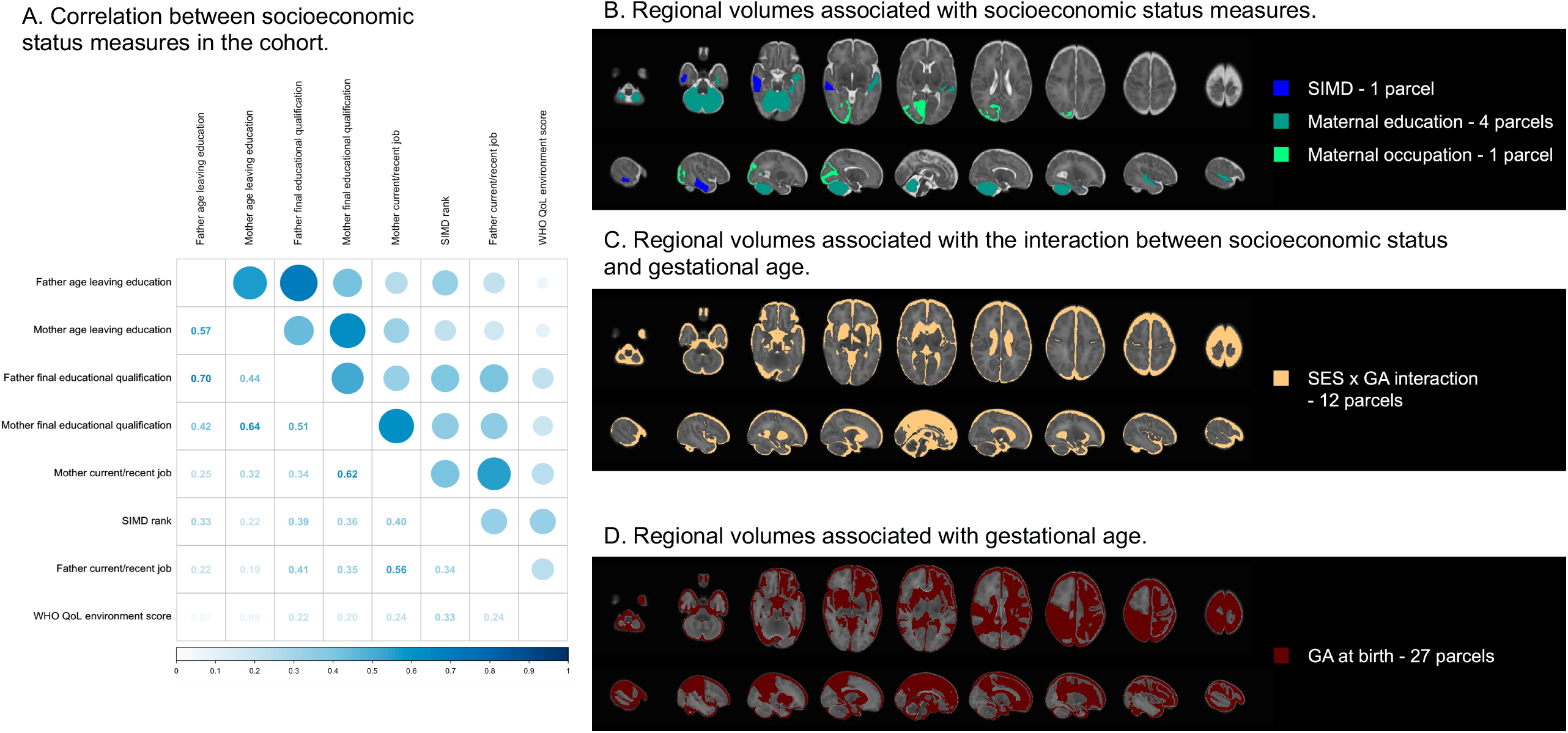
Socioeconomic status measures. A: Spearman rank correlations between socioeconomic status measures in the cohort, showing coefficients when *p*<0.05. B: Parcels associated with socioeconomic status measures. C: Parcels associated with the interaction between socioeconomic status and gestational age. D: Parcels associated with gestational age at birth. B-D show all regions/parcels that were significantly associated with respective measures after Benjamini-Hochberg correction in fully adjusted ridge regression models. GA = gestational age, SES = socioeconomic status, SIMD = Scottish Index of Multiple Deprivation, WHO QoL = World Health Organization Quality of Life assessment.

### Associations between brain features and family- and subjective-level SES measures

In exploratory analyses defining SES by parental education/occupation and subjective measures, a similar pattern of results was obtained as with SIMD: a greater proportion of brain volumes were associated with GA than SES (McNemar’s test comparing frequency of associations *p*=<.001 for all SES measures; Table 4, eTables 6-10, Figure 1B-D). However, there were differences in associations between SES and brain structure depending on the SES measure used. Specifically, maternal education was associated with volume of 4 parcels (left and right cerebella, left middle superior temporal gyrus, and left anterior lateral occipitotemporal gyrus/gyrus fusiformis, β range |0.09| to |0.15|, eTable 6), whereas neighborhood SES (Table 2) and maternal occupation (eTable 8) were each associated with one parcel volume: right anterior medial and inferior temporal gyri (white matter) (β=0.17, *p*=.03), and right occipital lobe (gray matter) (β=0.06, *p*=.0496), respectively. Associations between parcel volumes and family-level SES were positive, meaning that higher family-level SES was associated with increased regional tissue volume.

**Table 4.** Summary table of associations between gestational age and neighborhood-level, family-level and subjective-level measures of socioeconomic status with brain features.

|  | Gestational age | Socioeconomic status measure | Interaction effect |
| --- | --- | --- | --- |
| *Proportion of brain volumes associated with GA and SES* |  |  |  |
| SIMD | 22/85 (25.9%) | 1/85 (1.2%) | 0/85 (0.0%) |
| Maternal education | 15/85 (17.6%) | 4/85 (4.7%) | 5/85 (5.9%) |
| Paternal education | 22/85 (25.9%) | 0/85 (0.0%) | 7/85 (8.2%) |
| Maternal occupation | 12/85 (14.1%) | 1/85 (1.2%) | 10/85 (11.8%) |
| Paternal occupation | 21/85 (24.7%) | 0/85 (0.0%) | 5/85 (5.9%) |
| Subjective socioeconomic status | 17/85 (20.0%) | 0/85 (0.0%) | 4/85 (4.7%) |
| *Proportion of cortical features associated with GA and SES* |  |  |  |
| SIMD | 2/5 (40.0%) | 0/5 (0.0%) | 0/5 (0.0%) |
| Maternal education | 0/5 (0.0%) | 0/5 (0.0%) | 0/5 (0.0%) |
| Paternal education | 0/5 (0.0%) | 0/5 (0.0%) | 0/5 (0.0%) |
| Maternal occupation | 0/5 (0.0%) | 0/5 (0.0%) | 0/5 (0.0%) |
| Paternal occupation | 0/5 (0.0%) | 0/5 (0.0%) | 0/5 (0.0%) |
| Subjective socioeconomic status | 1/5 (20.0%) | 0/5 (0.0%) | 0/5 (0.0%) |

Results summary from ridge regression models for each socioeconomic status measure, for the 85
regional brain volumes and 5 mean cortical measures. Significance threshold is $p < .05$ following
Benjamini-Hochberg correction. Full results are available in Table 3 and eTables 3 and 6-15.
GA = gestational age, SES = socioeconomic status, SIMD = Scottish Index of Multiple Deprivation.

GA-volume associations for paternal education and occupation were 22/85 (25.9%) and 21/85 (24.7%), for maternal education and occupation 15/85 (17.6%) and 12/85 (14.1%), and subjective SES 17/85 (20.0%) respectively (Table 4).

In contrast to SIMD, there were significant interactions between family- and subjective-level SES measures and GA for several brain structures (eTable 6-15, Figure 1C). For family-level and subjective SES measures, the impact of SES was smaller in children with higher GA for extracerebral CSF and left and right lateral ventricular volumes (β range |-0.05| to |-0.001|). The direction of the interaction effect varied for those involving white and gray tissue parcels, such as left anterior gyri parahippocampalis et ambiens (white matter), left caudate nucleus, and left subthalamic nucleus.

For the 5 global cortical measures (eTables 11-15), there were no associations with family-level SES in the fully adjusted model. However, subjective-level SES associated with mean gyrification index (β=0.16 [95% CI 0.07-0.26], *p*=.002).

## Discussion

In a high-income setting, low GA at birth and SES both contribute to regional alterations in brain structure that are apparent at the end of neonatal care. Low GA is associated with widely distributed alterations in brain structure and cortical morphology, whereas effects associated with SES are less widely distributed. We found that neighborhood-, family-, and subjective-level measures of SES are only weakly to moderately correlated; of these, family-level measures (parental education and occupation) associated with more alterations in brain structure than subjective SES and neighborhood deprivation. Furthermore, there is an interaction effect between low GA and family and subjective-level SES measures. These results suggest that atypical brain development seen in preterm infants is driven predominantly by GA at birth, but prematurity does not override SES-brain structure patterning, so interventions designed to attenuate family-level socioeconomic disadvantage in the perinatal period^44^ could promote healthier brain development in preterm infants.

Our results are consistent with studies suggesting that SES may modify the relationship between preterm birth and neurodevelopmental and educational outcomes, especially in the context of preterm brain injury^45–49^. Although few studies have reported the impact of SES on brain morphology of preterm infants, there is some consensus that socioeconomic factors play a role. In a large study of US infants with GA range 27-42 weeks who were scanned between term and 4 months, higher SES, indexed by parental education, had marginal associations with brain volume in infancy and paternal education associated with gray matter volume after accounting for birthweight. However, extremely preterm infants were not represented in this study and findings could have been confounded by post-neonatal intensive care unit (NICU) exposures^47^. In an Australian cohort, a multidimensional measure of social risk was associated with global and regional brain volumes in a group of preterm infants at term-equivalent age but the strength of association varied by GA category (strongest for late preterm [34-36 weeks of gestation] and full-term infants compared with moderate or very preterm infants [<34 weeks of gestation]), and was diminished in multivariable models adjusting for poor intrauterine growth, multiple birth and male sex^49^. Our results indicate that although low GA affects the widest distribution of parcels, SES effects are detectable at the end of NICU care after adjustment for GA at birth, age at MRI, birth weight z-score, birth head circumference z-score, sex, smoking in pregnancy, and nutrition during NICU care.

The regional volumes associated with low GA are in white and gray matter bilaterally, with concurrent increases in extra-axial and lateral ventricular CSF spaces. This pattern is consistent with the cerebral signature of preterm birth, which includes altered cortical morphology, focal white matter volume loss, and reduced deep gray matter volume^5^. By using a contemporary atlas^50^, we have added anatomic granularity to the preterm brain structural phenotype. In adjusted models, regions associated with SES were right anterior medial and inferior temporal gyri (white matter) (SIMD), right occipital lobe (gray matter) (maternal occupation), left and right cerebella, left middle superior temporal gyrus, and left anterior lateral occipitotemporal gyrus/gyrus fusiformis (maternal education). SES measures have previously shown variable association in childhood with the cerebellum^25,46,51^, occipital lobe^25,52^, and temporal lobe^24,25,52^, and the middle temporal gyri and occipitotemporal regions specifically have been positively associated with a composite SES of maternal education/occupation^53^. The possible functions of these SES-associated regions include memory, visual information processing, speech, motor control, balance, and cognition^54^.

Most studies examining relationships between SES and brain morphology have used individual or family measures of SES (parental education/occupation, or family income)^46–48,52,55–57^, or a composite^49,58^, and only one included neighborhood deprivation^58^. We found that family-level SES measures were most closely associated measures with brain structure. This could be explained by family-level measures capturing shared genetic determinants of brain anatomy and exposures to stress, nutrition, or smoking that influence brain development^36,59,60^, which are not fully captured by neighborhood deprivation. Neighborhood-level SES is associated with functional changes in later infancy within this cohort, including preference to view social stimuli^61^ and emotional regulation and cortisol response^62^, suggesting that neighborhood deprivation could have a greater impact after discharge from hospital.

## Strengths and limitations

The study has the following strengths. First, the preterm infants did not have focal parenchymal brain injuries so are representative of the majority of survivors of modern intensive care in high income countries. Second, we assessed whole brain anatomy and global cortical structure using an open-source age-specific atlas. Third, we adjusted for real and potential confounders of neonatal brain structure and used ridge regression to mitigate the problem of multicollinearity. Fourth, we explored SES-brain effects with SES operationalized at different levels, showing that family and individual measures interact with the effects of GA on the brain. Finally, we adhered to a pre-registered analysis plan reducing the risk of false positive results.

The study has some limitations. Although the study population is comparable to neonatal populations in high-income settings, the results may not be generalizable to settings with different socioeconomic profiles. Our choice to study morphology was based on existing literature. We did not investigate possible SES associations with structural or functional connectivity, or other metrics of early brain development such as network complexity^63^ or brain age^64^; future research could investigate whether these image phenotypes map with greater/lesser effect size to SES. Finally, we cannot comment on whether SES patterning of brain structure is dynamic through childhood after preterm birth. The consensus for associations between SES and brain structure in older children and adolescents is much greater^65,66^ than it is for neonates^46–49,52,55–58^. By inference, SES impacts, which were relatively modest in neonates in comparison to low GA, might accumulate through childhood. Of note, functional outcome data from preterm infants suggest the importance of SES increases after the age of 5 years while the importance of birth events as determinants diminishes^67^. To address this question, longitudinal imaging of participants is planned.

## Conclusions

In comparison to socioeconomic factors, low GA is associated with more widely distributed measures of brain structure in preterm infants at term-equivalent age. However, family-level SES measures of parental education and occupation are associated with neonatal brain development, and they interact with low GA. This suggests that strategies designed to mitigate the adverse effects of family-level disadvantage during neonatal intensive care could improve the brain development of preterm infants. Further research is warranted to understand the biological mechanisms that embed preterm birth and level-specific social disadvantage in brain development.

## Data Availability

A copy of the dataset will be placed in the University of Edinburgh DataVault (https://datavault.ed.ac.uk/). Requests for access should be made to the TEBC Chief Investigator (Professor J Boardman).

## Acknowledgements

This work was supported by Theirworld (www.theirworld.org) and was carried out in the Medical Research Council Centre for Reproductive Health, which was funded by Medical Research Council Centre Grant (MRC G1002033). Participants were scanned in the University of Edinburgh Imaging Research MRI Facility at the Royal Infirmary of Edinburgh, which was established with funding from The Wellcome Trust, Dunhill Medical Trust, Edinburgh and Lothians Research Foundation, Theirworld, The Muir Maxwell Trust and other sources. KV and LJ-S are funded by the Wellcome Translational Neuroscience PhD Programme at the University of Edinburgh (108890/Z/15/Z). The authors are grateful to the families who consented to take part in the study and to all the University’s imaging research staff for providing the infant scanning.

## Notes

### Competing Interest Statement

The authors have declared no competing interest.

### Clinical Protocols

https://bmjopen.bmj.com/content/10/3/e035854

https://osf.io/ftv2s

### Author Declarations

Ethical approval was obtained from the UK National Research Ethics Service (South East Scotland Research Ethic Committee 16/SS/0154).

## References

1. Chawanpaiboon S, Vogel JP, Moller AB, et al. Global, regional, and national estimates of levels of preterm birth in 2014: a systematic review and modelling analysis. Lancet Global Heal. 2019;7(1):e37–e46. doi:10.1016/s2214-109x(18)30451-0

2. Johnson S, Marlow N. Early and long-term outcome of infants born extremely preterm. Archives of Disease in Childhood. 2017;102(1):97–102. doi:10.1136/archdischild-2015-309581

3. Agrawal S, Rao SC, Bulsara MK, Patole SK. Prevalence of Autism Spectrum Disorder in Preterm Infants: A Meta-analysis. Pediatrics. 2018;142(3):1–14.

4. Twilhaar ES, Wade RM, Kieviet JF de, Goudoever JB van, Elburg RM van, Oosterlaan J. Cognitive Outcomes of Children Born Extremely or Very Preterm Since the 1990s and Associated Risk Factors: A Meta-analysis and Meta-regression. Jama Pediatr. 2018;172(4):361. doi:10.1001/jamapediatrics.2017.5323

5. Batalle D, Edwards AD, O’Muircheartaigh J. Annual Research Review: Not just a small adult brain: understanding later neurodevelopment through imaging the neonatal brain. J Child Psychol Psyc. 2018;59(4):350–371. doi:10.1111/jcpp.12838

6. Boardman JP, Counsell SJ, Rueckert D, et al. Early growth in brain volume is preserved in the majority of preterm infants. Ann Neurol. 2007;62(2):185–192. doi:10.1002/ana.21171

7. Romberg J, Wilke M, Allgaier C, et al. MRI-based brain volumes of preterm infants at term: a systematic review and meta-analysis. Archives Dis Child - Fetal Neonatal Ed. Published online 2022:fetalneonatal-2021-322846. doi:10.1136/archdischild-2021-322846

8. Dimitrova R, Pietsch M, Ciarrusta J, et al. Preterm birth alters the development of cortical microstructure and morphology at term-equivalent age. Neuroimage. 2021;243:118488. doi:10.1016/j.neuroimage.2021.118488

9. Thompson DK, Matthews LG, Alexander B, et al. Tracking regional brain growth up to age 13 in children born term and very preterm. Nat Commun. 2020;11(1):696. doi:10.1038/s41467-020-14334-9

10. Boardman JP, Craven C, Valappil S, et al. A common neonatal image phenotype predicts adverse neurodevelopmental outcome in children born preterm. NeuroImage. 2010;52(2):409–414. doi:10.1016/j.neuroimage.2010.04.261

11. Dubois J, Benders M, Borradori-Tolsa C, et al. Primary cortical folding in the human newborn: an early marker of later functional development. Brain. 2008;131(8):2028–2041. doi:10.1093/brain/awn137

12. Kapellou O, Counsell SJ, Kennea N, et al. Abnormal Cortical Development after Premature Birth Shown by Altered Allometric Scaling of Brain Growth. Plos Med. 2006;3(8):e265. doi:10.1371/journal.pmed.0030265

13. Farah MJ. The Neuroscience of Socioeconomic Status: Correlates, Causes, and Consequences. Neuron. 2017;96(1):56–71. doi:10.1016/j.neuron.2017.08.034

14. Johnson SB, Riis JL, Noble KG. State of the Art Review: Poverty and the Developing Brain. Pediatrics. 2016;137(4):e20153075. doi:10.1542/peds.2015-3075

15. von Stumm S, Plomin R. Socioeconomic status and the growth of intelligence from infancy through adolescence. Intelligence. 2015;48:30–36. doi:10.1016/j.intell.2014.10.002

16. Farah MJ, Shera DM, Savage JH, et al. Childhood poverty: Specific associations with neurocognitive development. Brain Res. 2006;1110(1):166–174. doi:10.1016/j.brainres.2006.06.072

17. Noble KG, Norman MF, Farah MJ. Neurocognitive correlates of socioeconomic status in kindergarten children. Developmental Sci. 2005;8(1):74–87. doi:10.1111/j.1467-7687.2005.00394.x

18. McLoyd VC. Socioeconomic Disadvantage and Child Development. Am Psychol. 1998;53(2):185–204. doi:10.1037/0003-066x.53.2.185

19. Duncan GJ, Yeung WJ, Brooks-Gunn J, Smith JR. How Much Does Childhood Poverty Affect the Life Chances of Children? Am Sociol Rev. 1998;63(3):406. doi:10.2307/2657556

20. Duncan GJ, Ziol-Guest KM, Kalil A. Early-Childhood Poverty and Adult Attainment, Behavior, and Health. Child Dev. 2010;81(1):306–325. doi:10.1111/j.1467-8624.2009.01396.x

21. Pillas D, Marmot M, Naicker K, Goldblatt P, Morrison J, Pikhart H. Social inequalities in early childhood health and development: a European-wide systematic review. Pediatr Res. 2014;76(5):418–424. doi:10.1038/pr.2014.122

22. Noble KG, Engelhardt LE, Brito NH, et al. Socioeconomic disparities in neurocognitive development in the first two years of life. Dev Psychobiol. 2015;57(5):535–551. doi:10.1002/dev.21303

23. Mackey AP, Finn AS, Leonard JA, et al. Neuroanatomical Correlates of the Income-Achievement Gap. Psychol Sci. 2015;26(6):925–933. doi:10.1177/0956797615572233

24. Hair NL, Hanson JL, Wolfe BL, Pollak SD. Association of Child Poverty, Brain Development, and Academic Achievement. Jama Pediatr. 2015;169(9):822–829. doi:10.1001/jamapediatrics.2015.1475

25. Gur RE, Moore TM, Rosen AFG, et al. Burden of Environmental Adversity Associated With Psychopathology, Maturation, and Brain Behavior Parameters in Youths. Jama Psychiat. 2019;76(9):966–975. doi:10.1001/jamapsychiatry.2019.0943

26. Boardman JP, Hall J, Thrippleton MJ, et al. Impact of preterm birth on brain development and long-term outcome: protocol for a cohort study in Scotland. Bmj Open. 2020;10:e035854. doi:10.1136/bmjopen-2019-035854

27. Scottish National Statistics. SIMD - Scottish Index of Multiple Deprivation: SIMD16 Technical Notes. Scottish National Statistics; 2016:1-69.

28. WHO. World Health Organization Quality of Life Assessment: WHOQOL-BREF. World Health Organization; 1996.

29. Minh A, Muhajarine N, Janus M, Brownell M, Guhn M. A review of neighborhood effects and early child development: How, where, and for whom, do neighborhoods matter? Health Place. 2017;46:155–174. doi:10.1016/j.healthplace.2017.04.012

30. Mckinnon K, Galdi P, Cabez MB, et al. The impact of preterm birth and socioeconomic status on neonatal brain morphology. OSF. Published online 2022. doi:10.17605/osf.io/ftv2s

31. Miller SL, Huppi PS, Mallard C. The consequences of fetal growth restriction on brain structure and neurodevelopmental outcome. J Physiology. 2016;594(4):807–823. doi:10.1113/jp271402

32. Thompson DK, Warfield SK, Carlin JB, et al. Perinatal risk factors altering regional brain structure in the preterm infant. Brain. 2007;130(3):667–677. doi:10.1093/brain/awl277

33. Selvanathan T, Guo T, Kwan E, et al. Head circumference, total cerebral volume and neurodevelopment in preterm neonates. Archives Dis Child - Fetal Neonatal Ed. Published online 2021:fetalneonatal-2020-321397. doi:10.1136/archdischild-2020-321397

34. Lehtola SJ, Tuulari JJ, Karlsson L, et al. Associations of age and sex with brain volumes and asymmetry in 2–5-week-old infants. Brain Struct Funct. 2019;224(1):501–513. doi:10.1007/s00429-018-1787-x

35. Roustaei Z, Räisänen S, Gissler M, Heinonen S. Associations between maternal age and socioeconomic status with smoking during the second and third trimesters of pregnancy: a register-based study of 932 671 women in Finland from 2000 to 2015. Bmj Open. 2020;10(8):e034839. doi:10.1136/bmjopen-2019-034839

36. Ekblad M, Korkeila J, Lehtonen L. Smoking during pregnancy affects foetal brain development. Acta Paediatr. 2015;104(1):12–18. doi:10.1111/apa.12791

37. Blesa M, Sullivan G, Anblagan D, et al. Early breast milk exposure modifies brain connectivity in preterm infants. Neuroimage. 2019;184:431–439. doi:10.1016/j.neuroimage.2018.09.045

38. Sullivan G, Vaher K, Blesa M, et al. Breast milk exposure is associated with cortical maturation in preterm infants. Ann Neurol. Published online 2022. doi:10.1002/ana.26559

39. Makropoulos A, Robinson EC, Schuh A, et al. The developing human connectome project: A minimal processing pipeline for neonatal cortical surface reconstruction. NeuroImage. 2018;173:88–112. doi:10.1016/j.neuroimage.2018.01.054

40. R Core Team. R: A Language and Environment for Statistical Computing. R Foundation for Statistical Computing; 2022. https://www.R-project.org/

41. Hastie T, Tibshirani R, Friedman J. The Elements of Statistical Learning: Data Mining, Inference, and Prediction. 2nd Edition. Springer; 2009.

42. Benjamini Y, Hochberg Y. Controlling the False Discovery Rate: A Practical and Powerful Approach to Multiple Testing. Journal of the Royal Statistical Society. 1995;57(1):289–300.

43. National Records of Scotland. Scotland’s Census. Published 2020. Accessed December 2, 2021. https://www.scotlandscensus.gov.uk/

44. Noble KG, Magnuson K, Gennetian LA, et al. Baby’s First Years: Design of a Randomized Controlled Trial of Poverty Reduction in the United States. Pediatrics. 2021;148(4):e2020049702. doi:10.1542/peds.2020-049702

45. Benavente-Fernández I, Siddiqi A, Miller SP. Socioeconomic status and brain injury in children born preterm: modifying neurodevelopmental outcome. Pediatr Res. 2020;87(2):391–398. doi:10.1038/s41390-019-0646-7

46. Stiver ML, Kamino D, Guo T, et al. Maternal Postsecondary Education Associated With Improved Cerebellar Growth After Preterm Birth. J Child Neurol. 2015;30(12):1633–1639. doi:10.1177/0883073815576790

47. Knickmeyer RC, Xia K, Lu Z, et al. Impact of Demographic and Obstetric Factors on Infant Brain Volumes: A Population Neuroscience Study. Cereb Cortex. 2017;27(12):5616–5625. doi:10.1093/cercor/bhw331

48. Gui L, Loukas S, Lazeyras F, Hüppi PS, Meskaldji DE, Tolsa CB. Longitudinal study of neonatal brain tissue volumes in preterm infants and their ability to predict neurodevelopmental outcome. Neuroimage. 2018;185:728–741. doi:10.1016/j.neuroimage.2018.06.034

49. Thompson DK, Kelly CE, Chen J, et al. Early life predictors of brain development at term-equivalent age in infants born across the gestational age spectrum. Neuroimage. 2018;185:813–824. doi:10.1016/j.neuroimage.2018.04.031

50. Makropoulos A, Aljabar P, Wright R, et al. Regional growth and atlasing of the developing human brain. Neuroimage. 2016;125:456–478. doi:10.1016/j.neuroimage.2015.10.047

51. Machlin L, McLaughlin KA, Sheridan MA. Brain structure mediates the association between socioeconomic status and attention-deficit/hyperactivity disorder. Developmental Sci. 2020;23(1):e12844. doi:10.1111/desc.12844

52. Spann M, Bansal R, Hao X, Rosen T, Peterson B. Prenatal socioeconomic status and social support are associated with neonatal brain morphology, toddler language and psychiatric symptoms. Child Neuropsychology. 2020;26(2):170–188.

53. Jednoróg K, Altarelli I, Monzalvo K, et al. The Influence of Socioeconomic Status on Children’s Brain Structure. Plos One. 2012;7(8):e42486. doi:10.1371/journal.pone.0042486

54. Nieuwenhuys R, Voogd J, Huijzen C van. The Human Central Nervous System. 4th Edition. Springer; 2008. doi:10.1007/978-3-540-34686-9

55. Kersbergen KJ, Leroy F, Išgum I, et al. Relation between clinical risk factors, early cortical changes, and neurodevelopmental outcome in preterm infants. NeuroImage. 2016;142(C):301–310. doi:10.1016/j.neuroimage.2016.07.010

56. Jha SC, Xia K, Ahn M, et al. Environmental Influences on Infant Cortical Thickness and Surface Area. Cereb Cortex. 2019;29(3):1139–1149. doi:10.1093/cercor/bhy020

57. Betancourt LM, Avants B, Farah MJ, et al. Effect of socioeconomic status (SES) disparity on neural development in female African-American infants at age 1 month. Developmental Sci. 2016;19(6):947–956. doi:10.1111/desc.12344

58. Triplett RL, Lean RE, Parikh A, et al. Association of Prenatal Exposure to Early-Life Adversity With Neonatal Brain Volumes at Birth. Jama Netw Open. 2022;5(4):e227045. doi:10.1001/jamanetworkopen.2022.7045

59. DiPietro JA. Maternal Stress in Pregnancy: Considerations for Fetal Development. J Adolescent Health. 2012;51(2):S3–S8. doi:10.1016/j.jadohealth.2012.04.008

60. Baron R, Manniën J, Velde SJ te, Klomp T, Hutton EK, Brug J. Socio-demographic inequalities across a range of health status indicators and health behaviours among pregnant women in prenatal primary care: a cross-sectional study. Bmc Pregnancy Childb. 2015;15(1):261. doi:10.1186/s12884-015-0676-z

61. Dean B, Ginnell L, Ledsham V, et al. Eye-tracking for longitudinal assessment of social cognition in children born preterm. J Child Psychol Psyc. 2021;62(4):470–480. doi:10.1111/jcpp.13304

62. Ginnell L, O’Carroll S, Ledsham V, et al. Emotion regulation and cortisol response to the still-face procedure in preterm and full-term infants. Psychoneuroendocrino. 2022;141:105760. doi:10.1016/j.psyneuen.2022.105760

63. Blesa M, Galdi P, Cox SR, et al. Hierarchical Complexity of the Macro-Scale Neonatal Brain. Cereb Cortex. 2020;31(4):bhaa345.. doi:10.1093/cercor/bhaa345

64. Galdi P, Blesa M, Stoye DQ, et al. Neonatal morphometric similarity mapping for predicting brain age and characterizing neuroanatomic variation associated with preterm birth. Neuroimage Clin. 2020;25:102195. doi:10.1016/j.nicl.2020.102195

65. Rakesh D, Whittle S. Socioeconomic status and the developing brain – a systematic review of neuroimaging findings in youth. Neurosci Biobehav Rev. 2021;130:379–407. doi:10.1016/j.neubiorev.2021.08.027

66. Gard AM, Maxwell AM, Shaw DS, et al. Beyond family-level adversities: Exploring the developmental timing of neighborhood disadvantage effects on the brain. Developmental Sci. 2021;24(1):e12985. doi:10.1111/desc.12985

67. Linsell L, Malouf R, Morris J, Kurinczuk JJ, Marlow N. Prognostic Factors for Poor Cognitive Development in Children Born Very Preterm or With Very Low Birth Weight: A Systematic Review. Jama Pediatr. 2015;169(12):1–11. doi:10.1001/jamapediatrics.2015.2175

