## Supplementary Text for "Integrated Analysis of Preterm Birth and Socioeconomic Status with Neonatal Brain Structure"

### Integrated Analysis of Preterm Birth and Socioeconomic Deprivation with Neonatal Brain Structure – Supplementary Information

Katie Mckinnon, Paola Galdi, Manuel Blesa, Gemma Sullivan, Kadi Vaher, Amy Corrigan, Jill Hall, Lorena Jimenez Sanchez, Michael Thrippleton, Mark Bastin, Alan J. Quigley, Evdoxia Valavani, Athanasios Tsanas, Hilary Richardson, James P. Boardman

**eMethods:** Case definition and sources.
**eFigure 1**: Flow diagram of participants.
**eTable 1:** Comparison of included and excluded participants.
**eTable 2:** Clinical features of the preterm sample.
**eTable 3:** Regional brain volumes – relationship with gestational age, the Scottish Index of Multiple Deprivation and interaction effect (fully adjusted ridge regression model).
**eTable 4:** Regional brain volumes – relationship with gestational age, the Scottish Index of Multiple Deprivation and interaction effect (baseline unadjusted linear regression model).
**eTable 5:** Cortical measures – relationship with gestational age and the Scottish Index of Multiple Deprivation (baseline unadjusted linear regression model).
**eTable 6:** Regional brain volumes – relationship with gestational age, maternal final educational qualification, and interaction effect (fully adjusted ridge regression model).
**eTable 7:** Regional brain volumes – relationship with gestational age, paternal final educational qualification, and interaction effect (fully adjusted ridge regression model).
**eTable 8:** Regional brain volumes – relationship with gestational age, maternal occupation, and interaction effect (fully adjusted ridge regression model).
**eTable 9:** Regional brain volumes – relationship with gestational age, paternal occupation, and interaction effect (fully adjusted ridge regression model).
**eTable 10:** Regional brain volumes – relationship with gestational age, subjective socioeconomic status, and interaction effect (fully adjusted ridge regression model).
**eTable 11:** Cortical measures – relationship with gestational age and maternal final educational qualification (fully adjusted ridge regression model).
**eTable 12:** Cortical measures – relationship with gestational age and paternal final educational qualification (fully adjusted ridge regression model).
**eTable 13:** Cortical measures – relationship with gestational age, maternal occupation, and interaction effect (fully adjusted ridge regression model).
**eTable 14:** Cortical measures – relationship with gestational age and paternal occupation (ridge regression model).
**eTable 15:** Cortical measures – relationship with gestational age, subjective socioeconomic status, and interaction effect (fully adjusted ridge regression model).

#### eMethods: Case definitions and sources

Gestational age at birth: from medical records, based on dating ultrasound scan at 12 weeks' gestation if available, or first scan beyond this point.
Birthweight: from medical records, measured at birth.
Head circumference: from medical records, measured at birth.
Maternal smoking during pregnancy: yes/no, at any point during pregnancy, parental questionnaire after birth.
Multiple pregnancy: twins or higher multiples, from medical records.
Breast milk at discharge: breast fed or mixed fed at discharge from hospital, parental questionnaire.
Child ethnicity: parental questionnaire after birth.

- White: any White background.
- Asian: Indian, Pakistani, Bangladeshi, Chinese, any other Asian background.
- Black: Caribbean, African, any other African background.
- Mixed ethnicity: White and Black Caribbean, White and Black African, White and Asian, any other mixed background.
- Other ethnic group: any other ethnic group.

Scottish Index of Multiple Deprivation: derived from postcode at birth.
Parent age leaving education: parental questionnaire after birth.
Parent final educational qualification: parental questionnaire after birth.

- None: no qualifications obtained.
- Basic high school qualification: National 5s, Standard Grades, GCSEs (General Certificates of Secondary Education) or equivalent. For the purposes of analysis, this was divided into 1-4 and >5 qualifications.
- Advanced high school qualification: Highers, A levels (Advanced levels) or equivalent.
- College qualification: e.g. National Certificate, Higher National Diploma, Higher National Certificate, vocational qualifications.
- University undergraduate degree.
- University postgraduate degree.
- Not applicable.

Parent current/recent job: parental questionnaire after birth.

- Professional: e.g. doctors, lawyers, teachers, managers.
- Non-manual skilled: e.g. typist, police officer, fireman.
- Manual skilled: e.g. toolmaker, foreman, ambulance man.
- Partly skilled: e.g. bus conductor, postman.
- Unskilled: cleaners, porters, messengers.
- Unemployed: for the majority of adult life.
- Homemaker.
- Still in full time education.

World Health Organization Quality of Life – environment domain: parental questionnaire at term-corrected age (at time of MRI).
Preterm sample clinical features: from medical records.

- Sepsis: positive blood culture and/or physician decision to treat with five days of antibiotics; early-onset is <72 hours after birth and late-onset is after 72 hours.
- Retinopathy of prematurity: requiring treatment.
- Bronchopulmonary dysplasia: requirement for respiratory support and/or supplemental oxygen after 36 weeks' gestation.
- Necrotising enterocolitis: medical (7 days nil by mouth) or surgical management.
- Antenatal steroids: at least one dose of maternal steroids given for fetal lung maturation.
- Antenatal magnesium sulphate: magnesium sulphate given to mother in the antenatal period.
- Postnatal steroids: dexamethasone given to ventilator-dependent infants to facilitate extubation.

#### eFigure 1: Flow diagram of participants

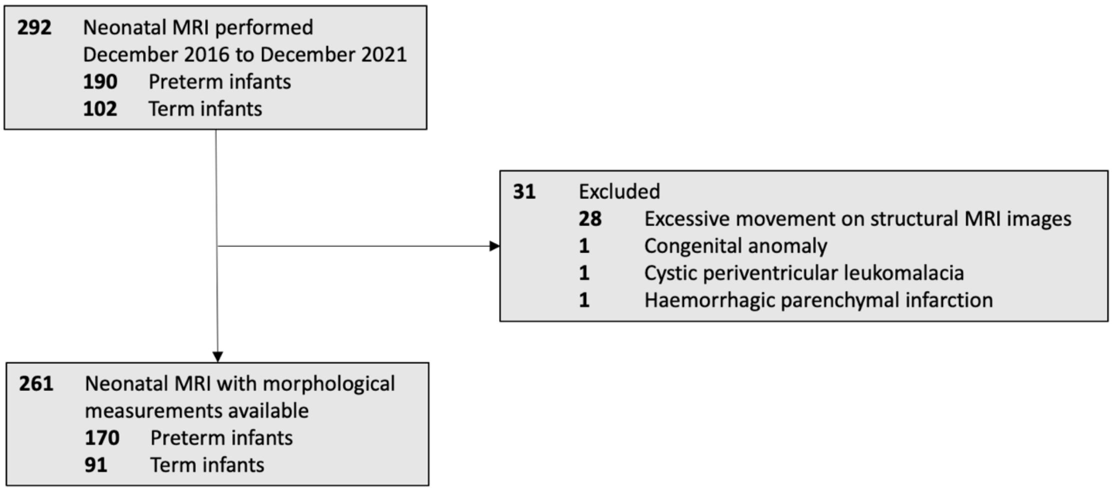


Study exclusion criteria were major congenital malformation, chromosomal abnormality, congenital infection, cystic periventricular leukomalacia, hemorrhagic parenchymal infarction, and post-hemorrhagic ventricular dilatation. In individuals excluded following MRI, these lesions had not been identified on prior routine ultrasound in the neonatal unit.

#### eTable 1: Comparison of included and excluded participants

| Measure | Included in study (n=261) | Excluded from study (n=31) | *p* value |
|---|---|---|---|
| Gestational age at birth (weeks) - median (range) | 31+2 (22+1 - 42+1) | 30+0 (24+0 - 41+4) | .56 |
| Gestational age at MRI (weeks) - median (range) | 41+1 (40+5 - 46+1) | 41+5 (37+7 - 45+1) | .41 |
| Birthweight (grams) - median (range) | 1620 (370 - 4560) | 1445 (454 - 4170) | .89 |
| Birthweight z-score - median (range) | 0.25 (-3.13 - 2.57) | 0.25 (-5.07 - 2.18) | .40 |
| Head circumference (cm) - median (range)[a] | 28.9 (17.5 - 39) | 27.5 (22.1 - 37) | .22 |
| Head circumference z-score - median (range)[a] | 0.14 (-3.13 - 2.57) | 0.077 (-29 - 2.07) | .45 |
| Sex (M:F) | 145:116 | 15:16 | .90 |
| Maternal smoking during pregnancy | 33/258 (12.8%) | 4/31 (12.9%) | .99 |
| Multiple pregnancy | 53/261 (20.3%) | 3/31 (9.7%) | .15 |
| Any breast milk at discharge | 207/258 (80.2%) | 28/31 (90.3%) | .90 |
| Child ethnicity | | | |
| White | 224/252 (88.9%) | 21/30 (70.0%) | .08 |
| Asian | 7/252 (2.8%) | 3/30 (10.0%) | |
| Black | 2/252 (0.8%) | 1/30 (3.3%) | |
| Mixed ethnicity | 14/252 (5.6%) | 4/30 (13.3%) | |
| Other ethnic group | 5/252 (2.0%) | 1/30 (3.3%) | |
| SIMD rank - median (range) | 4510 (6-6967) | 3720 (51 - 6966) | .56 |
| Mother age leaving education (years) - median (range)[b] | 21 (14 - 36) | 22 (16 - 32) | .15 |
| Father age leaving education (years) - median (range)[c] | 21 (14 - 36) | 21 (16 - 36) | .14 |
| Mother highest educational qualification | | | |
| None | 5/256 (2.0%) | 0/31 (0.0%) | .74 |
| Basic high school qualification | 23/256 (9.0%) | 3/31 (9.7%) | |
| Advanced high school qualification | 15/256 (5.9%) | 1/31 (3.2%) | |
| College qualification | 51/256 (19.9%) | 8/31 (25.8%) | |
| University undergraduate degree | 84/256 (32.8%) | 10/31 (32.3%) | |
| University postgraduate degree | 69/256 (27.0%) | 9/31 (29.0%) | |
| Not applicable | 9/256 (3.5%) | 1/31 (3.2%) | |
| Father highest educational qualification | | | |
| None | 4/238 (1.7%) | 0/30 (0.0%) | .50 |
| Basic high school qualification | 40/238 (16.8%) | 4/30 (13.3%) | |
| Advanced high school qualification | 18/238 (7.6%) | 3/30 (10.0%) | |
| College qualification | 45/238 (18.9%) | 5/30 (16.7%) | |
| University undergraduate degree | 74/238 (31.1%) | 8/30 (26.7%) | |
| University postgraduate degree | 47/238 (19.7%) | 10/30 (33.3%) | |
| Not applicable | 10/238 (4.2%) | 0/30 (0.0%) | |

| Measure | Included in study (n=261) | Excluded from study (n=31) | *p* value |
|---|---|---|---|
| Mother current/recent job | | | |
| Professional | 151/257 (58.8%) | 17/31 (54.8%) | .85 |
| Non-manual skilled | 38/257 (14.8%) | 9/31 (29.0%) | |
| Manual skilled | 17/257 (6.6%) | 0/31 (0.0%) | |
| Partly skilled | 12/257 (4.7%) | 1/31 (3.2%) | |
| Unskilled | 16/257 (6.2%) | 1/31 (3.2%) | |
| Unemployed | 4/257 (1.6%) | 1/31 (3.2%) | |
| Homemaker | 10/257 (3.9%) | 1/31 (3.2%) | |
| Still in full time education | 8/257 (3.1%) | 0/31 (0.0%) | |
| Partner current/ recent job | | | |
| Professional | 122/248 (49.2%) | 19/30 (63.3%) | .33 |
| Non-manual skilled | 29/248 (11.7%) | 1/30 (3.3%) | |
| Manual skilled | 44/248 (17.7%) | 6/30 (20%) | |
| Partly skilled | 28/248 (11.3%) | 3/30 (10.0%) | |
| Unskilled | 9/248 (3.6%) | 1/30 (3.3%) | |
| Unemployed | 3/248 (1.2%) | 0/30 (0.0%) | |
| Homemaker | 3/248 (1.2%) | 0/30 (0.0%) | |
| Still in full time education | 9/248 (3.6%) | 0/30 (0.0%) | |
| WHO QoL environment score - median (range)[d] | 78.1 (21.9 - 100.0) | 81.3 (53.1 - 100) | .48 |

*P* values compare individuals included in this study with those excluded following MRI.
For binary data, we used Chi-square testing for *p* values. For continuous data, we used Mann-Whitney U testing for *p* values.
Case definitions are available in eMethods.
[a]For head circumference, and head circumference z-score, included n=240 and excluded n=30.
[b]For maternal age leaving education, included n=241 and excluded n=28.
[c]For partner age leaving education, included n=216 and excluded n=25.
[d]For WHO QoL environment score, included n=242 and excluded n=28.

#### eTable 2. Clinical features of the preterm sample

| Characteristic | Preterm (total n=170) |
|---|---|
| Early-onset sepsis | 14/169 (8.3%) |
| Late-onset sepsis | 27/167 (16.2%) |
| Retinopathy of prematurity | 7/160 (4.4%) |
| Bronchopulmonary dysplasia | 41/168 (24.4%) |
| Necrotizing enterocolitis | 7/166 (4.2%) |
| Antenatal steroids | 162/170 (95.3%) |
| Antenatal magnesium sulfate | 134/170 (78.8%) |
| Postnatal steroids | 6/168 (3.6%) |

Case definitions are available in eMethods. Information not available for all children, for example if transferred to another unit prior to discharge home.

**eTable 3: Regional brain volumes – relationship with gestational age, the Scottish Index of Multiple Deprivation and interaction effect (fully adjusted ridge regression model)**

| Regional volume | Standardized β coefficient (95% confidence intervals) | Raw *p* value | BH corrected *p* value |
|---|---|---|---|
| *Gestation* | | | |
| Amygdala left | 0.08 (-0.02-0.17) | .06 | .20 |
| Amygdala right | 0.02 (-0.07-0.12) | .57 | .70 |
| Anterior temporal lobe lateral part left gray matter | 0.14 (0.06-0.23) | <.001 | .003 |
| Anterior temporal lobe lateral part left white matter | 0.11 (0.01-0.21) | .01 | .08 |
| Anterior temporal lobe lateral part right gray matter | 0.07 (-0.01-0.15) | .08 | .22 |
| Anterior temporal lobe lateral part right white matter | 0.09 (-0.004-0.19) | .02 | .11 |
| Anterior temporal lobe medial part left gray matter | 0.05 (-0.04-0.14) | .26 | .47 |
| Anterior temporal lobe medial part left white matter | 0.03 (-0.07-0.14) | .37 | .53 |
| Anterior temporal lobe medial part right gray matter | 0.05 (-0.04-0.15) | .20 | .42 |
| Anterior temporal lobe medial part right white matter | 0.05 (-0.05-0.15) | .24 | .47 |
| Brainstem | -0.03 (-0.11-0.06) | .52 | .66 |
| Caudate nucleus left | 0.16 (0.07-0.25) | <.001 | <.001 |
| Caudate nucleus right | 0.13 (0.04-0.22) | <.001 | .01 |
| Cerebellum left | -0.01 (-0.08-0.06) | .80 | .87 |
| Cerebellum right | -0.03 (-0.10-0.04) | .38 | .54 |
| Cerebrospinal fluid | -0.11 (-0.21- -0.005) | <.001 | <.001 |
| Cingulate gyrus anterior part left gray matter | 0.01 (-0.09-0.10) | .87 | .92 |
| Cingulate gyrus anterior part left white matter | 0.06 (-0.04-0.16) | .09 | .25 |
| Cingulate gyrus anterior part right gray matter | -0.07 (-0.16-0.03) | .10 | .26 |
| Cingulate gyrus anterior part right white matter | 0.04 (-0.06-0.14) | .27 | .48 |
| Cingulate gyrus posterior part left gray matter | -0.08 (-0.17-0.01) | .051 | .17 |
| Cingulate gyrus posterior part left white matter | 0.05 (-0.04-0.14) | .20 | .42 |
| Cingulate gyrus posterior part right gray matter | -0.12 (-0.21- -0.02) | .01 | .047 |
| Cingulate gyrus posterior part right white matter | 0.07 (-0.01-0.16) | .06 | .20 |
| Corpus callosum | 0.08 (-0.01-0.17) | .046 | .16 |
| Frontal lobe left gray matter | 0.02 (-0.06-0.09) | .63 | .74 |
| Frontal lobe left white matter | 0.20 (0.12-0.28) | <.001 | <.001 |
| Frontal lobe right gray matter | 0.02 (-0.05-0.10) | .52 | .66 |
| Frontal lobe right white matter | 0.16 (0.08-0.25) | <.001 | <.001 |
| Gyri parahippocampalis et ambiens anterior part left gray matter | 0.07 (-0.03-0.17) | .08 | .24 |
| Gyri parahippocampalis et ambiens anterior part left white matter | 0.16 (0.06-0.26) | <.001 | .001 |
| Gyri parahippocampalis et ambiens anterior part right gray matter | 0.07 (-0.03-0.17) | .09 | .25 |
| Gyri parahippocampalis et ambiens anterior part right white matter | 0.09 (-0.01-0.19) | .03 | .13 |
| Gyri parahippocampalis et ambiens posterior part left gray matter | 0.11 (0.02-0.20) | .01 | .06 |
| Gyri parahippocampalis et ambiens posterior part left white matter | 0.07 (-0.04-0.17) | .07 | .20 |

| **Regional volume** | **Standardized β coefficient (95% confidence intervals)** | **Raw *p* value** | **BH corrected *p* value** |
|---|---|---|---|
| Gyri parahippocampalis et ambiens posterior part right gray matter | 0.02 (-0.07-0.12) | .59 | .71 |
| Gyri parahippocampalis et ambiens posterior part right white matter | 0.14 (0.04-0.24) | <.001 | .01 |
| Hippocampus left | -0.08 (-0.17-0.02) | .06 | .20 |
| Hippocampus right | -0.06 (-0.16-0.04) | .16 | .34 |
| Insula left gray matter | 0.04 (-0.04-0.12) | .28 | .48 |
| Insula left white matter | 0.17 (0.08-0.26) | <.001 | <.001 |
| Insula right gray matter | -0.02 (-0.10-0.05) | .53 | .67 |
| Insula right white matter | 0.16 (0.08-0.25) | <.001 | <.001 |
| Lateral occipitotemporal gyrus gyrus fusiformis anterior part left gray matter | 0.20 (0.12-0.29) | <.001 | <.001 |
| Lateral occipitotemporal gyrus gyrus fusiformis anterior part left white matter | -0.08 (-0.18-0.02) | .06 | .20 |
| Lateral occipitotemporal gyrus gyrus fusiformis anterior part right gray matter | 0.15 (0.07-0.24) | <.001 | .001 |
| Lateral occipitotemporal gyrus gyrus fusiformis anterior part right white matter | -0.04 (-0.14-0.06) | .38 | .54 |
| Lateral occipitotemporal gyrus gyrus fusiformis posterior part left gray matter | 0.11 (0.02-0.20) | .01 | .06 |
| Lateral occipitotemporal gyrus gyrus fusiformis posterior part left white matter | -0.04 (-0.14-0.05) | .29 | .49 |
| Lateral occipitotemporal gyrus gyrus fusiformis posterior part right gray matter | 0.09 (-0.01-0.18) | .03 | .13 |
| Lateral occipitotemporal gyrus gyrus fusiformis posterior part right white matter | 0.005 (-0.10-0.11) | .89 | .92 |
| Lateral ventricle left | -0.12 (-0.23- -0.02) | <.001 | .001 |
| Lateral ventricle right | -0.10 (-0.20-0.004) | <.001 | .002 |
| Lentiform nucleus left | -0.07 (-0.15-0.02) | .10 | .25 |
| Lentiform nucleus right | -0.10 (-0.18- -0.02) | .02 | .09 |
| Medial and inferior temporal gyri anterior part left gray matter | 0.22 (0.15-0.29) | <.001 | <.001 |
| Medial and inferior temporal gyri anterior part left white matter | 0.04 (-0.05-0.13) | .31 | .50 |
| Medial and inferior temporal gyri anterior part right gray matter | 0.22 (0.16-0.29) | <.001 | <.001 |
| Medial and inferior temporal gyri anterior part right white matter | 0.01 (-0.09-0.11) | .78 | .86 |
| Medial and inferior temporal gyri posterior part left gray matter | 0.11 (0.05-0.18) | <.001 | .005 |
| Medial and inferior temporal gyri posterior part left white matter | -0.06 (-0.16-0.04) | .17 | .36 |
| Medial and inferior temporal gyri posterior part right gray matter | 0.14 (0.07-0.20) | <.001 | <.001 |
| Medial and inferior temporal gyri posterior part right white matter | -0.04 (-0.14-0.06) | .30 | .50 |
| Occipital lobe left gray matter | -0.02 (-0.09-0.04) | .48 | .64 |
| Occipital lobe left white matter | -0.08 (-0.18-0.03) | .09 | .24 |
| Occipital lobe right gray matter | 0.01 (-0.07-0.08) | .85 | .92 |
| Occipital lobe right white matter | -0.06 (-0.16-0.04) | .16 | .34 |
| Parietal lobe left gray matter | 0.07 (0.01-0.12) | .02 | .09 |
| Parietal lobe left white matter | 0.11 (0.02-0.20) | .01 | .06 |
| Parietal lobe right gray matter | 0.09 (0.03-0.15) | .001 | .01 |

| Regional volume | Standardized β coefficient (95% confidence intervals) | Raw *p* value | BH corrected *p* value |
|---|---|---|---|
| Parietal lobe right white matter | 0.07 (-0.03-0.16) | .11 | .26 |
| Subthalamic nucleus left | -0.13 (-0.23- -0.03) | .003 | .03 |
| Subthalamic nucleus right | -0.10 (-0.19-0.002) | .01 | .06 |
| Superior temporal gyrus middle part left gray matter | 0.08 (-0.0002-0.16) | .04 | .14 |
| Superior temporal gyrus middle part left white matter | 0.06 (-0.04-0.16) | .13 | .29 |
| Superior temporal gyrus middle part right gray matter | 0.11 (0.04-0.18) | .001 | .01 |
| Superior temporal gyrus middle part right white matter | 0.05 (-0.05-0.14) | .27 | .47 |
| Superior temporal gyrus posterior part left gray matter | 0.06 (-0.02-0.14) | .10 | .25 |
| Superior temporal gyrus posterior part left white matter | 0.03 (-0.07-0.13) | .36 | .52 |
| Superior temporal gyrus posterior part right gray matter | 0.04 (-0.04-0.12) | .26 | .47 |
| Superior temporal gyrus posterior part right white matter | 0.04 (-0.06-0.13) | .35 | .52 |
| Thalamus left high intensity part in T2 | 0.005 (-0.07-0.08) | .91 | .92 |
| Thalamus left low intensity part in T2 | 0.01 (-0.10-0.12) | .27 | .47 |
| Thalamus right high intensity part in T2 | -0.03 (-0.12-0.05) | .39 | .55 |
| Thalamus right low intensity part in T2 | 0.02 (-0.08-0.12) | .67 | .79 |
| *Interaction* | | | |
| Cingulate gyrus anterior part left white matter | 0.03 (-0.06-0.12) | .68 | .79 |
| Gyri parahippocampalis et ambiens anterior part left white matter | 0.04 (-0.05-0.12) | .03 | .12 |
| Lateral occipitotemporal gyrus gyrus fusiformis anterior part right white matter | 0.04 (-0.05-0.12) | .04 | .16 |
| Lentiform nucleus right | -0.06 (-0.13-0.01) | .03 | .12 |
| Medial and inferior temporal gyri anterior part right white matter | -0.04 (-0.12-0.05) | .02 | .10 |
| Occipital lobe left gray matter | -0.06 (-0.12- -0.001) | .01 | .08 |
| *SIMD* | | | |
| Amygdala left | 0.05 (-0.07-0.18) | .36 | .52 |
| Amygdala right | -0.06 (-0.18-0.07) | .33 | .51 |
| Anterior temporal lobe lateral part left gray matter | -0.004 (-0.11-0.10) | .93 | .94 |
| Anterior temporal lobe lateral part left white matter | 0.04 (-0.09-0.17) | .46 | .62 |
| Anterior temporal lobe lateral part right gray matter | 0.05 (-0.06-0.15) | .38 | .54 |
| Anterior temporal lobe lateral part right white matter | -0.01 (-0.14-0.12) | .89 | .92 |
| Anterior temporal lobe medial part left gray matter | 0.02 (-0.10-0.14) | .77 | .85 |
| Anterior temporal lobe medial part left white matter | 0.02 (-0.12-0.16) | .63 | .75 |
| Anterior temporal lobe medial part right gray matter | 0.01 (-0.11-0.13) | .91 | .92 |
| Anterior temporal lobe medial part right white matter | 0.07 (-0.06-0.20) | .22 | .44 |
| Brainstem | 0.12 (0.003-0.23) | .03 | .13 |
| Caudate nucleus left | 0.08 (-0.03-0.20) | .10 | .26 |
| Caudate nucleus right | 0.10 (-0.02-0.22) | .06 | .20 |
| Cerebellum left | 0.04 (-0.05-0.14) | .34 | .52 |
| Cerebellum right | 0.06 (-0.03-0.15) | .19 | .41 |

| Regional volume | Standardized β coefficient (95% confidence intervals) | Raw *p* value | BH corrected *p* value |
|---|---|---|---|
| Cerebrospinal fluid | 0.01 (-0.12-0.15) | .52 | .66 |
| Cingulate gyrus anterior part left gray matter | 0.05 (-0.07-0.18) | .31 | .50 |
| Cingulate gyrus anterior part left white matter | 0.02 (-0.32-0.35) | .02 | .09 |
| Cingulate gyrus anterior part right gray matter | -0.03 (-0.16-0.10) | .59 | .71 |
| Cingulate gyrus anterior part right white matter | 0.05 (-0.08-0.18) | .31 | .50 |
| Cingulate gyrus posterior part left gray matter | 0.01 (-0.10-0.12) | .89 | .92 |
| Cingulate gyrus posterior part left white matter | 0.11 (-0.005-0.22) | .04 | .14 |
| Cingulate gyrus posterior part right gray matter | 0.02 (-0.10-0.14) | .75 | .85 |
| Cingulate gyrus posterior part right white matter | 0.10 (-0.02-0.21) | .07 | .20 |
| Corpus callosum | 0.12 (-0.001-0.24) | .03 | .12 |
| Frontal lobe left gray matter | 0.07 (-0.02-0.17) | .12 | .29 |
| Frontal lobe left white matter | 0.06 (-0.05-0.16) | .26 | .47 |
| Frontal lobe right gray matter | 0.03 (-0.07-0.13) | .55 | .69 |
| Frontal lobe right white matter | 0.12 (0.01-0.23) | .02 | .10 |
| Gyri parahippocampalis et ambiens anterior part left gray matter | 0.03 (-0.11-0.16) | .58 | .71 |
| Gyri parahippocampalis et ambiens anterior part left white matter | -0.10 (-0.43-0.23) | .11 | .26 |
| Gyri parahippocampalis et ambiens anterior part right gray matter | -0.02 (-0.15-0.11) | .74 | .84 |
| Gyri parahippocampalis et ambiens anterior part right white matter | 0.03 (-0.09-0.16) | .55 | .69 |
| Gyri parahippocampalis et ambiens posterior part left gray matter | 0.13 (0.01-0.26) | .02 | .10 |
| Gyri parahippocampalis et ambiens posterior part left white matter | 0.06 (-0.08-0.20) | .23 | .45 |
| Gyri parahippocampalis et ambiens posterior part right gray matter | 0.06 (-0.07-0.19) | .32 | .51 |
| Gyri parahippocampalis et ambiens posterior part right white matter | -0.02 (-0.15-0.11) | .71 | .81 |
| Hippocampus left | 0.06 (-0.07-0.02) | .31 | .50 |
| Hippocampus right | 0.03 (-0.10-0.16) | .59 | .71 |
| Insula left gray matter | -0.04 (-0.14-0.07) | .48 | .64 |
| Insula left white matter | 0.12 (0.01-0.24) | .02 | .10 |
| Insula right gray matter | -0.001 (-0.10-0.10) | .98 | .98 |
| Insula right white matter | 0.09 (-0.02-0.21) | .07 | .21 |
| Lateral occipitotemporal gyrus gyrus fusiformis anterior part left gray matter | 0.13 (0.01-0.25) | .02 | .09 |
| Lateral occipitotemporal gyrus gyrus fusiformis anterior part left white matter | 0.09 (-0.03-0.22) | .10 | .25 |
| Lateral occipitotemporal gyrus gyrus fusiformis anterior part right gray matter | 0.07 (-0.04-0.18) | .17 | .36 |
| Lateral occipitotemporal gyrus gyrus fusiformis anterior part right white matter | 0.01 (-0.32-0.34) | .87 | .92 |
| Lateral occipitotemporal gyrus gyrus fusiformis posterior part left gray matter | 0.11 (-0.01-0.23) | .04 | .15 |
| Lateral occipitotemporal gyrus gyrus fusiformis posterior part left white matter | 0.06 (-0.06-0.19) | .26 | .47 |
| Lateral occipitotemporal gyrus gyrus fusiformis posterior part right gray matter | 0.05 (-0.07-0.17) | .39 | .54 |

| Regional volume | Standardized β coefficient (95% confidence intervals) | Raw *p* value | BH corrected *p* value |
|---|---|---|---|
| Lateral occipitotemporal gyrus gyrus fusiformis posterior part right white matter | 0.05 (-0.09-0.18) | .35 | .52 |
| Lateral ventricle left | 0.08 (-0.06-0.22) | .051 | .17 |
| Lateral ventricle right | 0.06 (-0.08-0.19) | .11 | .26 |
| Lentiform nucleus left | -0.08 (-0.19-0.03) | .13 | .29 |
| Lentiform nucleus right | 0.10 (-0.17-0.37) | .33 | .52 |
| Medial and inferior temporal gyri anterior part left gray matter | 0.04 (-0.05-0.14) | .34 | .52 |
| Medial and inferior temporal gyri anterior part left white matter | 0.07 (-0.05-0.19) | .22 | .44 |
| Medial and inferior temporal gyri anterior part right gray matter | 0.03 (-0.05-0.11) | .49 | .65 |
| Medial and inferior temporal gyri anterior part right white matter | 0.17 (-0.16-0.50) | .004 | .03 |
| Medial and inferior temporal gyri posterior part left gray matter | 0.02 (-0.07-0.10) | .73 | .82 |
| Medial and inferior temporal gyri posterior part left white matter | -0.01 (-0.14-0.12) | .88 | .92 |
| Medial and inferior temporal gyri posterior part right gray matter | 0.005 (-0.08-0.09) | .91 | .92 |
| Medial and inferior temporal gyri posterior part right white matter | 0.08 (-0.05-0.21) | .13 | .29 |
| Occipital lobe left gray matter | 0.20 (-0.02-0.42) | .03 | .12 |
| Occipital lobe left white matter | 0.05 (-0.08-0.19) | .36 | .53 |
| Occipital lobe right gray matter | 0.02 (-0.07-0.11) | .65 | .76 |
| Occipital lobe right white matter | 0.07 (-0.06-0.20) | .23 | .45 |
| Parietal lobe left gray matter | 0.01 (-0.06-0.09) | .69 | .80 |
| Parietal lobe left white matter | 0.06 (-0.06-0.18) | .26 | .47 |
| Parietal lobe right gray matter | 0.01 (-0.06-0.09) | .76 | .85 |
| Parietal lobe right white matter | 0.06 (-0.06-0.17) | .29 | .49 |
| Subthalamic nucleus left | -0.03 (-0.17-0.10) | .56 | .69 |
| Subthalamic nucleus right | 0.06 (-0.07-0.18) | .25 | .47 |
| Superior temporal gyrus middle part left gray matter | 0.01 (-0.10-0.11) | .91 | .92 |
| Superior temporal gyrus middle part left white matter | -0.04 (-0.16-0.09) | .50 | .65 |
| Superior temporal gyrus middle part right gray matter | -0.01 (-0.10-0.09) | .90 | .92 |
| Superior temporal gyrus middle part right white matter | 0.07 (-0.06-0.19) | .21 | .42 |
| Superior temporal gyrus posterior part left gray matter | 0.05 (-0.05-0.16) | .28 | .48 |
| Superior temporal gyrus posterior part left white matter | 0.04 (-0.09-0.17) | .43 | .59 |
| Superior temporal gyrus posterior part right gray matter | 0.05 (-0.06-0.15) | .35 | .52 |
| Superior temporal gyrus posterior part right white matter | 0.01 (-0.11-0.14) | .80 | .87 |
| Thalamus left high intensity part in T2 | -0.11 (-0.21- -0.001) | .04 | .14 |
| Thalamus left low intensity part in T2 | 0.02 (-0.13-0.16) | .41 | .57 |
| Thalamus right high intensity part in T2 | -0.09 (-0.20-0.02) | .09 | .25 |
| Thalamus right low intensity part in T2 | -0.04 (-0.17-0.09) | .47 | .63 |

Fully adjusted ridge regression model, including gestation at birth, SIMD, gestation at MRI, the interaction term (where significant), birth weight z-score, birth head circumference z-score, sex, smoking in pregnancy, and breast milk at discharge. Corrected for false discovery rate.
BH = Benjamini-Hochberg correction, SIMD = Scottish Index of Multiple Deprivation.

**eTable 4. Regional brain volumes – relationship with gestational age, the Scottish Index of Multiple Deprivation and interaction effect (baseline unadjusted linear regression model)**

| Regional volume | Standardized β coefficient (95% confidence intervals) | Raw *p* value | BH corrected *p* value |
|---|---|---|---|
| *Gestation* | | | |
| Amygdala left | 0.12 (0.03-0.21) | .01 | .03 |
| Amygdala right | 0.07 (-0.03-0.16) | .16 | .28 |
| Anterior temporal lobe lateral part left gray matter | 0.19 (0.11-0.26) | <.001 | <.001 |
| Anterior temporal lobe lateral part left white matter | 0.15 (0.07-0.24) | <.001 | .003 |
| Anterior temporal lobe lateral part right gray matter | 0.13 (0.05-0.20) | .002 | .01 |
| Anterior temporal lobe lateral part right white matter | 0.16 (0.07-0.25) | <.001 | .002 |
| Anterior temporal lobe medial part left gray matter | 0.10 (0.02-0.19) | .02 | .051 |
| Anterior temporal lobe medial part left white matter | 0.11 (0.01-0.20) | .03 | .08 |
| Anterior temporal lobe medial part right gray matter | 0.10 (0.02-0.19) | .02 | .05 |
| Anterior temporal lobe medial part right white matter | 0.12 (0.03-0.22) | .01 | .03 |
| Brainstem | 0.06 (-0.02-0.15) | .13 | .24 |
| Caudate nucleus left | 0.26 (0.18-0.34) | <.001 | <.001 |
| Caudate nucleus right | 0.23 (0.15-0.32) | <.001 | <.001 |
| Cerebellum left | 0.07 (0.002-0.14) | .045 | .10 |
| Cerebellum right | 0.04 (-0.02-0.11) | .19 | .32 |
| Cerebrospinal fluid | -0.33 (-0.42- -0.24) | <.001 | <.001 |
| Cingulate gyrus anterior part left gray matter | 0.05 (-0.04-0.15) | .27 | .40 |
| Cingulate gyrus anterior part left white matter | 0.17 (0.08-0.27) | <.001 | .002 |
| Cingulate gyrus anterior part right gray matter | -0.03 (-0.12-0.07) | .60 | .71 |
| Cingulate gyrus anterior part right white matter | 0.13 (0.04-0.23) | .01 | .03 |
| Cingulate gyrus posterior part left gray matter | 0.01 (-0.08-0.09) | .88 | .91 |
| Cingulate gyrus posterior part left white matter | 0.17 (0.09-0.26) | <.001 | <.001 |
| Cingulate gyrus posterior part right gray matter | -0.04 (-0.13-0.05) | .35 | .47 |
| Cingulate gyrus posterior part right white matter | 0.20 (0.11-0.28) | <.001 | <.001 |
| Corpus callosum | 0.16 (0.07-0.24) | <.001 | .002 |
| Frontal lobe left gray matter | 0.09 (0.01-0.16) | .03 | .07 |
| Frontal lobe left white matter | 0.34 (0.25-0.42) | <.001 | <.001 |
| Frontal lobe right gray matter | 0.09 (0.01-0.17) | .02 | .06 |
| Frontal lobe right white matter | 0.30 (0.21-0.38) | <.001 | <.001 |
| Gyri parahippocampalis et ambiens anterior part left gray matter | 0.11 (0.02-0.21) | .02 | .0499 |
| Gyri parahippocampalis et ambiens anterior part left white matter | 0.23 (0.14-0.31) | <.001 | <.001 |
| Gyri parahippocampalis et ambiens anterior part right gray matter | 0.13 (0.04-0.22) | .01 | .02 |
| Gyri parahippocampalis et ambiens anterior part right white matter | 0.14 (0.05-0.23) | .002 | .01 |
| Gyri parahippocampalis et ambiens posterior part left gray matter | 0.13 (0.04-0.21) | .003 | .01 |
| Gyri parahippocampalis et ambiens posterior part left white matter | 0.12 (0.03-0.22) | .01 | .04 |

| Regional volume | Standardized β coefficient (95% confidence intervals) | Raw *p* value | BH corrected *p* value |
|---|---|---|---|
| Gyri parahippocampalis et ambiens posterior part right gray matter | 0.06 (-0.03-0.15) | .18 | .30 |
| Gyri parahippocampalis et ambiens posterior part right white matter | 0.23 (0.14-0.32) | .002 | .01 |
| Hippocampus left | -0.02 (-0.11-0.07) | .67 | .75 |
| Hippocampus right | -0.002 (-0.10-0.09) | .97 | .98 |
| Insula left gray matter | 0.13 (0.05-0.21) | .001 | .01 |
| Insula left white matter | 0.29 (0.20-0.38) | <.001 | <.001 |
| Insula right gray matter | 0.07 (-0.01-0.15) | .08 | .16 |
| Insula right white matter | 0.29 (0.20-0.38) | <.001 | <.001 |
| Lateral occipitotemporal gyrus gyrus fusiformis anterior part left gray matter | 0.25 (0.17-0.34) | <.001 | <.001 |
| Lateral occipitotemporal gyrus gyrus fusiformis anterior part left white matter | -0.02 (-0.11-0.07) | .72 | .80 |
| Lateral occipitotemporal gyrus gyrus fusiformis anterior part right gray matter | 0.22 (0.14-0.30) | <.001 | <.001 |
| Lateral occipitotemporal gyrus gyrus fusiformis anterior part right white matter | 0.04 (-0.05-0.13) | .37 | .49 |
| Lateral occipitotemporal gyrus gyrus fusiformis posterior part left gray matter | 0.14 (0.05-0.22) | .001 | .01 |
| Lateral occipitotemporal gyrus gyrus fusiformis posterior part left white matter | 0.02 (-0.07-0.12) | .62 | .72 |
| Lateral occipitotemporal gyrus gyrus fusiformis posterior part right gray matter | 0.14 (0.05-0.22) | .001 | .01 |
| Lateral occipitotemporal gyrus gyrus fusiformis posterior part right white matter | 0.07 (-0.03-0.16) | .17 | .30 |
| Lateral ventricle left | -0.22 (-0.31- -0.12) | <.001 | <.001 |
| Lateral ventricle right | -0.20 (-0.29- -0.10) | <.001 | <.001 |
| Lentiform nucleus left | -0.01 (-0.09-0.07) | .79 | .85 |
| Lentiform nucleus right | -0.04 (-0.12-0.03) | .28 | .40 |
| Medial and inferior temporal gyri anterior part left gray matter | 0.29 (0.22-0.35) | <.001 | <.001 |
| Medial and inferior temporal gyri anterior part left white matter | 0.16 (0.07-0.25) | <.001 | .003 |
| Medial and inferior temporal gyri anterior part right gray matter | 0.28 (0.23-0.34) | <.001 | <.001 |
| Medial and inferior temporal gyri anterior part right white matter | 0.11 (0.01-0.20) | .02 | .07 |
| Medial and inferior temporal gyri posterior part left gray matter | 0.18 (0.12-0.24) | <.001 | <.001 |
| Medial and inferior temporal gyri posterior part left white matter | 0.04 (-0.06-0.13) | .46 | .59 |
| Medial and inferior temporal gyri posterior part right gray matter | 0.20 (0.14-0.26) | <.001 | <.001 |
| Medial and inferior temporal gyri posterior part right white matter | 0.04 (-0.06-0.13) | .42 | .55 |
| Occipital lobe left gray matter | 0.05 (-0.01-0.11) | .12 | .23 |
| Occipital lobe left white matter | -0.003 (-0.10-0.09) | .95 | .96 |
| Occipital lobe right gray matter | 0.08 (0.01-0.15) | .02 | .06 |
| Occipital lobe right white matter | 0.03 (-0.07-0.12) | .60 | .71 |
| Parietal lobe left gray matter | 0.17 (0.11-0.23) | <.001 | <.001 |
| Parietal lobe left white matter | 0.24 (0.14-0.33) | <.001 | <.001 |
| Parietal lobe right gray matter | 0.18 (0.12-0.24) | <.001 | <.001 |

| **Regional volume** | **Standardized β coefficient (95% confidence intervals)** | **Raw *p* value** | **BH corrected *p* value** |
|---|---|---|---|
| Parietal lobe right white matter | 0.20 (0.10-0.29) | <.001 | <.001 |
| Subthalamic nucleus left | -0.12 (-0.21- -0.02) | .01 | .04 |
| Subthalamic nucleus right | -0.09 (-0.18-0.003) | .06 | .13 |
| Superior temporal gyrus middle part left gray matter | 0.16 (0.08-0.24) | <.001 | <.001 |
| Superior temporal gyrus middle part left white matter | 0.17 (0.08-0.26) | <.001 | .003 |
| Superior temporal gyrus middle part right gray matter | 0.18 (0.11-0.25) | <.001 | <.001 |
| Superior temporal gyrus middle part right white matter | 0.14 (0.04-0.23) | .004 | .02 |
| Superior temporal gyrus posterior part left gray matter | 0.14 (0.06-0.21) | <.001 | .003 |
| Superior temporal gyrus posterior part left white matter | 0.13 (0.03-0.22) | .01 | .03 |
| Superior temporal gyrus posterior part right gray matter | 0.13 (0.05-0.20) | .001 | .01 |
| Superior temporal gyrus posterior part right white matter | 0.15 (0.05-0.25) | .002 | .01 |
| Thalamus left high intensity part in T2 | 0.09 (0.01-0.16) | .02 | .06 |
| Thalamus left low intensity part in T2 | 0.06 (-0.04-0.15) | .24 | .37 |
| Thalamus right high intensity part in T2 | 0.05 (-0.03-0.13) | .24 | .37 |
| Thalamus right low intensity part in T2 | 0.03 (-0.06-0.11) | .56 | .68 |
| *Interaction* | | | |
| Cerebrospinal fluid | -0.11 (-0.20- -0.03) | .01 | .03 |
| Corpus callosum | -0.09 (-0.17- -0.01) | .03 | .08 |
| Gyri parahippocampalis et ambiens anterior part left white matter | 0.09 (0.01-0.17) | .04 | .09 |
| Lateral ventricle left | -0.09 (-0.18- -0.0002) | .0496 | .11 |
| Medial and inferior temporal gyri anterior part right white matter | -0.11 (-0.19- -0.02) | .02 | .050 |
| Occipital lobe left gray matter | -0.06 (-0.12- -0.002) | .04 | .10 |
| *SIMD* | | | |
| Amygdala left | 0.04 (-0.07-0.16) | .47 | .59 |
| Amygdala right | -0.06 (-0.18-0.06) | .33 | .45 |
| Anterior temporal lobe lateral part left gray matter | -0.01 (-0.11-0.08) | .78 | .83 |
| Anterior temporal lobe lateral part left white matter | 0.05 (-0.06-0.16) | .37 | .49 |
| Anterior temporal lobe lateral part right gray matter | 0.02 (-0.08-0.12) | .69 | .77 |
| Anterior temporal lobe lateral part right white matter | -0.03 (-0.14-0.09) | .61 | .71 |
| Anterior temporal lobe medial part left gray matter | 0.00002 (-0.11-0.11) | .9997 | .9997 |
| Anterior temporal lobe medial part left white matter | 0.03 (-0.10-0.15) | .67 | .75 |
| Anterior temporal lobe medial part right gray matter | -0.02 (-0.13-0.09) | .76 | .83 |
| Anterior temporal lobe medial part right white matter | 0.07 (-0.05-0.19) | .28 | .40 |
| Brainstem | 0.11 (0.004-0.22) | .04 | .10 |
| Caudate nucleus left | 0.11 (0.002-0.21) | .04 | .10 |
| Caudate nucleus right | 0.12 (0.01-0.23) | .04 | .10 |
| Cerebellum left | 0.06 (-0.03-0.15) | .18 | .30 |
| Cerebellum right | 0.08 (-0.01-0.16) | .09 | .18 |
| Cerebrospinal fluid | 0.46 (0.16-0.76) | .003 | .01 |

| Regional volume | Standardized β coefficient (95% confidence intervals) | Raw *p* value | BH corrected *p* value |
|---|---|---|---|
| Cingulate gyrus anterior part left gray matter | 0.001 (-0.12-0.12) | .99 | .999 |
| Cingulate gyrus anterior part left white matter | 0.08 (-0.05-0.20) | .23 | .37 |
| Cingulate gyrus anterior part right gray matter | -0.05 (-0.17-0.07) | .39 | .51 |
| Cingulate gyrus anterior part right white matter | 0.06 (-0.06-0.18) | .35 | .47 |
| Cingulate gyrus posterior part left gray matter | 0.02 (-0.08-0.13) | .65 | .75 |
| Cingulate gyrus posterior part left white matter | 0.17 (0.06-0.28) | .003 | .01 |
| Cingulate gyrus posterior part right gray matter | 0.01 (-0.10-0.13) | .83 | .87 |
| Cingulate gyrus posterior part right white matter | 0.11 (0.001-0.23) | .047 | .11 |
| Corpus callosum | 0.43 (0.14-0.73) | .004 | .02 |
| Frontal lobe left gray matter | 0.03 (-0.07-0.13) | .53 | .65 |
| Frontal lobe left white matter | 0.07 (-0.03-0.18) | .18 | .31 |
| Frontal lobe right gray matter | -0.02 (-0.11-0.08) | .75 | .82 |
| Frontal lobe right white matter | 0.13 (0.02-0.24) | .02 | .05 |
| Gyri parahippocampalis et ambiens anterior part left gray matter | 0.02 (-0.10-0.14) | .75 | .82 |
| Gyri parahippocampalis et ambiens anterior part left white matter | -0.28 (-0.58-0.02) | .06 | .13 |
| Gyri parahippocampalis et ambiens anterior part right gray matter | -0.04 (-0.16-0.08) | .46 | .59 |
| Gyri parahippocampalis et ambiens anterior part right white matter | 0.06 (-0.06-0.17) | .32 | .45 |
| Gyri parahippocampalis et ambiens posterior part left gray matter | 0.10 (-0.01-0.21) | .06 | .13 |
| Gyri parahippocampalis et ambiens posterior part left white matter | 0.07 (-0.05-0.19) | .25 | .39 |
| Gyri parahippocampalis et ambiens posterior part right gray matter | 0.03 (-0.08-0.14) | .61 | .71 |
| Gyri parahippocampalis et ambiens posterior part right white matter | 0.01 (-0.11-0.12) | .93 | .95 |
| Hippocampus left | 0.08 (-0.04-0.20) | .20 | .33 |
| Hippocampus right | 0.06 (-0.06-0.19) | .30 | .43 |
| Insula left gray matter | -0.03 (-0.13-0.08) | .60 | .71 |
| Insula left white matter | 0.13 (0.02-0.25) | .02 | .07 |
| Insula right gray matter | -0.01 (-0.11-0.09) | .86 | .89 |
| Insula right white matter | 0.12 (0.01-0.23) | .04 | .10 |
| Lateral occipitotemporal gyrus gyrus fusiformis anterior part left gray matter | 0.09 (-0.02-0.19) | .11 | .21 |
| Lateral occipitotemporal gyrus gyrus fusiformis anterior part left white matter | 0.12 (0.0003-0.24) | .049 | .11 |
| Lateral occipitotemporal gyrus gyrus fusiformis anterior part right gray matter | 0.05 (-0.05-0.16) | .30 | .42 |
| Lateral occipitotemporal gyrus gyrus fusiformis anterior part right white matter | 0.14 (0.02-0.26) | .02 | .06 |
| Lateral occipitotemporal gyrus gyrus fusiformis posterior part left gray matter | 0.08 (-0.03-0.18) | .15 | .28 |
| Lateral occipitotemporal gyrus gyrus fusiformis posterior part left white matter | 0.07 (-0.05-0.19) | .24 | .37 |
| Lateral occipitotemporal gyrus gyrus fusiformis posterior part right gray matter | 0.06 (-0.05-0.17) | .27 | .40 |
| Lateral occipitotemporal gyrus gyrus fusiformis posterior part right white matter | 0.09 (-0.03-0.21) | .15 | .28 |

| **Regional volume** | **Standardized β coefficient (95% confidence intervals)** | **Raw *p* value** | **BH corrected *p* value** |
|---|---|---|---|
| Lateral ventricle left | 0.44 (0.11-0.76) | .01 | .03 |
| Lateral ventricle right | 0.11 (-0.01-0.23) | .09 | .17 |
| Lentiform nucleus left | -0.07 (-0.17-0.03) | .19 | .31 |
| Lentiform nucleus right | -0.10 (-0.20- -0.01) | .04 | .09 |
| Medial and inferior temporal gyri anterior part left gray matter | 0.06 (-0.02-0.15) | .14 | .25 |
| Medial and inferior temporal gyri anterior part left white matter | 0.10 (-0.02-0.21) | .11 | .21 |
| Medial and inferior temporal gyri anterior part right gray matter | 0.04 (-0.03-0.12) | .27 | .40 |
| Medial and inferior temporal gyri anterior part right white matter | 0.47 (0.14-0.79) | .005 | .02 |
| Medial and inferior temporal gyri posterior part left gray matter | 0.03 (-0.05-0.11) | .42 | .55 |
| Medial and inferior temporal gyri posterior part left white matter | 0.03 (-0.10-0.15) | .66 | .75 |
| Medial and inferior temporal gyri posterior part right gray matter | 0.03 (-0.05-0.10) | .49 | .61 |
| Medial and inferior temporal gyri posterior part right white matter | 0.10 (-0.02-0.22) | .11 | .21 |
| Occipital lobe left gray matter | 0.21 (-0.01-0.43) | .06 | .13 |
| Occipital lobe left white matter | 0.04 (-0.09-0.17) | .53 | .65 |
| Occipital lobe right gray matter | 0.01 (-0.08-0.10) | .82 | .86 |
| Occipital lobe right white matter | 0.05 (-0.08-0.17) | .46 | .59 |
| Parietal lobe left gray matter | 0.02 (-0.05-0.10) | .53 | .65 |
| Parietal lobe left white matter | 0.07 (-0.05-0.19) | .23 | .37 |
| Parietal lobe right gray matter | 0.02 (-0.06-0.09) | .66 | .75 |
| Parietal lobe right white matter | 0.07 (-0.05-0.19) | .24 | .37 |
| Subthalamic nucleus left | -0.07 (-0.19-0.05) | .27 | .40 |
| Subthalamic nucleus right | 0.04 (-0.07-0.16) | .49 | .61 |
| Superior temporal gyrus middle part left gray matter | 0.02 (-0.08-0.12) | .73 | .80 |
| Superior temporal gyrus middle part left white matter | 0.02 (-0.11-0.14) | .80 | .85 |
| Superior temporal gyrus middle part right gray matter | 0.01 (-0.08-0.09) | .85 | .88 |
| Superior temporal gyrus middle part right white matter | 0.10 (-0.02-0.22) | .11 | .21 |
| Superior temporal gyrus posterior part left gray matter | 0.06 (-0.04-0.16) | .23 | .37 |
| Superior temporal gyrus posterior part left white matter | 0.08 (-0.04-0.21) | .19 | .32 |
| Superior temporal gyrus posterior part right gray matter | 0.07 (-0.03-0.17) | .15 | .28 |
| Superior temporal gyrus posterior part right white matter | 0.06 (-0.07-0.18) | .36 | .49 |
| Thalamus left high intensity part in T2 | -0.09 (-0.19-0.004) | .06 | .13 |
| Thalamus left low intensity part in T2 | 0.04 (-0.09-0.16) | .58 | .70 |
| Thalamus right high intensity part in T2 | -0.07 (-0.17-0.03) | .16 | .28 |
| Thalamus right low intensity part in T2 | -0.06 (-0.17-0.06) | .31 | .44 |

Baseline unadjusted linear regression model, including gestation at birth, SIMD, gestation at MRI, and the interaction term (where significant). Corrected for false discovery rate.
BH = Benjamini-Hochberg correction, SIMD = Scottish Index of Multiple Deprivation.

**eTable 5: Cortical measures – relationship with gestational age and the Scottish Index of Multiple Deprivation (baseline unadjusted linear regression model)**

| Cortical measure | Standardized β coefficient (95% confidence intervals) | Raw *p* value | BH corrected *p* value |
|---|---|---|---|
| *Gestational age* | | | |
| Mean cortical curvature | -0.02 (-0.09-0.05) | .57 | .57 |
| Mean cortical surface area | 0.19 (0.11-0.26) | <.001 | <.001 |
| Mean cortical thickness | -0.07 (-0.16-0.02) | .15 | .36 |
| Mean gyrification index | 0.18 (0.10-0.26) | <.001 | <.001 |
| Mean sulcal depth | 0.08 (-0.02-0.18) | .11 | .36 |
| *SIMD* | | | |
| Mean cortical curvature | -0.06 (-0.15-0.04) | .22 | .36 |
| Mean cortical surface area | 0.05 (-0.05-0.15) | .29 | .42 |
| Mean cortical thickness | -0.08 (-0.19-0.04) | .37 | .46 |
| Mean gyrification index | 0.05 (-0.06-0.15) | .22 | .36 |
| Mean sulcal depth | 0.05 (-0.08-0.17) | .46 | .51 |

Baseline unadjusted linear regression model, including gestation at birth, SIMD, gestation at MRI, and the interaction term (where significant – none were, so not included). Corrected for false discovery rate.
BH = Benjamini-Hochberg correction, SIMD = Scottish Index of Multiple Deprivation.

**eTable 6: Regional brain volumes – relationship with gestational age, maternal final educational qualification, and interaction effect (fully adjusted ridge regression model)**

| Regional volume | Standardized β coefficient (95% confidence intervals) | Raw *p* value | BH corrected *p* value |
|---|---|---|---|
| *Gestation* | | | |
| Amygdala left | 0.03 (-0.29-0.34) | .53 | .66 |
| Amygdala right | 0.01 (-0.08-0.11) | .74 | .83 |
| Anterior temporal lobe lateral part left gray matter | 0.16 (0.08-0.24) | <.001 | .002 |
| Anterior temporal lobe lateral part left white matter | 0.11 (0.01-0.21) | .01 | .07 |
| Anterior temporal lobe lateral part right gray matter | 0.07 (-0.02-0.15) | .10 | .26 |
| Anterior temporal lobe lateral part right white matter | 0.10 (0.0003-0.21) | .02 | .09 |
| Anterior temporal lobe medial part left gray matter | 0.04 (-0.06-0.13) | .39 | .58 |
| Anterior temporal lobe medial part left white matter | 0.03 (-0.08-0.14) | .46 | .61 |
| Anterior temporal lobe medial part right gray matter | 0.04 (-0.06-0.14) | .37 | .56 |
| Anterior temporal lobe medial part right white matter | 0.04 (-0.06-0.15) | .35 | .54 |
| Brainstem | -0.02 (-0.11-0.07) | .63 | .73 |
| Caudate nucleus left | 0.07 (-0.21-0.36) | .08 | .22 |
| Caudate nucleus right | 0.07 (-0.24-0.37) | .08 | .23 |
| Cerebellum left | -0.04 (-0.11-0.03) | .23 | .41 |
| Cerebellum right | -0.06 (-0.12-0.01) | .10 | .25 |
| Cerebrospinal fluid | -0.15 (-0.46-0.16) | <.001 | <.001 |
| Cingulate gyrus anterior part left gray matter | 0.03 (-0.07-0.12) | .53 | .66 |
| Cingulate gyrus anterior part left white matter | 0.07 (-0.03-0.18) | .07 | .20 |
| Cingulate gyrus anterior part right gray matter | -0.06 (-0.16-0.04) | .16 | .34 |
| Cingulate gyrus anterior part right white matter | 0.04 (-0.06-0.15) | .28 | .47 |
| Cingulate gyrus posterior part left gray matter | -0.07 (-0.15-0.02) | .11 | .28 |
| Cingulate gyrus posterior part left white matter | 0.05 (-0.03-0.14) | .19 | .36 |
| Cingulate gyrus posterior part right gray matter | -0.10 (-0.20- -0.01) | .02 | .09 |
| Cingulate gyrus posterior part right white matter | 0.08 (-0.01-0.17) | .047 | .16 |
| Corpus callosum | 0.08 (-0.02-0.18) | .06 | .19 |
| Frontal lobe left gray matter | 0.02 (-0.06-0.10) | .61 | .73 |
| Frontal lobe left white matter | 0.20 (0.12-0.29) | <.001 | <.001 |
| Frontal lobe right gray matter | 0.02 (-0.06-0.09) | .62 | .73 |
| Frontal lobe right white matter | 0.17 (0.09-0.26) | <.001 | <.001 |
| Gyri parahippocampalis et ambiens anterior part left gray matter | 0.08 (-0.02-0.19) | .046 | .16 |
| Gyri parahippocampalis et ambiens anterior part left white matter | 0.08 (-0.24-0.40) | .04 | .16 |
| Gyri parahippocampalis et ambiens anterior part right gray matter | 0.09 (-0.01-0.19) | .048 | .16 |
| Gyri parahippocampalis et ambiens anterior part right white matter | 0.03 (-0.29-0.35) | .49 | .63 |
| Gyri parahippocampalis et ambiens posterior part left gray matter | 0.14 (0.04-0.24) | .002 | .02 |
| Gyri parahippocampalis et ambiens posterior part left white matter | 0.04 (-0.31-0.38) | .18 | .36 |

| Regional volume | Standardized β coefficient (95% confidence intervals) | Raw *p* value | BH corrected *p* value |
|---|---|---|---|
| Gyri parahippocampalis et ambiens posterior part right gray matter | 0.05 (-0.05-0.15) | .29 | .47 |
| Gyri parahippocampalis et ambiens posterior part right white matter | 0.13 (0.03-0.24) | .002 | .02 |
| Hippocampus left | -0.08 (-0.18-0.02) | .06 | .18 |
| Hippocampus right | -0.04 (-0.15-0.07) | .30 | .48 |
| Insula left gray matter | 0.03 (-0.05-0.11) | .44 | .60 |
| Insula left white matter | 0.15 (0.06-0.24) | <.001 | .004 |
| Insula right gray matter | -0.04 (-0.12-0.04) | .32 | .50 |
| Insula right white matter | 0.15 (0.06-0.24) | <.001 | .004 |
| Lateral occipitotemporal gyrus gyrus fusiformis anterior part left gray matter | 0.16 (-0.14-0.46) | <.001 | .004 |
| Lateral occipitotemporal gyrus gyrus fusiformis anterior part left white matter | -0.10 (-0.20- -0.003) | .03 | .11 |
| Lateral occipitotemporal gyrus gyrus fusiformis anterior part right gray matter | 0.09 (-0.19-0.37) | .08 | .22 |
| Lateral occipitotemporal gyrus gyrus fusiformis anterior part right white matter | -0.04 (-0.14-0.06) | .40 | .59 |
| Lateral occipitotemporal gyrus gyrus fusiformis posterior part left gray matter | 0.11 (0.02-0.21) | .01 | .06 |
| Lateral occipitotemporal gyrus gyrus fusiformis posterior part left white matter | -0.07 (-0.17-0.02) | .10 | .26 |
| Lateral occipitotemporal gyrus gyrus fusiformis posterior part right gray matter | 0.03 (-0.29-0.34) | .55 | .68 |
| Lateral occipitotemporal gyrus gyrus fusiformis posterior part right white matter | -0.01 (-0.12-0.09) | .75 | .83 |
| Lateral ventricle left | -0.08 (-0.42-0.25) | .002 | .02 |
| Lateral ventricle right | -0.08 (-0.41-0.24) | <.001 | .01 |
| Lentiform nucleus left | -0.08 (-0.17-0.004) | .047 | .16 |
| Lentiform nucleus right | -0.11 (-0.19- -0.02) | .01 | .06 |
| Medial and inferior temporal gyri anterior part left gray matter | 0.12 (-0.11-0.35) | .03 | .11 |
| Medial and inferior temporal gyri anterior part left white matter | 0.03 (-0.06-0.13) | .43 | .60 |
| Medial and inferior temporal gyri anterior part right gray matter | 0.23 (0.16-0.29) | <.001 | <.001 |
| Medial and inferior temporal gyri anterior part right white matter | 0.06 (-0.26-0.38) | .20 | .38 |
| Medial and inferior temporal gyri posterior part left gray matter | 0.12 (0.06-0.19) | <.001 | .004 |
| Medial and inferior temporal gyri posterior part left white matter | -0.08 (-0.18-0.02) | .06 | .19 |
| Medial and inferior temporal gyri posterior part right gray matter | 0.002 (-0.21-0.22) | .97 | .98 |
| Medial and inferior temporal gyri posterior part right white matter | -0.05 (-0.15-0.05) | .25 | .43 |
| Occipital lobe left gray matter | -0.03 (-0.10-0.04) | .42 | .60 |
| Occipital lobe left white matter | -0.08 (-0.18-0.02) | .08 | .22 |
| Occipital lobe right gray matter | 0.002 (-0.07-0.08) | .97 | .98 |
| Occipital lobe right white matter | -0.07 (-0.17-0.04) | .15 | .33 |
| Parietal lobe left gray matter | 0.06 (0.01-0.12) | .02 | .10 |
| Parietal lobe left white matter | 0.10 (0.003-0.19) | .02 | .10 |
| Parietal lobe right gray matter | 0.08 (0.02-0.14) | .004 | .04 |

| Regional volume | Standardized β coefficient (95% confidence intervals) | Raw *p* value | BH corrected *p* value |
|---|---|---|---|
| Parietal lobe right white matter | 0.06 (-0.04-0.15) | .19 | .37 |
| Subthalamic nucleus left | -0.08 (-0.31-0.16) | .05 | .16 |
| Subthalamic nucleus right | -0.08 (-0.18-0.01) | .02 | .09 |
| Superior temporal gyrus middle part left gray matter | 0.09 (0.01-0.18) | .02 | .09 |
| Superior temporal gyrus middle part left white matter | 0.03 (-0.07-0.13) | .42 | .60 |
| Superior temporal gyrus middle part right gray matter | 0.11 (0.04-0.19) | .002 | .02 |
| Superior temporal gyrus middle part right white matter | 0.04 (-0.06-0.14) | .39 | .58 |
| Superior temporal gyrus posterior part left gray matter | 0.08 (-0.01-0.16) | .049 | .16 |
| Superior temporal gyrus posterior part left white matter | 0.03 (-0.07-0.13) | .43 | .60 |
| Superior temporal gyrus posterior part right gray matter | 0.04 (-0.05-0.12) | .35 | .54 |
| Superior temporal gyrus posterior part right white matter | 0.04 (-0.06-0.14) | .35 | .54 |
| Thalamus left high intensity part in T2 | -0.02 (-0.10-0.06) | .69 | .78 |
| Thalamus left low intensity part in T2 | 0.09 (-0.03-0.20) | .05 | .16 |
| Thalamus right high intensity part in T2 | -0.05 (-0.13-0.03) | .23 | .41 |
| Thalamus right low intensity part in T2 | 0.02 (-0.08-0.12) | .60 | .73 |
| *Interaction* | | | |
| Amygdala left | 0.01 (-0.04-0.07) | .93 | .96 |
| Caudate nucleus left | 0.02 (-0.03-0.07) | .004 | .03 |
| Caudate nucleus right | 0.01 (-0.04-0.07) | .01 | .06 |
| Cerebrospinal fluid | -0.01 (-0.07-0.04) | <.001 | <.001 |
| Gyri parahippocampalis et ambiens anterior part left white matter | 0.01 (-0.04-0.07) | .01 | .04 |
| Gyri parahippocampalis et ambiens anterior part right white matter | 0.01 (-0.05-0.07) | .04 | .14 |
| Gyri parahippocampalis et ambiens posterior part left white matter | 0.01 (-0.05-0.07) | .04 | .16 |
| Lateral occipitotemporal gyrus gyrus fusiformis anterior part left gray matter | 0.02 (-0.04-0.07) | .01 | .06 |
| Lateral occipitotemporal gyrus gyrus fusiformis anterior part right gray matter | 0.02 (-0.03-0.07) | .01 | .08 |
| Lateral occipitotemporal gyrus gyrus fusiformis posterior part right gray matter | 0.01 (-0.04-0.07) | .02 | .09 |
| Lateral ventricle left | -0.01 (-0.07-0.05) | .001 | .02 |
| Lateral ventricle right | -0.01 (-0.06-0.05) | .003 | .03 |
| Medial and inferior temporal gyri anterior part left gray matter | 0.02 (-0.02-0.06) | .01 | .07 |
| Medial and inferior temporal gyri anterior part right white matter | -0.01 (-0.07-0.04) | .03 | .11 |
| Medial and inferior temporal gyri posterior part right gray matter | 0.02 (-0.01-0.06) | .02 | .09 |
| Subthalamic nucleus left | -0.01 (-0.05-0.03) | .04 | .16 |
| *Maternal education* | | | |
| Amygdala left | 0.00 (-0.21-0.21) | .02 | .10 |
| Amygdala right | 0.08 (-0.01-0.18) | .049 | .16 |
| Anterior temporal lobe lateral part left gray matter | -0.003 (-0.09-0.08) | .93 | .96 |
| Anterior temporal lobe lateral part left white matter | 0.05 (-0.05-0.15) | .21 | .39 |
| Anterior temporal lobe lateral part right gray matter | 0.03 (-0.06-0.11) | .47 | .62 |

| Regional measure | Standardized β coefficient (95% confidence intervals) | Raw *p* value | BH corrected *p* value |
|---|---|---|---|
| Anterior temporal lobe lateral part right white matter | 0.01 (-0.09-0.11) | .77 | .84 |
| Anterior temporal lobe medial part left gray matter | 0.04 (-0.06-0.13) | .36 | .54 |
| Anterior temporal lobe medial part left white matter | 0.01 (-0.10-0.12) | .82 | .88 |
| Anterior temporal lobe medial part right gray matter | 0.08 (-0.02-0.17) | .07 | .21 |
| Anterior temporal lobe medial part right white matter | 0.02 (-0.09-0.12) | .65 | .75 |
| Brainstem | 0.03 (-0.06-0.12) | .41 | .60 |
| Caudate nucleus left | 0.03 (-0.16-0.22) | .49 | .63 |
| Caudate nucleus right | -0.02 (-0.22-0.18) | .61 | .73 |
| Cerebellum left | 0.11 (0.04-0.18) | .001 | .02 |
| Cerebellum right | 0.09 (0.02-0.16) | .01 | .0496 |
| Cerebrospinal fluid | 0.05 (-0.16-0.26) | .02 | .09 |
| Cingulate gyrus anterior part left gray matter | -0.05 (-0.14-0.05) | .23 | .41 |
| Cingulate gyrus anterior part left white matter | 0.05 (-0.05-0.16) | .14 | .31 |
| Cingulate gyrus anterior part right gray matter | -0.05 (-0.14-0.05) | .22 | .41 |
| Cingulate gyrus anterior part right white matter | 0.05 (-0.05-0.16) | .15 | .32 |
| Cingulate gyrus posterior part left gray matter | -0.01 (-0.10-0.07) | .74 | .83 |
| Cingulate gyrus posterior part left white matter | 0.05 (-0.03-0.14) | .16 | .34 |
| Cingulate gyrus posterior part right gray matter | 0.001 (-0.09-0.09) | .99 | .99 |
| Cingulate gyrus posterior part right white matter | 0.03 (-0.06-0.12) | .44 | .60 |
| Corpus callosum | 0.06 (-0.04-0.15) | .16 | .34 |
| Frontal lobe left gray matter | -0.01 (-0.08-0.07) | .88 | .93 |
| Frontal lobe left white matter | 0.06 (-0.03-0.14) | .11 | .28 |
| Frontal lobe right gray matter | 0.01 (-0.07-0.08) | .85 | .91 |
| Frontal lobe right white matter | 0.04 (-0.05-0.13) | .31 | .50 |
| Gyri parahippocampalis et ambiens anterior part left gray matter | -0.02 (-0.13-0.09) | .62 | .73 |
| Gyri parahippocampalis et ambiens anterior part left white matter | 0.02 (-0.19-0.24) | .52 | .66 |
| Gyri parahippocampalis et ambiens anterior part right gray matter | 0.01 (-0.09-0.11) | .78 | .85 |
| Gyri parahippocampalis et ambiens anterior part right white matter | 0.03 (-0.18-0.25) | .41 | .60 |
| Gyri parahippocampalis et ambiens posterior part left gray matter | 0.02 (-0.08-0.12) | .58 | .71 |
| Gyri parahippocampalis et ambiens posterior part left white matter | 0.03 (-0.20-0.26) | .30 | .48 |
| Gyri parahippocampalis et ambiens posterior part right gray matter | 0.01 (-0.09-0.11) | .89 | .94 |
| Gyri parahippocampalis et ambiens posterior part right white matter | 0.03 (-0.07-0.13) | .42 | .60 |
| Hippocampus left | 0.06 (-0.04-0.16) | .14 | .32 |
| Hippocampus right | 0.05 (-0.05-0.16) | .13 | .31 |
| Insula left gray matter | -0.003 (-0.08-0.08) | .94 | .96 |
| Insula left white matter | 0.10 (0.01-0.19) | .01 | .06 |
| Insula right gray matter | 0.03 (-0.05-0.11) | .47 | .62 |
| Insula right white matter | 0.05 (-0.04-0.14) | .22 | .41 |

| Regional volume | Standardized β coefficient (95% confidence intervals) | Raw *p* value | BH corrected *p* value |
|---|---|---|---|
| Lateral occipitotemporal gyrus gyrus fusiformis anterior part left gray matter | -0.04 (-0.24-0.16) | .28 | .47 |
| Lateral occipitotemporal gyrus gyrus fusiformis anterior part left white matter | 0.15 (0.05-0.25) | <.001 | .01 |
| Lateral occipitotemporal gyrus gyrus fusiformis anterior part right gray matter | -0.02 (-0.21-0.17) | .64 | .74 |
| Lateral occipitotemporal gyrus gyrus fusiformis anterior part right white matter | 0.10 (0.001-0.20) | .02 | .09 |
| Lateral occipitotemporal gyrus gyrus fusiformis posterior part left gray matter | 0.03 (-0.07-0.13) | .49 | .63 |
| Lateral occipitotemporal gyrus gyrus fusiformis posterior part left white matter | 0.05 (-0.05-0.15) | .24 | .41 |
| Lateral occipitotemporal gyrus gyrus fusiformis posterior part right gray matter | -0.003 (-0.21-0.20) | .93 | .96 |
| Lateral occipitotemporal gyrus gyrus fusiformis posterior part right white matter | 0.02 (-0.08-0.12) | .59 | .72 |
| Lateral ventricle left | 0.04 (-0.18-0.26) | .09 | .25 |
| Lateral ventricle right | 0.04 (-0.18-0.25) | .10 | .26 |
| Lentiform nucleus left | -0.01 (-0.09-0.08) | .90 | .95 |
| Lentiform nucleus right | 0.03 (-0.05-0.11) | .42 | .60 |
| Medial and inferior temporal gyri anterior part left gray matter | -0.07 (-0.22-0.09) | .12 | .28 |
| Medial and inferior temporal gyri anterior part left white matter | 0.07 (-0.02-0.16) | .08 | .23 |
| Medial and inferior temporal gyri anterior part right gray matter | 0.02 (-0.05-0.08) | .63 | .73 |
| Medial and inferior temporal gyri anterior part right white matter | 0.08 (-0.13-0.29) | .05 | .17 |
| Medial and inferior temporal gyri posterior part left gray matter | -0.02 (-0.09-0.05) | .51 | .65 |
| Medial and inferior temporal gyri posterior part left white matter | 0.06 (-0.04-0.16) | .16 | .34 |
| Medial and inferior temporal gyri posterior part right gray matter | -0.05 (-0.19-0.09) | .28 | .47 |
| Medial and inferior temporal gyri posterior part right white matter | 0.05 (-0.05-0.15) | .23 | .41 |
| Occipital lobe left gray matter | 0.05 (-0.02-0.11) | .18 | .36 |
| Occipital lobe left white matter | 0.02 (-0.08-0.13) | .58 | .71 |
| Occipital lobe right gray matter | 0.05 (-0.02-0.12) | .15 | .33 |
| Occipital lobe right white matter | 0.06 (-0.04-0.16) | .14 | .32 |
| Parietal lobe left gray matter | 0.04 (-0.02-0.09) | .18 | .36 |
| Parietal lobe left white matter | 0.03 (-0.06-0.12) | .44 | .60 |
| Parietal lobe right gray matter | 0.03 (-0.02-0.09) | .24 | .41 |
| Parietal lobe right white matter | 0.03 (-0.06-0.12) | .45 | .61 |
| Subthalamic nucleus left | 0.06 (-0.09-0.21) | .10 | .26 |
| Subthalamic nucleus right | 0.04 (-0.05-0.14) | .20 | .38 |
| Superior temporal gyrus middle part left gray matter | 0.01 (-0.07-0.09) | .80 | .86 |
| Superior temporal gyrus middle part left white matter | 0.11 (0.01-0.21) | .01 | .048 |
| Superior temporal gyrus middle part right gray matter | 0.01 (-0.06-0.08) | .74 | .83 |
| Superior temporal gyrus middle part right white matter | 0.06 (-0.04-0.16) | .12 | .29 |
| Superior temporal gyrus posterior part left gray matter | -0.01 (-0.09-0.07) | .75 | .83 |
| Superior temporal gyrus posterior part left white matter | 0.04 (-0.06-0.14) | .23 | .41 |

| Regional volume | Standardized β coefficient (95% confidence intervals) | Raw *p* value | BH corrected *p* value |
|---|---|---|---|
| Superior temporal gyrus posterior part right gray matter | 0.04 (-0.04-0.12) | .32 | .51 |
| Superior temporal gyrus posterior part right white matter | 0.002 (-0.10-0.10) | .96 | .98 |
| Thalamus left high intensity part in T2 | 0.03 (-0.05-0.11) | .45 | .60 |
| Thalamus left low intensity part in T2 | -0.06 (-0.17-0.06) | .18 | .36 |
| Thalamus right high intensity part in T2 | 0.01 (-0.07-0.10) | .70 | .80 |
| Thalamus right low intensity part in T2 | -0.004 (-0.10-0.09) | .92 | .96 |

Fully adjusted ridge regression model, including gestation at birth, maternal final educational qualification, gestation at MRI, the interaction term (where significant), birth weight z-score, birth head circumference z-score, sex, smoking in pregnancy, and breast milk at discharge. Corrected for false discovery rate.
BH = Benjamini-Hochberg correction.

##### eTable 7: Regional brain volumes – relationship with gestational age, paternal final educational qualification, and interaction effect (fully adjusted ridge regression model)

| Regional volume | Standardized β coefficient (95% confidence intervals) | Raw *p* value | BH corrected *p* value |
|---|---|---|---|
| *Gestation* | | | |
| Amygdala left | 0.07 (-0.23-0.37) | .07 | .22 |
| Amygdala right | 0.04 (-0.06-0.14) | .37 | .61 |
| Anterior temporal lobe lateral part left gray matter | 0.13 (0.04-0.23) | .002 | .02 |
| Anterior temporal lobe lateral part left white matter | 0.10 (-0.01-0.20) | .04 | .16 |
| Anterior temporal lobe lateral part right gray matter | 0.04 (-0.05-0.13) | .29 | .51 |
| Anterior temporal lobe lateral part right white matter | 0.10 (-0.01-0.20) | .04 | .16 |
| Anterior temporal lobe medial part left gray matter | 0.04 (-0.06-0.14) | .41 | .61 |
| Anterior temporal lobe medial part left white matter | 0.03 (-0.08-0.15) | .38 | .61 |
| Anterior temporal lobe medial part right gray matter | 0.03 (-0.07-0.13) | .47 | .67 |
| Anterior temporal lobe medial part right white matter | 0.05 (-0.06-0.16) | .24 | .50 |
| Brainstem | -0.01 (-0.10-0.09) | .90 | .93 |
| Caudate nucleus left | 0.11 (-0.16-0.38) | .004 | .03 |
| Caudate nucleus right | 0.15 (0.06-0.25) | <.001 | .004 |
| Cerebellum left | -0.02 (-0.10-0.06) | .61 | .75 |
| Cerebellum right | -0.04 (-0.11-0.04) | .31 | .54 |
| Cerebrospinal fluid | -0.20 (-0.51-0.11) | <.001 | <.001 |
| Cingulate gyrus anterior part left gray matter | 0.03 (-0.07-0.13) | .41 | .61 |
| Cingulate gyrus anterior part left white matter | 0.04 (-0.27-0.36) | .18 | .43 |
| Cingulate gyrus anterior part right gray matter | -0.04 (-0.14-0.06) | .32 | .54 |
| Cingulate gyrus anterior part right white matter | 0.06 (-0.05-0.17) | .12 | .33 |
| Cingulate gyrus posterior part left gray matter | -0.07 (-0.16-0.03) | .13 | .34 |
| Cingulate gyrus posterior part left white matter | 0.05 (-0.04-0.15) | .20 | .44 |
| Cingulate gyrus posterior part right gray matter | -0.11 (-0.21- -0.01) | .01 | .08 |
| Cingulate gyrus posterior part right white matter | 0.08 (-0.01-0.18) | .05 | .17 |
| Corpus callosum | 0.07 (-0.03-0.17) | .09 | .27 |
| Frontal lobe left gray matter | 0.03 (-0.06-0.11) | .51 | .70 |
| Frontal lobe left white matter | 0.22 (0.13-0.31) | <.001 | <.001 |
| Frontal lobe right gray matter | 0.03 (-0.05-0.11) | .39 | .61 |
| Frontal lobe right white matter | 0.18 (0.09-0.27) | <.001 | <.001 |
| Gyri parahippocampalis et ambiens anterior part left gray matter | 0.05 (-0.28-0.39) | .15 | .38 |
| Gyri parahippocampalis et ambiens anterior part left white matter | 0.11 (-0.20-0.41) | .01 | .047 |
| Gyri parahippocampalis et ambiens anterior part right gray matter | 0.04 (-0.27-0.35) | .32 | .54 |
| Gyri parahippocampalis et ambiens anterior part right white matter | 0.03 (-0.28-0.33) | .45 | .65 |
| Gyri parahippocampalis et ambiens posterior part left gray matter | 0.14 (0.03-0.24) | .002 | .02 |
| Gyri parahippocampalis et ambiens posterior part left white matter | 0.07 (-0.04-0.19) | .05 | .17 |

| Regional volume | Standardized β coefficient (95% confidence intervals) | Raw *p* value | BH corrected *p* value |
|---|---|---|---|
| Gyri parahippocampalis et ambiens posterior part right gray matter | 0.05 (-0.05-0.16) | .25 | .50 |
| Gyri parahippocampalis et ambiens posterior part right white matter | 0.15 (0.04-0.25) | <.001 | .01 |
| Hippocampus left | -0.04 (-0.15-0.06) | .25 | .50 |
| Hippocampus right | -0.02 (-0.13-0.09) | .56 | .74 |
| Insula left gray matter | 0.04 (-0.05-0.12) | .34 | .57 |
| Insula left white matter | 0.10 (-0.18-0.38) | .02 | .11 |
| Insula right gray matter | -0.02 (-0.10-0.06) | .62 | .76 |
| Insula right white matter | 0.12 (-0.16-0.40) | .004 | .03 |
| Lateral occipitotemporal gyrus gyrus fusiformis anterior part left gray matter | 0.24 (0.14-0.34) | <.001 | <.001 |
| Lateral occipitotemporal gyrus gyrus fusiformis anterior part left white matter | -0.07 (-0.18-0.03) | .10 | .29 |
| Lateral occipitotemporal gyrus gyrus fusiformis anterior part right gray matter | 0.09 (-0.18-0.36) | .06 | .20 |
| Lateral occipitotemporal gyrus gyrus fusiformis anterior part right white matter | -0.04 (-0.35-0.27) | .27 | .50 |
| Lateral occipitotemporal gyrus gyrus fusiformis posterior part left gray matter | 0.13 (0.03-0.23) | .004 | .03 |
| Lateral occipitotemporal gyrus gyrus fusiformis posterior part left white matter | -0.03 (-0.13-0.07) | .52 | .70 |
| Lateral occipitotemporal gyrus gyrus fusiformis posterior part right gray matter | 0.09 (-0.01-0.19) | .04 | .17 |
| Lateral occipitotemporal gyrus gyrus fusiformis posterior part right white matter | 0.01 (-0.09-0.12) | .69 | .81 |
| Lateral ventricle left | -0.09 (-0.41-0.23) | <.001 | .004 |
| Lateral ventricle right | -0.09 (-0.39-0.22) | <.001 | .01 |
| Lentiform nucleus left | -0.09 (-0.18-0.002) | .04 | .17 |
| Lentiform nucleus right | -0.12 (-0.21- -0.04) | .003 | .03 |
| Medial and inferior temporal gyri anterior part left gray matter | 0.23 (0.16-0.31) | <.001 | <.001 |
| Medial and inferior temporal gyri anterior part left white matter | 0.05 (-0.06-0.15) | .29 | .51 |
| Medial and inferior temporal gyri anterior part right gray matter | 0.23 (0.16-0.30) | <.001 | <.001 |
| Medial and inferior temporal gyri anterior part right white matter | 0.02 (-0.08-0.13) | .58 | .74 |
| Medial and inferior temporal gyri posterior part left gray matter | 0.12 (0.05-0.19) | <.001 | .01 |
| Medial and inferior temporal gyri posterior part left white matter | -0.03 (-0.14-0.07) | .41 | .61 |
| Medial and inferior temporal gyri posterior part right gray matter | 0.13 (0.06-0.20) | <.001 | .002 |
| Medial and inferior temporal gyri posterior part right white matter | -0.01 (-0.12-0.09) | .72 | .82 |
| Occipital lobe left gray matter | -0.02 (-0.09-0.05) | .57 | .74 |
| Occipital lobe left white matter | -0.04 (-0.14-0.06) | .34 | .57 |
| Occipital lobe right gray matter | 0.01 (-0.07-0.09) | .72 | .82 |
| Occipital lobe right white matter | -0.02 (-0.13-0.08) | .60 | .75 |
| Parietal lobe left gray matter | 0.06 (-0.001-0.12) | .05 | .17 |
| Parietal lobe left white matter | 0.13 (0.03-0.23) | .002 | .02 |
| Parietal lobe right gray matter | 0.09 (0.03-0.15) | .004 | .03 |

| Regional volume | Standardized β coefficient (95% confidence intervals) | Raw *p* value | BH corrected *p* value |
|---|---|---|---|
| Parietal lobe right white matter | 0.09 (-0.01-0.19) | .04 | .16 |
| Subthalamic nucleus left | -0.07 (-0.22-0.09) | .05 | .17 |
| Subthalamic nucleus right | -0.03 (-0.31-0.24) | .16 | .40 |
| Superior temporal gyrus middle part left gray matter | 0.09 (-0.002-0.17) | .04 | .16 |
| Superior temporal gyrus middle part left white matter | 0.06 (-0.05-0.16) | .18 | .42 |
| Superior temporal gyrus middle part right gray matter | 0.10 (0.02-0.18) | .01 | .047 |
| Superior temporal gyrus middle part right white matter | 0.06 (-0.05-0.16) | .17 | .42 |
| Superior temporal gyrus posterior part left gray matter | 0.08 (-0.01-0.17) | .06 | .20 |
| Superior temporal gyrus posterior part left white matter | 0.04 (-0.07-0.15) | .34 | .57 |
| Superior temporal gyrus posterior part right gray matter | 0.03 (-0.06-0.12) | .43 | .64 |
| Superior temporal gyrus posterior part right white matter | 0.04 (-0.06-0.15) | .33 | .56 |
| Thalamus left high intensity part in T2 | -0.002 (-0.09-0.08) | .96 | .97 |
| Thalamus left low intensity part in T2 | 0.005 (-0.004-0.01) | .25 | .50 |
| Thalamus right high intensity part in T2 | -0.05 (-0.13-0.04) | .25 | .50 |
| Thalamus right low intensity part in T2 | -0.01 (-0.11-0.10) | .88 | .92 |
| *Interaction* | | | |
| Amygdala left | 0.01 (-0.05-0.06) | .04 | .17 |
| Caudate nucleus left | 0.01 (-0.04-0.06) | .01 | .0499 |
| Cerebrospinal fluid | -0.02 (-0.08-0.04) | <.001 | <.001 |
| Cingulate gyrus anterior part left white matter | 0.01 (-0.05-0.07) | .05 | .17 |
| Gyri parahippocampalis et ambiens anterior part left gray matter | 0.01 (-0.05-0.07) | .03 | .16 |
| Gyri parahippocampalis et ambiens anterior part left white matter | 0.02 (-0.04-0.07) | .001 | .01 |
| Gyri parahippocampalis et ambiens anterior part right gray matter | 0.01 (-0.05-0.07) | .04 | .17 |
| Gyri parahippocampalis et ambiens anterior part right white matter | 0.02 (-0.04-0.07) | .001 | .02 |
| Insula left white matter | 0.02 (-0.17-0.20) | .01 | .06 |
| Insula right white matter | 0.01 (-0.04-0.06) | .04 | .17 |
| Lateral occipitotemporal gyrus gyrus fusiformis anterior part right gray matter | 0.02 (-0.03-0.07) | .02 | .12 |
| Lateral occipitotemporal gyrus gyrus fusiformis anterior part right white matter | 0.01 (-0.05-0.07) | .05 | .17 |
| Lateral ventricle left | -0.01 (-0.07-0.05) | .002 | .02 |
| Lateral ventricle right | -0.05 (-0.07-0.05) | .002 | .02 |
| Subthalamic nucleus left | -0.01 (-0.04-0.02) | .003 | .03 |
| Subthalamic nucleus right | -0.01 (-0.06-0.05) | .05 | .17 |
| *Paternal education* | | | |
| Amygdala left | -0.02 (-0.23-0.18) | .48 | .67 |
| Amygdala right | 0.01 (-0.07-0.10) | .69 | .81 |
| Anterior temporal lobe lateral part left gray matter | -0.01 (-0.08-0.07) | .81 | .88 |
| Anterior temporal lobe lateral part left white matter | 0.04 (-0.05-0.13) | .28 | .50 |
| Anterior temporal lobe lateral part right gray matter | 0.05 (-0.03-0.12) | .17 | .42 |

| Regional volume | Standardized β coefficient (95% confidence intervals) | Raw *p* value | BH corrected *p* value |
|---|---|---|---|
| Anterior temporal lobe lateral part right white matter | 0.02 (-0.07-0.11) | .69 | .81 |
| Anterior temporal lobe medial part left gray matter | 0.03 (-0.05-0.11) | .40 | .61 |
| Anterior temporal lobe medial part left white matter | 0.01 (-0.09-0.10) | .84 | .89 |
| Anterior temporal lobe medial part right gray matter | 0.04 (-0.04-0.13) | .28 | .50 |
| Anterior temporal lobe medial part right white matter | 0.02 (-0.07-0.11) | .57 | .74 |
| Brainstem | 0.01 (-0.06-0.09) | .69 | .81 |
| Caudate nucleus left | 0.04 (-0.14-0.23) | .20 | .44 |
| Caudate nucleus right | 0.08 (0.01-0.16) | .02 | .09 |
| Cerebellum left | 0.06 (-0.004-0.12) | .05 | .18 |
| Cerebellum right | 0.05 (-0.01-0.11) | .09 | .27 |
| Cerebrospinal fluid | 0.04 (-0.17-0.25) | .19 | .44 |
| Cingulate gyrus anterior part left gray matter | 0.002 (-0.08-0.09) | .94 | .96 |
| Cingulate gyrus anterior part left white matter | 0.03 (-0.18-0.25) | .25 | .50 |
| Cingulate gyrus anterior part right gray matter | -0.01 (-0.10-0.07) | .63 | .76 |
| Cingulate gyrus anterior part right white matter | 0.02 (-0.07-0.11) | .52 | .70 |
| Cingulate gyrus posterior part left gray matter | -0.01 (-0.08-0.07) | .83 | .89 |
| Cingulate gyrus posterior part left white matter | 0.05 (-0.02-0.13) | .14 | .36 |
| Cingulate gyrus posterior part right gray matter | 0.02 (-0.06-0.11) | .52 | .70 |
| Cingulate gyrus posterior part right white matter | 0.02 (-0.06-0.10) | .57 | .74 |
| Corpus callosum | 0.02 (-0.06-0.11) | .51 | .70 |
| Frontal lobe left gray matter | -0.01 (-0.08-0.05) | .67 | .80 |
| Frontal lobe left white matter | 0.03 (-0.04-0.10) | .36 | .58 |
| Frontal lobe right gray matter | -0.02 (-0.08-0.05) | .58 | .74 |
| Frontal lobe right white matter | 0.03 (-0.05-0.10) | .41 | .61 |
| Gyri parahippocampalis et ambiens anterior part left gray matter | -0.04 (-0.26-0.19) | .27 | .50 |
| Gyri parahippocampalis et ambiens anterior part left white matter | -0.07 (-0.29-0.14) | .03 | .15 |
| Gyri parahippocampalis et ambiens anterior part right gray matter | -0.03 (-0.24-0.18) | .44 | .64 |
| Gyri parahippocampalis et ambiens anterior part right white matter | -0.06 (-0.26-0.15) | .10 | .28 |
| Gyri parahippocampalis et ambiens posterior part left gray matter | 0.01 (-0.07-0.10) | .73 | .83 |
| Gyri parahippocampalis et ambiens posterior part left white matter | 0.02 (-0.07-0.11) | .52 | .70 |
| Gyri parahippocampalis et ambiens posterior part right gray matter | -0.0003 (-0.09-0.09) | .99 | .99 |
| Gyri parahippocampalis et ambiens posterior part right white matter | -0.02 (-0.11-0.07) | .59 | .74 |
| Hippocampus left | 0.01 (-0.08-0.10) | .78 | .88 |
| Hippocampus right | 0.01 (-0.09-0.10) | .79 | .88 |
| Insula left gray matter | -0.04 (-0.11-0.03) | .26 | .50 |
| Insula left white matter | 0.02 (-0.04-0.07) | .67 | .80 |
| Insula right gray matter | -0.04 (-0.10-0.03) | .27 | .50 |
| Insula right white matter | 0.001 (-0.19-0.19) | .97 | .98 |

| **Regional volume** | **Standardized β coefficient (95% confidence intervals)** | **Raw *p* value** | **BH corrected *p* value** |
|---|---|---|---|
| Lateral occipitotemporal gyrus gyrus fusiformis anterior part left gray matter | 0.03 (-0.05-0.11) | .38 | .61 |
| Lateral occipitotemporal gyrus gyrus fusiformis anterior part left white matter | 0.08 (-0.01-0.17) | .03 | .16 |
| Lateral occipitotemporal gyrus gyrus fusiformis anterior part right gray matter | -0.01 (-0.19-0.17) | .79 | .88 |
| Lateral occipitotemporal gyrus gyrus fusiformis anterior part right white matter | 0.05 (-0.16-0.26) | .10 | .29 |
| Lateral occipitotemporal gyrus gyrus fusiformis posterior part left gray matter | 0.06 (-0.02-0.15) | .09 | .27 |
| Lateral occipitotemporal gyrus gyrus fusiformis posterior part left white matter | 0.05 (-0.04-0.13) | .19 | .44 |
| Lateral occipitotemporal gyrus gyrus fusiformis posterior part right gray matter | 0.06 (-0.02-0.14) | .09 | .27 |
| Lateral occipitotemporal gyrus gyrus fusiformis posterior part right white matter | 0.04 (-0.05-0.13) | .21 | .45 |
| Lateral ventricle left | 0.05 (-0.17-0.26) | .04 | .16 |
| Lateral ventricle right | -0.01 (-0.16-0.26) | .02 | .10 |
| Lentiform nucleus left | -0.003 (-0.08-0.07) | .94 | .96 |
| Lentiform nucleus right | 0.004 (-0.07-0.07) | .91 | .93 |
| Medial and inferior temporal gyri anterior part left gray matter | 0.03 (-0.03-0.09) | .26 | .50 |
| Medial and inferior temporal gyri anterior part left white matter | 0.05 (-0.04-0.13) | .19 | .44 |
| Medial and inferior temporal gyri anterior part right gray matter | 0.01 (-0.04-0.07) | .60 | .75 |
| Medial and inferior temporal gyri anterior part right white matter | -0.01 (-0.10-0.08) | .82 | .88 |
| Medial and inferior temporal gyri posterior part left gray matter | 0.01 (-0.05-0.07) | .65 | .79 |
| Medial and inferior temporal gyri posterior part left white matter | 0.05 (-0.04-0.14) | .14 | .37 |
| Medial and inferior temporal gyri posterior part right gray matter | 0.05 (-0.004-0.11) | .06 | .19 |
| Medial and inferior temporal gyri posterior part right white matter | 0.04 (-0.05-0.13) | .24 | .50 |
| Occipital lobe left gray matter | -0.004 (-0.06-0.06) | .88 | .92 |
| Occipital lobe left white matter | -0.03 (-0.12-0.06) | .41 | .61 |
| Occipital lobe right gray matter | 0.01 (-0.06-0.07) | .81 | .88 |
| Occipital lobe right white matter | -0.03 (-0.12-0.06) | .46 | .66 |
| Parietal lobe left gray matter | 0.02 (-0.03-0.07) | .49 | .68 |
| Parietal lobe left white matter | 0.005 (-0.08-0.09) | .90 | .93 |
| Parietal lobe right gray matter | 0.02 (-0.03-0.07) | .48 | .68 |
| Parietal lobe right white matter | -0.01 (-0.09-0.07) | .81 | .88 |
| Subthalamic nucleus left | 0.01 (-0.09-0.11) | .83 | .89 |
| Subthalamic nucleus right | 0.01 (-0.18-0.19) | .78 | .88 |
| Superior temporal gyrus middle part left gray matter | 0.02 (-0.05-0.09) | .57 | .74 |
| Superior temporal gyrus middle part left white matter | 0.04 (-0.04-0.13) | .20 | .44 |
| Superior temporal gyrus middle part right gray matter | 0.05 (-0.01-0.11) | .11 | .29 |
| Superior temporal gyrus middle part right white matter | 0.05 (-0.04-0.13) | .16 | .41 |
| Superior temporal gyrus posterior part left gray matter | -0.02 (-0.10- -0.10) | .50 | .70 |
| Superior temporal gyrus posterior part left white matter | 0.04 (-0.05-0.13) | .27 | .50 |

| Regional volume | Standardized β coefficient (95% confidence intervals) | Raw *p* value | BH corrected *p* value |
|---|---|---|---|
| Superior temporal gyrus posterior part right gray matter | 0.04 (-0.03-0.12) | .23 | .50 |
| Superior temporal gyrus posterior part right white matter | -0.01 (-0.09-0.08) | .85 | .90 |
| Thalamus left high intensity part in T2 | -0.04 (-0.10-0.03) | .29 | .51 |
| Thalamus left low intensity part in T2 | 0.001 (-0.01-0.01) | .71 | .82 |
| Thalamus right high intensity part in T2 | -0.04 (-0.11-0.03) | .26 | .50 |
| Thalamus right low intensity part in T2 | 0.03 (-0.05-0.12) | .39 | .61 |

Fully adjusted ridge regression model, including gestation at birth, paternal final educational qualification, gestation at MRI, the interaction term (where significant), birth weight z-score, birth head circumference z-score, sex, smoking in pregnancy, and breast milk at discharge. Corrected for false discovery rate.
BH = Benjamini-Hochberg correction.

#### eTable 8: Regional brain volumes – relationship with gestational age, maternal occupation, and interaction effect (fully adjusted ridge regression model)

| Regional volume | Standardized β coefficient (95% confidence intervals) | Raw *p* value | BH corrected *p* value |
|---|---|---|---|
| *Gestation* | | | |
| Amygdala left | 0.08 (-0.01-0.18) | .04 | .17 |
| Amygdala right | 0.03 (-0.07-0.12) | .52 | .72 |
| Anterior temporal lobe lateral part left gray matter | 0.14 (0.06-0.22) | <.001 | .01 |
| Anterior temporal lobe lateral part left white matter | 0.11 (0.01-0.21) | .01 | .07 |
| Anterior temporal lobe lateral part right gray matter | 0.06 (-0.02-0.15) | .11 | .28 |
| Anterior temporal lobe lateral part right white matter | 0.10 (-0.003-0.19) | .02 | .11 |
| Anterior temporal lobe medial part left gray matter | 0.04 (-0.05-0.13) | .38 | .62 |
| Anterior temporal lobe medial part left white matter | 0.03 (-0.07-0.14) | .36 | .60 |
| Anterior temporal lobe medial part right gray matter | 0.05 (-0.04-0.14) | .25 | .50 |
| Anterior temporal lobe medial part right white matter | 0.06 (-0.05-0.16) | .19 | .42 |
| Brainstem | -0.03 (-0.11-0.06) | .52 | .72 |
| Caudate nucleus left | 0.06 (-0.24-0.36) | .18 | .40 |
| Caudate nucleus right | 0.07 (-0.24-0.39) | .06 | .21 |
| Cerebellum left | -0.02 (-0.09-0.05) | .57 | .75 |
| Cerebellum right | -0.04 (-0.11-0.03) | .28 | .53 |
| Cerebrospinal fluid | -0.16 (-0.48-0.16) | .002 | .02 |
| Cingulate gyrus anterior part left gray matter | 0.02 (-0.08-0.11) | .69 | .83 |
| Cingulate gyrus anterior part left white matter | 0.07 (-0.03-0.17) | .06 | .21 |
| Cingulate gyrus anterior part right gray matter | -0.06 (-0.16-0.04) | .12 | .30 |
| Cingulate gyrus anterior part right white matter | 0.04 (-0.06-0.14) | .32 | .57 |
| Cingulate gyrus posterior part left gray matter | -0.07 (-0.16-0.01) | .07 | .22 |
| Cingulate gyrus posterior part left white matter | 0.06 (-0.03-0.14) | .14 | .34 |
| Cingulate gyrus posterior part right gray matter | -0.11 (-0.20- -0.02) | .01 | .07 |
| Cingulate gyrus posterior part right white matter | 0.08 (-0.01-0.16) | .05 | .18 |
| Corpus callosum | 0.10 (0.004-0.19) | .02 | .09 |
| Frontal lobe left gray matter | 0.02 (-0.06-0.09) | .61 | .78 |
| Frontal lobe left white matter | 0.14 (-0.15-0.42) | <.001 | .01 |
| Frontal lobe right gray matter | 0.02 (-0.05-0.10) | .55 | .74 |
| Frontal lobe right white matter | 0.17 (0.09-0.26) | <.001 | .001 |
| Gyri parahippocampalis et ambiens anterior part left gray matter | 0.07 (-0.03-0.18) | .07 | .22 |
| Gyri parahippocampalis et ambiens anterior part left white matter | 0.07 (-0.27-0.41) | .05 | .18 |
| Gyri parahippocampalis et ambiens anterior part right gray matter | 0.07 (-0.03-0.17) | .10 | .28 |
| Gyri parahippocampalis et ambiens anterior part right white matter | 0.02 (-0.31-0.35) | .67 | .82 |
| Gyri parahippocampalis et ambiens posterior part left gray matter | 0.05 (-0.28-0.38) | .23 | .47 |
| Gyri parahippocampalis et ambiens posterior part left white matter | 0.03 (-0.33-0.38) | .39 | .62 |

| **Regional volume** | **Standardized β coefficient (95% confidence intervals)** | **Raw *p* value** | **BH corrected *p* value** |
|---|---|---|---|
| Gyri parahippocampalis et ambiens posterior part right gray matter | 0.02 (-0.08-0.12) | .60 | .77 |
| Gyri parahippocampalis et ambiens posterior part right white matter | 0.09 (-0.25-0.44) | .01 | .07 |
| Hippocampus left | -0.08 (-0.18-0.01) | .05 | .18 |
| Hippocampus right | -0.06 (-0.16-0.04) | .18 | .39 |
| Insula left gray matter | 0.03 (-0.04-0.11) | .37 | .60 |
| Insula left white matter | 0.09 (-0.21-0.39) | .03 | .13 |
| Insula right gray matter | -0.03 (-0.10-0.05) | .44 | .64 |
| Insula right white matter | 0.17 (0.08-0.25) | <.001 | .002 |
| Lateral occipitotemporal gyrus gyrus fusiformis anterior part left gray matter | 0.14 (-0.18-0.45) | .001 | .02 |
| Lateral occipitotemporal gyrus gyrus fusiformis anterior part left white matter | -0.08 (-0.17-0.02) | .08 | .23 |
| Lateral occipitotemporal gyrus gyrus fusiformis anterior part right gray matter | 0.07 (-0.22-0.37) | .12 | .30 |
| Lateral occipitotemporal gyrus gyrus fusiformis anterior part right white matter | -0.02 (-0.12-0.07) | .58 | .76 |
| Lateral occipitotemporal gyrus gyrus fusiformis posterior part left gray matter | 0.12 (0.02-0.21) | .01 | .0496 |
| Lateral occipitotemporal gyrus gyrus fusiformis posterior part left white matter | -0.04 (-0.13-0.06) | .35 | .60 |
| Lateral occipitotemporal gyrus gyrus fusiformis posterior part right gray matter | 0.08 (-0.01-0.17) | .05 | .18 |
| Lateral occipitotemporal gyrus gyrus fusiformis posterior part right white matter | 0.002 (-0.10-0.10) | .96 | .96 |
| Lateral ventricle left | -0.08 (-0.44-0.27) | <.001 | .01 |
| Lateral ventricle right | -0.07 (-0.41-0.27) | <.001 | .01 |
| Lentiform nucleus left | -0.07 (-0.16-0.01) | .06 | .21 |
| Lentiform nucleus right | 0.03 (-0.24-0.30) | .63 | .80 |
| Medial and inferior temporal gyri anterior part left gray matter | 0.07 (-0.17-0.31) | .22 | .45 |
| Medial and inferior temporal gyri anterior part left white matter | 0.04 (-0.05-0.13) | .34 | .59 |
| Medial and inferior temporal gyri anterior part right gray matter | 0.09 (-0.13-0.31) | .11 | .28 |
| Medial and inferior temporal gyri anterior part right white matter | 0.01 (-0.09-0.10) | .86 | .91 |
| Medial and inferior temporal gyri posterior part left gray matter | 0.12 (0.05-0.18) | <.001 | .01 |
| Medial and inferior temporal gyri posterior part left white matter | -0.06 (-0.16-0.03) | .14 | .33 |
| Medial and inferior temporal gyri posterior part right gray matter | 0.03 (-0.20-0.25) | .65 | .82 |
| Medial and inferior temporal gyri posterior part right white matter | -0.04 (-0.13-0.06) | .38 | .62 |
| Occipital lobe left gray matter | -0.04 (-0.10-0.03) | .28 | .53 |
| Occipital lobe left white matter | -0.08 (-0.18-0.02) | .06 | .21 |
| Occipital lobe right gray matter | -0.01 (-0.08-0.06) | .83 | .90 |
| Occipital lobe right white matter | -0.06 (-0.16-0.03) | .14 | .34 |
| Parietal lobe left gray matter | 0.06 (0.01-0.12) | .03 | .13 |
| Parietal lobe left white matter | 0.11 (0.02-0.20) | .01 | .06 |
| Parietal lobe right gray matter | 0.08 (0.03-0.14) | .003 | .03 |

| **Regional volume** | **Standardized β coefficient (95% confidence intervals)** | **Raw *p* value** | **BH corrected *p* value** |
|---|---|---|---|
| Parietal lobe right white matter | 0.07 (-0.02-0.16) | .10 | .28 |
| Subthalamic nucleus left | -0.07 (-0.31-0.17) | .09 | .27 |
| Subthalamic nucleus right | -0.09 (-0.19-0.01) | .01 | .08 |
| Superior temporal gyrus middle part left gray matter | 0.08 (-0.004-0.16) | .05 | .18 |
| Superior temporal gyrus middle part left white matter | 0.05 (-0.05-0.14) | .24 | .48 |
| Superior temporal gyrus middle part right gray matter | 0.10 (0.03-0.17) | .003 | .03 |
| Superior temporal gyrus middle part right white matter | 0.05 (-0.05-0.14) | .25 | .50 |
| Superior temporal gyrus posterior part left gray matter | 0.07 (-0.01-0.15) | .09 | .26 |
| Superior temporal gyrus posterior part left white matter | 0.03 (-0.07-0.13) | .36 | .60 |
| Superior temporal gyrus posterior part right gray matter | 0.04 (-0.04-0.12) | .27 | .51 |
| Superior temporal gyrus posterior part right white matter | 0.04 (-0.06-0.14) | .34 | .59 |
| Thalamus left high intensity part in T2 | -0.01 (-0.09-0.07) | .80 | .89 |
| Thalamus left low intensity part in T2 | 0.02 (-0.10-0.13) | .27 | .51 |
| Thalamus right high intensity part in T2 | -0.05 (-0.13-0.03) | .20 | .42 |
| Thalamus right low intensity part in T2 | 0.01 (-0.08-0.11) | .76 | .88 |
| *Interaction* | | | |
| Caudate nucleus left | 0.01 (-0.02-0.05) | .001 | .02 |
| Caudate nucleus right | 0.01 (-0.03-0.05) | .002 | .03 |
| Cerebrospinal fluid | -0.03 (-0.07-0.01) | <.001 | <.001 |
| Frontal lobe left white matter | 0.01 (-0.03-0.05) | .02 | .09 |
| Gyri parahippocampalis et ambiens anterior part left white matter | 0.01 (-0.03-0.06) | <.001 | .003 |
| Gyri parahippocampalis et ambiens anterior part right white matter | 0.01 (-0.03-0.05) | .01 | .07 |
| Gyri parahippocampalis et ambiens posterior part left gray matter | 0.01 (-0.03-0.05) | .02 | .09 |
| Gyri parahippocampalis et ambiens posterior part left white matter | 0.01 (-0.04-0.05) | .02 | .09 |
| Gyri parahippocampalis et ambiens posterior part right white matter | 0.01 (-0.04-0.05) | .03 | .13 |
| Insula left white matter | 0.01 (-0.03-0.05) | .003 | .03 |
| Lateral occipitotemporal gyrus gyrus fusiformis anterior part left gray matter | 0.01 (-0.03-0.05) | .003 | .03 |
| Lateral occipitotemporal gyrus gyrus fusiformis anterior part right gray matter | 0.01 (-0.02-0.05) | .01 | .06 |
| Lateral ventricle left | -0.01 (-0.05-0.04) | .001 | .02 |
| Lateral ventricle right | -0.01 (-0.05-0.04) | <.001 | .01 |
| Lentiform nucleus right | -0.02 (-0.05-0.02) | .02 | .09 |
| Medial and inferior temporal gyri anterior part left gray matter | 0.02 (-0.01-0.05) | .002 | .02 |
| Medial and inferior temporal gyri anterior part right gray matter | 0.02 (-0.01-0.04) | .01 | .09 |
| Medial and inferior temporal gyri posterior part right gray matter | 0.01 (-0.01-0.04) | .05 | .18 |
| Subthalamic nucleus left | -0.01 (-0.04-0.02) | .004 | .04 |
| *Maternal occupation* | | | |
| Amygdala left | 0.02 (-0.04-0.08) | .46 | .67 |
| Amygdala right | -0.02 (-0.08-0.04) | .42 | .64 |

| **Regional volume** | **Standardized β coefficient (95% confidence intervals)** | **Raw *p* value** | **BH corrected *p* value** |
|---|---|---|---|
| Anterior temporal lobe lateral part left gray matter | 0.02 (-0.03-0.07) | .44 | .64 |
| Anterior temporal lobe lateral part left white matter | 0.03 (-0.03-0.10) | .25 | .50 |
| Anterior temporal lobe lateral part right gray matter | 0.04 (-0.01-0.10) | .10 | .28 |
| Anterior temporal lobe lateral part right white matter | -0.01 (-0.07-0.05) | .73 | .86 |
| Anterior temporal lobe medial part left gray matter | 0.05 (-0.01-0.11) | .09 | .27 |
| Anterior temporal lobe medial part left white matter | 0.01 (-0.06-0.07) | .82 | .90 |
| Anterior temporal lobe medial part right gray matter | 0.03 (-0.03-0.09) | .35 | .59 |
| Anterior temporal lobe medial part right white matter | 0.01 (-0.06-0.08) | .74 | .87 |
| Brainstem | 0.02 (-0.04-0.07) | .51 | .72 |
| Caudate nucleus left | -0.01 (-0.14-0.12) | .75 | .87 |
| Caudate nucleus right | -0.04 (-0.17-0.10) | .17 | .39 |
| Cerebellum left | 0.04 (-0.01-0.08) | .12 | .30 |
| Cerebellum right | 0.02 (-0.03-0.06) | .41 | .64 |
| Cerebrospinal fluid | 0.08 (-0.07-0.22) | .02 | .09 |
| Cingulate gyrus anterior part left gray matter | 0.01 (-0.05-0.07) | .64 | .81 |
| Cingulate gyrus anterior part left white matter | 0.01 (-0.05-0.08) | .55 | .74 |
| Cingulate gyrus anterior part right gray matter | 0.02 (-0.04-0.08) | .39 | .62 |
| Cingulate gyrus anterior part right white matter | 0.02 (-0.04-0.09) | .32 | .57 |
| Cingulate gyrus posterior part left gray matter | 0.01 (-0.05-0.06) | .78 | .88 |
| Cingulate gyrus posterior part left white matter | 0.01 (-0.05-0.06) | .73 | .86 |
| Cingulate gyrus posterior part right gray matter | 0.01 (-0.05-0.07) | .83 | .90 |
| Cingulate gyrus posterior part right white matter | 0.02 (-0.04-0.08) | .41 | .63 |
| Corpus callosum | -0.01 (-0.07-0.05) | .77 | .88 |
| Frontal lobe left gray matter | 0.01 (-0.03-0.06) | .54 | .74 |
| Frontal lobe left white matter | -0.004 (-0.13-0.12) | .88 | .92 |
| Frontal lobe right gray matter | 0.01 (-0.04-0.06) | .77 | .88 |
| Frontal lobe right white matter | 0.01 (-0.04-0.07) | .57 | .75 |
| Gyri parahippocampalis et ambiens anterior part left gray matter | 0.003 (-0.06-0.07) | .90 | .93 |
| Gyri parahippocampalis et ambiens anterior part left white matter | -0.003 (-0.15-0.15) | .92 | .94 |
| Gyri parahippocampalis et ambiens anterior part right gray matter | 0.02 (-0.04-0.08) | .50 | .71 |
| Gyri parahippocampalis et ambiens anterior part right white matter | 0.001 (-0.14-0.15) | .98 | .98 |
| Gyri parahippocampalis et ambiens posterior part left gray matter | 0.01 (-0.13-0.16) | .60 | .77 |
| Gyri parahippocampalis et ambiens posterior part left white matter | 0.04 (-0.11-0.20) | .05 | .18 |
| Gyri parahippocampalis et ambiens posterior part right gray matter | 0.04 (-0.03-0.10) | .18 | .39 |
| Gyri parahippocampalis et ambiens posterior part right white matter | -0.01 (-0.16-0.14) | .67 | .82 |
| Hippocampus left | 0.04 (-0.02-0.10) | .14 | .33 |
| Hippocampus right | 0.002 (-0.06-0.07) | .94 | .95 |
| Insula left gray matter | -0.02 (-0.07-0.03) | .50 | .71 |

| **Regional volume** | **Standardized β coefficient (95% confidence intervals)** | **Raw *p* value** | **BH corrected *p* value** |
|---|---|---|---|
| Insula left white matter | -0.03 (-0.16-0.11) | .33 | .59 |
| Insula right gray matter | 0.01 (-0.04-0.05) | .83 | .90 |
| Insula right white matter | 0.01 (-0.05-0.06) | .84 | .90 |
| Lateral occipitotemporal gyrus gyrus fusiformis anterior part left gray matter | -0.01 (-0.15-0.13) | .67 | .82 |
| Lateral occipitotemporal gyrus gyrus fusiformis anterior part left white matter | 0.04 (-0.03-0.10) | .19 | .42 |
| Lateral occipitotemporal gyrus gyrus fusiformis anterior part right gray matter | -0.02 (-0.15-0.11) | .49 | .70 |
| Lateral occipitotemporal gyrus gyrus fusiformis anterior part right white matter | -0.01 (-0.08-0.05) | .67 | .82 |
| Lateral occipitotemporal gyrus gyrus fusiformis posterior part left gray matter | -0.01 (-0.07-0.05) | .77 | .88 |
| Lateral occipitotemporal gyrus gyrus fusiformis posterior part left white matter | -0.02 (-0.08-0.04) | .43 | .64 |
| Lateral occipitotemporal gyrus gyrus fusiformis posterior part right gray matter | 0.06 (-0.004-0.12) | .04 | .16 |
| Lateral occipitotemporal gyrus gyrus fusiformis posterior part right white matter | -0.01 (-0.07-0.06) | .81 | .89 |
| Lateral ventricle left | 0.04 (-0.12-0.19) | .04 | .16 |
| Lateral ventricle right | 0.02 (-0.13-0.17) | .14 | .33 |
| Lentiform nucleus left | 0.01 (-0.05-0.06) | .80 | .89 |
| Lentiform nucleus right | 0.07 (-0.05-0.19) | .03 | .14 |
| Medial and inferior temporal gyri anterior part left gray matter | -0.01 (-0.12-0.09) | .66 | .82 |
| Medial and inferior temporal gyri anterior part left white matter | 0.05 (-0.01-0.11) | .07 | .21 |
| Medial and inferior temporal gyri anterior part right gray matter | 0.01 (-0.09-0.10) | .83 | .90 |
| Medial and inferior temporal gyri anterior part right white matter | 0.03 (-0.03-0.09) | .29 | .55 |
| Medial and inferior temporal gyri posterior part left gray matter | 0.002 (-0.04-0.05) | .92 | .94 |
| Medial and inferior temporal gyri posterior part left white matter | -0.004 (-0.07-0.06) | .90 | .93 |
| Medial and inferior temporal gyri posterior part right gray matter | -0.02 (-0.12-0.08) | .58 | .76 |
| Medial and inferior temporal gyri posterior part right white matter | 0.004 (-0.06-0.07) | .89 | .93 |
| Occipital lobe left gray matter | 0.05 (0.01-0.09) | .02 | .10 |
| Occipital lobe left white matter | 0.02 (-0.04-0.09) | .41 | .63 |
| Occipital lobe right gray matter | 0.06 (0.02-0.11) | .01 | .0496 |
| Occipital lobe right white matter | 0.02 (-0.04-0.09) | .43 | .64 |
| Parietal lobe left gray matter | 0.03 (-0.01-0.07) | .11 | .28 |
| Parietal lobe left white matter | 0.004 (-0.05-0.06) | .88 | .92 |
| Parietal lobe right gray matter | 0.04 (0.001-0.08) | .04 | .17 |
| Parietal lobe right white matter | 0.01 (-0.05-0.07) | .79 | .89 |
| Subthalamic nucleus left | 0.07 (-0.02-0.17) | .01 | .07 |
| Subthalamic nucleus right | 0.02 (-0.05-0.08) | .51 | .72 |
| Superior temporal gyrus middle part left gray matter | 0.02 (-0.03-0.07) | .43 | .64 |
| Superior temporal gyrus middle part left white matter | 0.03 (-0.03-0.10) | .22 | .45 |
| Superior temporal gyrus middle part right gray matter | 0.04 (-0.01-0.08) | .10 | .27 |

| **Regional volume** | **Standardized β coefficient (95% confidence intervals)** | **Raw *p* value** | **BH corrected *p* value** |
|---|---|---|---|
| Superior temporal gyrus middle part right white matter | 0.01 (-0.05-0.07) | .70 | .84 |
| Superior temporal gyrus posterior part left gray matter | 0.03 (-0.02-0.09) | .17 | .39 |
| Superior temporal gyrus posterior part left white matter | 0.02 (-0.04-0.09) | .36 | .60 |
| Superior temporal gyrus posterior part right gray matter | 0.03 (-0.03-0.08) | .31 | .57 |
| Superior temporal gyrus posterior part right white matter | -0.003 (-0.07-0.06) | .92 | .94 |
| Thalamus left high intensity part in T2 | 0.02 (-0.03-0.08) | .32 | .57 |
| Thalamus left low intensity part in T2 | 0.01 (-0.07-0.08) | .56 | .75 |
| Thalamus right high intensity part in T2 | 0.03 (-0.02-0.08) | .23 | .46 |
| Thalamus right low intensity part in T2 | 0.02 (-0.04-0.09) | .37 | .60 |

Fully adjusted ridge regression model, including gestation at birth, maternal occupation, gestation at MRI, the interaction term (where significant), birth weight z-score, birth head circumference z-score, sex, smoking in pregnancy, and breast milk at discharge. Corrected for false discovery rate.
BH = Benjamini-Hochberg correction.

#### eTable 9: Regional brain volumes – relationship with gestational age, paternal occupation, and interaction effect (fully adjusted ridge regression model)

| Regional volume | Standardized β coefficient (95% confidence intervals) | Raw *p* value | BH corrected *p* value |
|---|---|---|---|
| *Gestation* | | | |
| Amygdala left | 0.09 (-0.01-0.18) | .04 | .17 |
| Amygdala right | 0.02 (-0.07-0.12) | .59 | .76 |
| Anterior temporal lobe lateral part left gray matter | 0.14 (0.05-0.22) | <.001 | .01 |
| Anterior temporal lobe lateral part left white matter | 0.11 (0.01-0.21) | .01 | .08 |
| Anterior temporal lobe lateral part right gray matter | 0.06 (-0.02-0.15) | .12 | .31 |
| Anterior temporal lobe lateral part right white matter | 0.09 (-0.01-0.19) | .04 | .18 |
| Anterior temporal lobe medial part left gray matter | 0.03 (-0.06-0.12) | .44 | .65 |
| Anterior temporal lobe medial part left white matter | 0.03 (-0.08-0.14) | .41 | .64 |
| Anterior temporal lobe medial part right gray matter | 0.05 (-0.05-0.14) | .27 | .50 |
| Anterior temporal lobe medial part right white matter | 0.05 (-0.05-0.16) | .22 | .45 |
| Brainstem | -0.02 (-0.11-0.07) | .60 | .76 |
| Caudate nucleus left | 0.18 (0.09-0.26) | <.001 | <.001 |
| Caudate nucleus right | 0.10 (-0.28-0.48) | .01 | .04 |
| Cerebellum left | -0.01 (-0.08-0.07) | .88 | .94 |
| Cerebellum right | -0.02 (-0.09-0.04) | .48 | .69 |
| Cerebrospinal fluid | -0.20 (-0.57-0.17) | <.001 | <.001 |
| Cingulate gyrus anterior part left gray matter | 0.02 (-0.07-0.11) | .60 | .76 |
| Cingulate gyrus anterior part left white matter | 0.03 (-0.37-0.43) | .27 | .49 |
| Cingulate gyrus anterior part right gray matter | -0.06 (-0.16-0.03) | .14 | .35 |
| Cingulate gyrus anterior part right white matter | 0.04 (-0.07-0.14) | .34 | .57 |
| Cingulate gyrus posterior part left gray matter | -0.07 (-0.16-0.01) | .08 | .23 |
| Cingulate gyrus posterior part left white matter | 0.06 (-0.03-0.14) | .16 | .38 |
| Cingulate gyrus posterior part right gray matter | -0.12 (-0.21- -0.02) | .01 | .051 |
| Cingulate gyrus posterior part right white matter | 0.08 (-0.01-0.17) | .05 | .18 |
| Corpus callosum | 0.09 (-0.01-0.18) | .03 | .17 |
| Frontal lobe left gray matter | 0.02 (-0.05-0.10) | .56 | .75 |
| Frontal lobe left white matter | 0.21 (0.12-0.29) | <.001 | <.001 |
| Frontal lobe right gray matter | 0.03 (-0.05-0.10) | .47 | .69 |
| Frontal lobe right white matter | 0.17 (0.09-0.25) | <.001 | <.001 |
| Gyri parahippocampalis et ambiens anterior part left gray matter | 0.08 (-0.02-0.19) | .05 | .18 |
| Gyri parahippocampalis et ambiens anterior part left white matter | 0.07 (-0.34-0.48) | .06 | .20 |
| Gyri parahippocampalis et ambiens anterior part right gray matter | 0.08 (-0.02-0.18) | .05 | .19 |
| Gyri parahippocampalis et ambiens anterior part right white matter | 0.02 (-0.38-0.43) | .53 | .73 |
| Gyri parahippocampalis et ambiens posterior part left gray matter | 0.12 (0.02-0.22) | .004 | .03 |
| Gyri parahippocampalis et ambiens posterior part left white matter | 0.06 (-0.05-0.17) | .12 | .31 |
| Gyri parahippocampalis et ambiens posterior part right gray matter | 0.04 (-0.06-0.14) | .37 | .59 |

| Regional volume | Standardized β coefficient (95% confidence intervals) | Raw *p* value | BH corrected *p* value |
|---|---|---|---|
| Gyri parahippocampalis et ambiens posterior part right white matter | 0.14 (0.04-0.24) | <.001 | .01 |
| Hippocampus left | -0.06 (-0.16-0.03) | .12 | .31 |
| Hippocampus right | -0.05 (-0.16-0.05) | .21 | .45 |
| Insula left gray matter | 0.03 (-0.04-0.11) | .36 | .58 |
| Insula left white matter | 0.11 (-0.25-0.47) | .004 | .03 |
| Insula right gray matter | -0.03 (-0.10-0.05) | .49 | .70 |
| Insula right white matter | 0.11 (-0.25-0.47) | .003 | .03 |
| Lateral occipitotemporal gyrus gyrus fusiformis anterior part left gray matter | 0.22 (0.13-0.31) | <.001 | <.001 |
| Lateral occipitotemporal gyrus gyrus fusiformis anterior part left white matter | -0.08 (-0.18-0.02) | .06 | .20 |
| Lateral occipitotemporal gyrus gyrus fusiformis anterior part right gray matter | 0.17 (0.08-0.25) | <.001 | <.001 |
| Lateral occipitotemporal gyrus gyrus fusiformis anterior part right white matter | -0.02 (-0.12-0.08) | .57 | .76 |
| Lateral occipitotemporal gyrus gyrus fusiformis posterior part left gray matter | 0.12 (0.02-0.21) | .01 | .04 |
| Lateral occipitotemporal gyrus gyrus fusiformis posterior part left white matter | -0.03 (-0.13-0.06) | .41 | .64 |
| Lateral occipitotemporal gyrus gyrus fusiformis posterior part right gray matter | 0.08 (-0.01-0.18) | .04 | .18 |
| Lateral occipitotemporal gyrus gyrus fusiformis posterior part right white matter | 0.01 (-0.09-0.11) | .73 | .87 |
| Lateral ventricle left | -0.08 (-0.49-0.33) | .001 | .01 |
| Lateral ventricle right | -0.07 (-0.47-0.33) | .001 | .01 |
| Lentiform nucleus left | -0.08 (-0.16-0.01) | .06 | .20 |
| Lentiform nucleus right | 0.01 (-0.31-0.33) | .90 | .95 |
| Medial and inferior temporal gyri anterior part left gray matter | 0.22 (0.15-0.29) | <.001 | <.001 |
| Medial and inferior temporal gyri anterior part left white matter | 0.05 (-0.04-0.14) | .24 | .46 |
| Medial and inferior temporal gyri anterior part right gray matter | 0.12 (-0.15-0.39) | .04 | .18 |
| Medial and inferior temporal gyri anterior part right white matter | 0.02 (-0.08-0.12) | .59 | .76 |
| Medial and inferior temporal gyri posterior part left gray matter | 0.11 (0.04-0.17) | .001 | .01 |
| Medial and inferior temporal gyri posterior part left white matter | -0.05 (-0.15-0.05) | .22 | .45 |
| Medial and inferior temporal gyri posterior part right gray matter | 0.13 (0.06-0.19) | <.001 | .002 |
| Medial and inferior temporal gyri posterior part right white matter | -0.02 (-0.12-0.08) | .54 | .73 |
| Occipital lobe left gray matter | 0.10 (-0.17-0.38) | .13 | .32 |
| Occipital lobe left white matter | -0.07 (-0.17-0.03) | .12 | .31 |
| Occipital lobe right gray matter | 0.14 (-0.16-0.44) | .03 | .16 |
| Occipital lobe right white matter | -0.05 (-0.15-0.05) | .23 | .45 |
| Parietal lobe left gray matter | 0.06 (0.002-0.12) | .04 | .18 |
| Parietal lobe left white matter | 0.12 (0.03-0.21) | .004 | .03 |
| Parietal lobe right gray matter | 0.09 (0.03-0.14) | .003 | .03 |
| Parietal lobe right white matter | 0.08 (-0.01-0.17) | .06 | .20 |
| Subthalamic nucleus left | -0.08 (-0.25-0.08) | .03 | .16 |

| Regional volume | Standardized β coefficient (95% confidence intervals) | Raw *p* value | BH corrected *p* value |
|---|---|---|---|
| Subthalamic nucleus right | -0.05 (-0.36-0.25) | .21 | .45 |
| Superior temporal gyrus middle part left gray matter | 0.07 (-0.01-0.15) | .07 | .21 |
| Superior temporal gyrus middle part left white matter | 0.06 (-0.03-0.16) | .12 | .31 |
| Superior temporal gyrus middle part right gray matter | 0.10 (0.03-0.18) | .003 | .03 |
| Superior temporal gyrus middle part right white matter | 0.06 (-0.03-0.16) | .12 | .31 |
| Superior temporal gyrus posterior part left gray matter | 0.07 (-0.01-0.15) | .08 | .24 |
| Superior temporal gyrus posterior part left white matter | 0.04 (-0.06-0.14) | .34 | .57 |
| Superior temporal gyrus posterior part right gray matter | 0.04 (-0.04-0.12) | .33 | .55 |
| Superior temporal gyrus posterior part right white matter | 0.04 (-0.06-0.13) | .38 | .60 |
| Thalamus left high intensity part in T2 | -0.01 (-0.09-0.07) | .85 | .92 |
| Thalamus left low intensity part in T2 | 0.03 (-0.09-0.14) | .18 | .40 |
| Thalamus right high intensity part in T2 | -0.05 (-0.13-0.03) | .21 | .45 |
| Thalamus right low intensity part in T2 | 0.02 (-0.08-0.11) | .71 | .86 |
| *Interaction* | | | |
| Caudate nucleus right | 0.01 (-0.04-0.05) | .04 | .17 |
| Cerebrospinal fluid | -0.02 (-0.06-0.03) | <.001 | <.001 |
| Cingulate gyrus anterior part left white matter | 0.01 (-0.04-0.06) | .04 | .18 |
| Gyri parahippocampalis et ambiens anterior part left white matter | 0.01 (-0.04-0.07) | <.001 | <.001 |
| Gyri parahippocampalis et ambiens anterior part right white matter | 0.01 (-0.04-0.06) | .01 | .08 |
| Insula left white matter | 0.01 (-0.04-0.06) | .01 | .08 |
| Insula right white matter | 0.01 (-0.04-0.05) | .04 | .18 |
| Lateral ventricle left | -0.01 (-0.06-0.05) | <.001 | .01 |
| Lateral ventricle right | -0.01 (-0.06-0.04) | <.001 | .01 |
| Lentiform nucleus right | -0.01 (-0.06-0.03) | .05 | .18 |
| Medial and inferior temporal gyri anterior part right gray matter | 0.01 (-0.02-0.05) | .05 | .18 |
| Occipital lobe left gray matter | -0.02 (-0.05-0.02) | .04 | .18 |
| Occipital lobe right gray matter | -0.02 (-0.06-0.02) | .02 | .12 |
| Subthalamic nucleus left | -0.01 (-0.03-0.01) | .004 | .03 |
| Subthalamic nucleus right | -0.01 (-0.05-0.03) | .03 | .16 |
| *Paternal education* | | | |
| Amygdala left | 0.01 (-0.06-0.08) | .78 | .91 |
| Amygdala right | -0.01 (-0.07-0.06) | .87 | .94 |
| Anterior temporal lobe lateral part left gray matter | -0.02 (-0.08-0.04) | .52 | .73 |
| Anterior temporal lobe lateral part left white matter | 0.04 (-0.03-0.11) | .24 | .46 |
| Anterior temporal lobe lateral part right gray matter | 0.02 (-0.04-0.08) | .57 | .76 |
| Anterior temporal lobe lateral part right white matter | 0.05 (-0.03-0.12) | .15 | .35 |
| Anterior temporal lobe medial part left gray matter | 0.005 (-0.06-0.07) | .88 | .94 |
| Anterior temporal lobe medial part left white matter | 0.02 (-0.06-0.09) | .59 | .76 |
| Anterior temporal lobe medial part right gray matter | 0.04 (-0.02-0.11) | .17 | .39 |

| Regional volume | Standardized β coefficient (95% confidence intervals) | Raw *p* value | BH corrected *p* value |
|---|---|---|---|
| Anterior temporal lobe medial part right white matter | 0.07 (-0.01-0.14) | .04 | .18 |
| Brainstem | -0.02 (-0.08-0.04) | .52 | .73 |
| Caudate nucleus left | -0.01 (-0.07-0.05) | .67 | .83 |
| Caudate nucleus right | -0.03 (-0.22-0.17) | .36 | .58 |
| Cerebellum left | 0.001 (-0.05-0.05) | .97 | .99 |
| Cerebellum right | -0.01 (-0.06-0.04) | .78 | .91 |
| Cerebrospinal fluid | 0.04 (-0.15-0.23) | .09 | .25 |
| Cingulate gyrus anterior part left gray matter | 0.02 (-0.05-0.09) | .49 | .70 |
| Cingulate gyrus anterior part left white matter | 0.03 (-0.17-0.24) | .20 | .43 |
| Cingulate gyrus anterior part right gray matter | 0.04 (-0.03-0.10) | .23 | .45 |
| Cingulate gyrus anterior part right white matter | 0.06 (-0.01-0.13) | .05 | .18 |
| Cingulate gyrus posterior part left gray matter | -0.004 (-0.07-0.06) | .91 | .95 |
| Cingulate gyrus posterior part left white matter | 0.03 (-0.03-0.09) | .31 | .53 |
| Cingulate gyrus posterior part right gray matter | 0.01 (-0.05-0.08) | .70 | .86 |
| Cingulate gyrus posterior part right white matter | 0.03 (-0.03-0.10) | .27 | .50 |
| Corpus callosum | 0.04 (-0.03-0.10) | .22 | .45 |
| Frontal lobe left gray matter | 0.01 (-0.05-0.06) | .83 | .92 |
| Frontal lobe left white matter | 0.03 (-0.03-0.09) | .26 | .49 |
| Frontal lobe right gray matter | -0.01 (-0.06-0.05) | .83 | .92 |
| Frontal lobe right white matter | 0.05 (-0.01-0.11) | .08 | .23 |
| Gyri parahippocampalis et ambiens anterior part left gray matter | -0.03 (-0.11-0.04) | .30 | .51 |
| Gyri parahippocampalis et ambiens anterior part left white matter | -0.06 (-0.27-0.15) | .05 | .18 |
| Gyri parahippocampalis et ambiens anterior part right gray matter | -0.05 (-0.12-0.02) | .10 | .28 |
| Gyri parahippocampalis et ambiens anterior part right white matter | -0.01 (-0.21-0.20) | .78 | .91 |
| Gyri parahippocampalis et ambiens posterior part left gray matter | -0.001 (-0.07-0.07) | .98 | .99 |
| Gyri parahippocampalis et ambiens posterior part left white matter | 0.02 (-0.06-0.09) | .53 | .73 |
| Gyri parahippocampalis et ambiens posterior part right gray matter | -0.001 (-0.07-0.07) | .96 | .99 |
| Gyri parahippocampalis et ambiens posterior part right white matter | -0.01 (-0.08-0.06) | .82 | .92 |
| Hippocampus left | -0.01 (-0.07-0.06) | .85 | .92 |
| Hippocampus right | -0.003 (-0.08-0.07) | .93 | .97 |
| Insula left gray matter | -0.05 (-0.10-0.01) | .08 | .23 |
| Insula left white matter | 0.02 (-0.17-0.20) | .56 | .75 |
| Insula right gray matter | -0.03 (-0.08-0.03) | .28 | .50 |
| Insula right white matter | 0.02 (-0.17-0.20) | .58 | .76 |
| Lateral occipitotemporal gyrus gyrus fusiformis anterior part left gray matter | 0.01 (-0.06-0.07) | .80 | .91 |
| Lateral occipitotemporal gyrus gyrus fusiformis anterior part left white matter | 0.04 (-0.03-0.11) | .21 | .45 |
| Lateral occipitotemporal gyrus gyrus fusiformis anterior part right gray matter | -0.001 (-0.06-0.06) | .97 | .99 |

| Regional volume | Standardized β coefficient (95% confidence intervals) | Raw *p* value | BH corrected *p* value |
|---|---|---|---|
| Lateral occipitotemporal gyrus gyrus fusiformis anterior part right white matter | -0.002 (-0.07-0.07) | .95 | .99 |
| Lateral occipitotemporal gyrus gyrus fusiformis posterior part left gray matter | -0.03 (-0.10-0.04) | .35 | .57 |
| Lateral occipitotemporal gyrus gyrus fusiformis posterior part left white matter | 0.02 (-0.04-0.09) | .43 | .65 |
| Lateral occipitotemporal gyrus gyrus fusiformis posterior part right gray matter | -0.01 (-0.07-0.06) | .80 | .91 |
| Lateral occipitotemporal gyrus gyrus fusiformis posterior part right white matter | -0.02 (-0.09-0.05) | .49 | .70 |
| Lateral ventricle left | 0.04 (-0.17-0.25) | .06 | .20 |
| Lateral ventricle right | 0.02 (-0.18-0.22) | .27 | .49 |
| Lentiform nucleus left | -0.04 (-0.10-0.02) | .17 | .39 |
| Lentiform nucleus right | 0.04 (-0.12-0.20) | .29 | .51 |
| Medial and inferior temporal gyri anterior part left gray matter | 0.03 (-0.02-0.08) | .27 | .49 |
| Medial and inferior temporal gyri anterior part left white matter | 0.04 (-0.02-0.11) | .15 | .35 |
| Medial and inferior temporal gyri anterior part right gray matter | -0.01 (-0.15-0.13) | .78 | .91 |
| Medial and inferior temporal gyri anterior part right white matter | 0.03 (-0.04-0.10) | .40 | .62 |
| Medial and inferior temporal gyri posterior part left gray matter | 0.01 (-0.04-0.05) | .81 | .92 |
| Medial and inferior temporal gyri posterior part left white matter | -0.02 (-0.09-0.05) | .54 | .73 |
| Medial and inferior temporal gyri posterior part right gray matter | 0.01 (-0.04-0.05) | .79 | .91 |
| Medial and inferior temporal gyri posterior part right white matter | -0.001 (-0.07-0.07) | .96 | .99 |
| Occipital lobe left gray matter | 0.07 (-0.08-0.21) | .09 | .24 |
| Occipital lobe left white matter | 0.01 (-0.06-0.08) | .67 | .83 |
| Occipital lobe right gray matter | 0.07 (-0.09-0.22) | .07 | .21 |
| Occipital lobe right white matter | -0.01 (-0.08-0.06) | .69 | .84 |
| Parietal lobe left gray matter | 0.02 (-0.02-0.06) | .44 | .65 |
| Parietal lobe left white matter | 0.04 (-0.03-0.10) | .22 | .45 |
| Parietal lobe right gray matter | 0.01 (-0.03-0.05) | .72 | .86 |
| Parietal lobe right white matter | 0.03 (-0.03-0.10) | .30 | .51 |
| Subthalamic nucleus left | 0.0002 (-0.06-0.06) | .995 | .995 |
| Subthalamic nucleus right | -0.02 (-0.18-0.13) | .44 | .65 |
| Superior temporal gyrus middle part left gray matter | -0.001 (-0.06-0.06) | .98 | .99 |
| Superior temporal gyrus middle part left white matter | 0.03 (-0.04-0.10) | .28 | .50 |
| Superior temporal gyrus middle part right gray matter | 0.01 (-0.05-0.06) | .83 | .92 |
| Superior temporal gyrus middle part right white matter | 0.04 (-0.03-0.11) | .18 | .41 |
| Superior temporal gyrus posterior part left gray matter | -0.004 (-0.06-0.05) | .88 | .94 |
| Superior temporal gyrus posterior part left white matter | 0.05 (-0.02-0.12) | .07 | .21 |
| Superior temporal gyrus posterior part right gray matter | 0.01 (-0.05-0.07) | .66 | .83 |
| Superior temporal gyrus posterior part right white matter | 0.03 (-0.04-0.10) | .34 | .57 |
| Thalamus left high intensity part in T2 | -0.01 (-0.07-0.04) | .59 | .76 |
| Thalamus left low intensity part in T2 | -0.003 (-0.08-0.08) | .85 | .92 |

| Regional volume | Standardized β coefficient (95% confidence intervals) | Raw *p* value | BH corrected *p* value |
|---|---|---|---|
| Thalamus right high intensity part in T2 | -0.02 (-0.08-0.03) | .43 | .65 |
| Thalamus right low intensity part in T2 | -0.07 (-0.14- -0.01) | .02 | .12 |

Fully adjusted ridge regression model, including gestation at birth, paternal occupation, gestation at MRI, the interaction term (where significant), birth weight z-score, birth head circumference z-score, sex, smoking in pregnancy, and breast milk at discharge. Corrected for false discovery rate.
BH = Benjamini-Hochberg correction.

**eTable 10: Regional brain volumes – relationship with gestational age, subjective socioeconomic status, and interaction effect (fully adjusted ridge regression model)**

| Regional volume | Standardized β coefficient (95% confidence intervals) | Raw *p* value | BH corrected *p* value |
|---|---|---|---|
| *Gestation* | | | |
| Amygdala left | 0.06 (-0.05-0.16) | .21 | .46 |
| Amygdala right | 0.002 (-0.10-0.10) | .96 | .97 |
| Anterior temporal lobe lateral part left gray matter | 0.14 (0.06-0.23) | <.001 | .01 |
| Anterior temporal lobe lateral part left white matter | 0.09 (-0.02-0.19) | .05 | .18 |
| Anterior temporal lobe lateral part right gray matter | 0.06 (-0.03-0.15) | .19 | .45 |
| Anterior temporal lobe lateral part right white matter | 0.09 (-0.02-0.19) | .04 | .17 |
| Anterior temporal lobe medial part left gray matter | 0.06 (-0.04-0.15) | .21 | .46 |
| Anterior temporal lobe medial part left white matter | 0.02 (-0.10-0.13) | .67 | .81 |
| Anterior temporal lobe medial part right gray matter | 0.04 (-0.06-0.14) | .40 | .67 |
| Anterior temporal lobe medial part right white matter | 0.05 (-0.06-0.16) | .27 | .55 |
| Brainstem | -0.03 (-0.12-0.06) | .51 | .75 |
| Caudate nucleus left | 0.16 (0.07-0.26) | <.001 | .003 |
| Caudate nucleus right | 0.08 (-0.37-0.53) | .02 | .10 |
| Cerebellum left | -0.01 (-0.08-0.07) | .88 | .95 |
| Cerebellum right | -0.03 (-0.10-0.05) | .48 | .72 |
| Cerebrospinal fluid | -0.19 (-0.61-0.23) | <.001 | .003 |
| Cingulate gyrus anterior part left gray matter | -0.01 (-0.11-0.09) | .82 | .91 |
| Cingulate gyrus anterior part left white matter | 0.06 (-0.05-0.17) | .12 | .33 |
| Cingulate gyrus anterior part right gray matter | -0.07 (-0.17-0.04) | .13 | .36 |
| Cingulate gyrus anterior part right white matter | 0.03 (-0.08-0.14) | .42 | .67 |
| Cingulate gyrus posterior part left gray matter | -0.06 (-0.15-0.03) | .18 | .43 |
| Cingulate gyrus posterior part left white matter | 0.05 (-0.04-0.15) | .20 | .46 |
| Cingulate gyrus posterior part right gray matter | -0.10 (-0.19-0.002) | .03 | .14 |
| Cingulate gyrus posterior part right white matter | 0.07 (-0.03-0.16) | .12 | .33 |
| Corpus callosum | 0.09 (-0.01-0.19) | .03 | .13 |
| Frontal lobe left gray matter | 0.02 (-0.06-0.10) | .55 | .75 |
| Frontal lobe left white matter | 0.11 (-0.29-0.51) | .01 | .0499 |
| Frontal lobe right gray matter | 0.02 (-0.06-0.10) | .57 | .77 |
| Frontal lobe right white matter | 0.09 (-0.33-0.50) | .02 | .10 |
| Gyri parahippocampalis et ambiens anterior part left gray matter | 0.07 (-0.04-0.18) | .09 | .28 |
| Gyri parahippocampalis et ambiens anterior part left white matter | 0.09 (-0.39-0.57) | .01 | .0499 |
| Gyri parahippocampalis et ambiens anterior part right gray matter | 0.06 (-0.05-0.16) | .21 | .46 |
| Gyri parahippocampalis et ambiens anterior part right white matter | 0.09 (-0.01-0.19) | .05 | .17 |
| Gyri parahippocampalis et ambiens posterior part left gray matter | 0.12 (0.02-0.22) | .01 | .06 |
| Gyri parahippocampalis et ambiens posterior part left white matter | 0.06 (-0.05-0.17) | .16 | .40 |

| **Regional volume** | **Standardized β coefficient (95% confidence intervals)** | **Raw *p* value** | **BH corrected *p* value** |
|---|---|---|---|
| Gyri parahippocampalis et ambiens posterior part right gray matter | -0.003 (-0.11-0.10) | .95 | .97 |
| Gyri parahippocampalis et ambiens posterior part right white matter | 0.09 (-0.38-0.57) | .004 | .046 |
| Hippocampus left | -0.04 (-0.48-0.40) | .33 | .59 |
| Hippocampus right | -0.03 (-0.13-0.08) | .52 | .75 |
| Insula left gray matter | 0.04 (-0.05-0.12) | .36 | .62 |
| Insula left white matter | 0.18 (0.09-0.27) | <.001 | <.001 |
| Insula right gray matter | 0.10 (-0.27-0.47) | .11 | .33 |
| Insula right white matter | 0.10 (-0.34-0.54) | .01 | .07 |
| Lateral occipitotemporal gyrus gyrus fusiformis anterior part left gray matter | 0.14 (-0.31-0.58) | <.001 | .01 |
| Lateral occipitotemporal gyrus gyrus fusiformis anterior part left white matter | -0.08 (-0.18-0.02) | .08 | .27 |
| Lateral occipitotemporal gyrus gyrus fusiformis anterior part right gray matter | 0.15 (0.06-0.25) | <.001 | .005 |
| Lateral occipitotemporal gyrus gyrus fusiformis anterior part right white matter | -0.03 (-0.14-0.08) | .53 | .75 |
| Lateral occipitotemporal gyrus gyrus fusiformis posterior part left gray matter | 0.09 (-0.004-0.19) | .03 | .14 |
| Lateral occipitotemporal gyrus gyrus fusiformis posterior part left white matter | 0.002 (-0.44-0.44) | .96 | .97 |
| Lateral occipitotemporal gyrus gyrus fusiformis posterior part right gray matter | 0.05 (-0.04-0.15) | .22 | .48 |
| Lateral occipitotemporal gyrus gyrus fusiformis posterior part right white matter | -0.01 (-0.12-0.10) | .80 | .89 |
| Lateral ventricle left | -0.09 (-0.59-0.42) | <.001 | .005 |
| Lateral ventricle right | -0.07 (-0.55-0.40) | <.001 | .005 |
| Lentiform nucleus left | -0.10 (-0.19- -0.01) | .02 | .11 |
| Lentiform nucleus right | 0.06 (-0.32-0.44) | .38 | .64 |
| Medial and inferior temporal gyri anterior part left gray matter | 0.24 (0.16-0.31) | <.001 | <.001 |
| Medial and inferior temporal gyri anterior part left white matter | 0.04 (-0.06-0.13) | .41 | .67 |
| Medial and inferior temporal gyri anterior part right gray matter | 0.09 (-0.24-0.41) | .17 | .41 |
| Medial and inferior temporal gyri anterior part right white matter | 0.02 (-0.08-0.13) | .62 | .79 |
| Medial and inferior temporal gyri posterior part left gray matter | 0.12 (0.04-0.19) | .001 | .01 |
| Medial and inferior temporal gyri posterior part left white matter | -0.07 (-0.18-0.03) | .11 | .33 |
| Medial and inferior temporal gyri posterior part right gray matter | 0.13 (0.06-0.20) | <.001 | .005 |
| Medial and inferior temporal gyri posterior part right white matter | -0.03 (-0.13-0.08) | .54 | .75 |
| Occipital lobe left gray matter | -0.05 (-0.12-0.02) | .19 | .45 |
| Occipital lobe left white matter | -0.08 (-0.18-0.03) | .10 | .32 |
| Occipital lobe right gray matter | -0.02 (-0.10-0.05) | .52 | .75 |
| Occipital lobe right white matter | -0.07 (-0.18-0.03) | .11 | .33 |
| Parietal lobe left gray matter | 0.06 (0.004-0.12) | .03 | .14 |
| Parietal lobe left white matter | 0.11 (0.02-0.21) | .01 | .06 |
| Parietal lobe right gray matter | 0.08 (0.02-0.15) | .01 | .0499 |

| Regional volume | Standardized β coefficient (95% confidence intervals) | Raw *p* value | BH corrected *p* value |
|---|---|---|---|
| Parietal lobe right white matter | 0.07 (-0.03-0.17) | .10 | .31 |
| Subthalamic nucleus left | -0.18 (-0.28- -0.07) | <.001 | .005 |
| Subthalamic nucleus right | -0.12 (-0.22- -0.02) | .003 | .03 |
| Superior temporal gyrus middle part left gray matter | 0.08 (-0.003-0.17) | .04 | .17 |
| Superior temporal gyrus middle part left white matter | 0.06 (-0.05-0.16) | .16 | .41 |
| Superior temporal gyrus middle part right gray matter | 0.10 (0.02-0.18) | .01 | .06 |
| Superior temporal gyrus middle part right white matter | 0.06 (-0.04-0.17) | .12 | .34 |
| Superior temporal gyrus posterior part left gray matter | 0.07 (-0.02-0.16) | .10 | .31 |
| Superior temporal gyrus posterior part left white matter | 0.03 (-0.08-0.14) | .41 | .67 |
| Superior temporal gyrus posterior part right gray matter | 0.04 (-0.04-0.13) | .28 | .56 |
| Superior temporal gyrus posterior part right white matter | 0.05 (-0.05-0.16) | .21 | .46 |
| Thalamus left high intensity part in T2 | -0.03 (-0.12-0.05) | .41 | .67 |
| Thalamus left low intensity part in T2 | 0.02 (-0.11-0.14) | .28 | .56 |
| Thalamus right high intensity part in T2 | -0.06 (-0.15-0.03) | .17 | .41 |
| Thalamus right low intensity part in T2 | 0.01 (-0.10-0.12) | .77 | .87 |
| *Interaction* | | | |
| Caudate nucleus right | 0.001 (-0.005-0.01) | .01 | .06 |
| Cerebrospinal fluid | -0.003 (-0.01-0.003) | <.001 | <.001 |
| Frontal lobe left white matter | 0.001 (-0.004-0.01) | .01 | .08 |
| Frontal lobe right white matter | 0.001 (-0.004-0.01) | .03 | .13 |
| Gyri parahippocampalis et ambiens anterior part left white matter | 0.001 (-0.01-0.01) | .05 | .17 |
| Gyri parahippocampalis et ambiens posterior part right white matter | 0.001 (-0.01-0.01) | .05 | .17 |
| Hippocampus left | -0.001 (-0.01-0.005) | .04 | .15 |
| Insula right gray matter | -0.002 (-0.01-0.002) | .01 | .053 |
| Insula right white matter | 0.001 (-0.005-0.01) | .04 | .15 |
| Lateral occipitotemporal gyrus gyrus fusiformis anterior part left gray matter | 0.001 (-0.004-0.01) | .01 | .08 |
| Lateral occipitotemporal gyrus gyrus fusiformis posterior part left white matter | -0.001 (-0.01-0.004) | .01 | .06 |
| Lateral ventricle left | -0.001 (-0.01-0.01) | .001 | .02 |
| Lateral ventricle right | -0.001 (-0.01-0.01) | <.001 | .01 |
| Lentiform nucleus right | -0.003 (-0.01-0.002) | .001 | .02 |
| Medial and inferior temporal gyri anterior part right gray matter | 0.002 (-0.002-0.01) | .03 | .13 |
| *Subjective socioeconomic status* | | | |
| Amygdala left | -0.001 (-0.01-0.01) | .87 | .94 |
| Amygdala right | 0.0003 (-0.01-0.01) | .93 | .97 |
| Anterior temporal lobe lateral part left gray matter | -0.01 (-0.01-0.002) | .13 | .35 |
| Anterior temporal lobe lateral part left white matter | 0.002 (-0.01-0.01) | .65 | .80 |
| Anterior temporal lobe lateral part right gray matter | 0.002 (-0.01-0.01) | .64 | .79 |
| Anterior temporal lobe lateral part right white matter | 0.004 (-0.01-0.01) | .34 | .60 |

| **Regional volume** | **Standardized β coefficient (95% confidence intervals)** | **Raw *p* value** | **BH corrected *p* value** |
|---|---|---|---|
| Anterior temporal lobe medial part left gray matter | -0.01 (-0.02- -0.002) | .01 | .06 |
| Anterior temporal lobe medial part left white matter | -0.001 (-0.01-0.01) | .79 | .88 |
| Anterior temporal lobe medial part right gray matter | -0.002 (-0.01-0.01) | .66 | .80 |
| Anterior temporal lobe medial part right white matter | -0.00005 (-0.01-0.01) | .99 | .99 |
| Brainstem | 0.001 (-0.01-0.01) | .77 | .87 |
| Caudate nucleus left | 0.003 (-0.01-0.01) | .47 | .72 |
| Caudate nucleus right | 0.003 (-0.02-0.02) | .47 | .72 |
| Cerebellum left | 0.003 (-0.004-0.01) | .35 | .61 |
| Cerebellum right | 0.002 (-0.005-0.01) | .61 | .78 |
| Cerebrospinal fluid | 0.01 (-0.01-0.03) | .02 | .12 |
| Cingulate gyrus anterior part left gray matter | -0.0004 (-0.01-0.01) | .92 | .97 |
| Cingulate gyrus anterior part left white matter | 0.001 (-0.01-0.01) | .83 | .91 |
| Cingulate gyrus anterior part right gray matter | -0.002 (-0.01-0.01) | .63 | .79 |
| Cingulate gyrus anterior part right white matter | 0.001 (-0.01-0.01) | .75 | .86 |
| Cingulate gyrus posterior part left gray matter | -0.002 (-0.01-0.01) | .67 | .81 |
| Cingulate gyrus posterior part left white matter | 0.005 (-0.003-0.01) | .21 | .46 |
| Cingulate gyrus posterior part right gray matter | -0.0003 (-0.01-0.01) | .93 | .97 |
| Cingulate gyrus posterior part right white matter | -0.001 (-0.01-0.01) | .71 | .84 |
| Corpus callosum | 0.002 (-0.01-0.01) | .54 | .75 |
| Frontal lobe left gray matter | -0.003 (-0.01-0.004) | .37 | .63 |
| Frontal lobe left white matter | 0.003 (-0.01-0.02) | .34 | .60 |
| Frontal lobe right gray matter | -0.004 (-0.01-0.003) | .26 | .54 |
| Frontal lobe right white matter | 0.004 (-0.02-0.02) | .31 | .59 |
| Gyri parahippocampalis et ambiens anterior part left gray matter | -0.001 (-0.01-0.01) | .72 | .84 |
| Gyri parahippocampalis et ambiens anterior part left white matter | 0.002 (-0.02-0.02) | .68 | .82 |
| Gyri parahippocampalis et ambiens anterior part right gray matter | -0.002 (-0.01-0.01) | .53 | .75 |
| Gyri parahippocampalis et ambiens anterior part right white matter | -0.004 (-0.01-0.01) | .36 | .62 |
| Gyri parahippocampalis et ambiens posterior part left gray matter | 0.001 (-0.01-0.01) | .77 | .87 |
| Gyri parahippocampalis et ambiens posterior part left white matter | 0.003 (-0.01-0.01) | .44 | .69 |
| Gyri parahippocampalis et ambiens posterior part right gray matter | 0.01 (-0.001-0.02) | .04 | .15 |
| Gyri parahippocampalis et ambiens posterior part right white matter | 0.004 (-0.02-0.03) | .26 | .54 |
| Hippocampus left | 0.005 (-0.01-0.02) | .21 | .46 |
| Hippocampus right | -0.001 (-0.01-0.01) | .72 | .84 |
| Insula left gray matter | -0.005 (-0.01-0.003) | .19 | .46 |
| Insula left white matter | 0.01 (-0.002-0.01) | .08 | .28 |
| Insula right gray matter | 0.01 (-0.01-0.03) | .01 | .08 |
| Insula right white matter | 0.004 (-0.02-0.02) | .30 | .59 |
| Lateral occipitotemporal gyrus gyrus fusiformis anterior part left gray matter | -0.01 (-0.03-0.01) | .07 | .24 |

| Regional volume | Standardized β coefficient (95% confidence intervals) | Raw *p* value | BH corrected *p* value |
|---|---|---|---|
| Lateral occipitotemporal gyrus gyrus fusiformis anterior part left white matter | -0.004 (-0.01-0.005) | .32 | .59 |
| Lateral occipitotemporal gyrus gyrus fusiformis anterior part right gray matter | -0.001 (-0.01-0.01) | .84 | .91 |
| Lateral occipitotemporal gyrus gyrus fusiformis anterior part right white matter | -0.003 (-0.01-0.01) | .46 | .71 |
| Lateral occipitotemporal gyrus gyrus fusiformis posterior part left gray matter | 0.003 (-0.01-0.01) | .44 | .70 |
| Lateral occipitotemporal gyrus gyrus fusiformis posterior part left white matter | 0.01 (-0.01-0.03) | .01 | .06 |
| Lateral occipitotemporal gyrus gyrus fusiformis posterior part right gray matter | 0.002 (-0.01-0.01) | .58 | .78 |
| Lateral occipitotemporal gyrus gyrus fusiformis posterior part right white matter | 0.00005 (-0.01-0.01) | .99 | .99 |
| Lateral ventricle left | 0.001 (-0.02-0.02) | .60 | .78 |
| Lateral ventricle right | 0.001 (-0.02-0.02) | .52 | .75 |
| Lentiform nucleus left | -0.002 (-0.01-0.01) | .65 | .80 |
| Lentiform nucleus right | 0.01 (-0.01-0.03) | .05 | .17 |
| Medial and inferior temporal gyri anterior part left gray matter | -0.004 (-0.01-0.003) | .25 | .52 |
| Medial and inferior temporal gyri anterior part left white matter | 0.002 (-0.01-0.01) | .53 | .75 |
| Medial and inferior temporal gyri anterior part right gray matter | -0.004 (-0.02-0.01) | .33 | .59 |
| Medial and inferior temporal gyri anterior part right white matter | 0.0003 (-0.01-0.01) | .94 | .97 |
| Medial and inferior temporal gyri posterior part left gray matter | -0.0002 (-0.01-0.01) | .94 | .97 |
| Medial and inferior temporal gyri posterior part left white matter | 0.01 (-0.003-0.02) | .11 | .33 |
| Medial and inferior temporal gyri posterior part right gray matter | 0.002 (-0.004-0.01) | .55 | .75 |
| Medial and inferior temporal gyri posterior part right white matter | 0.003 (-0.01-0.01) | .47 | .72 |
| Occipital lobe left gray matter | 0.0005 (-0.01-0.01) | .88 | .94 |
| Occipital lobe left white matter | 0.004 (-0.01-0.01) | .31 | .59 |
| Occipital lobe right gray matter | 0.002 (-0.005-0.01) | .61 | .79 |
| Occipital lobe right white matter | 0.003 (-0.01-0.01) | .42 | .67 |
| Parietal lobe left gray matter | -0.001 (-0.01-0.004) | .60 | .78 |
| Parietal lobe left white matter | 0.003 (-0.01-0.01) | .51 | .75 |
| Parietal lobe right gray matter | -0.001 (-0.01-0.004) | .60 | .78 |
| Parietal lobe right white matter | 0.001 (-0.01-0.01) | .72 | .84 |
| Subthalamic nucleus left | -0.002 (-0.01-0.01) | .59 | .78 |
| Subthalamic nucleus right | -0.01 (-0.01-0.004) | .16 | .40 |
| Superior temporal gyrus middle part left gray matter | -0.0005 (-0.01-0.01) | .90 | .95 |
| Superior temporal gyrus middle part left white matter | -0.001 (-0.01-0.01) | .76 | .87 |
| Superior temporal gyrus middle part right gray matter | 0.0004 (-0.01-0.01) | .91 | .96 |
| Superior temporal gyrus middle part right white matter | -0.001 (-0.01-0.01) | .69 | .82 |
| Superior temporal gyrus posterior part left gray matter | -0.01 (-0.01-0.002) | .14 | .37 |
| Superior temporal gyrus posterior part left white matter | 0.003 (-0.01-0.01) | .32 | .59 |
| Superior temporal gyrus posterior part right gray matter | -0.004 (-0.01-0.004) | .31 | .59 |
| Superior temporal gyrus posterior part right white matter | -0.004 (-0.01-0.01) | .33 | .59 |

| **Regional volume** | **Standardized β coefficient (95% confidence intervals)** | **Raw *p* value** | **BH corrected *p* value** |
|---|---|---|---|
| Thalamus left high intensity part in T2 | 0.002 (-0.005-0.01) | .50 | .74 |
| Thalamus left low intensity part in T2 | 0.002 (-0.01-0.01) | .29 | .57 |
| Thalamus right high intensity part in T2 | 0.002 (-0.01-0.01) | .63 | .79 |
| Thalamus right low intensity part in T2 | 0.001 (-0.01-0.01) | .83 | .91 |

Fully adjusted ridge regression model, including gestation at birth, subjective socioeconomic status (as World Health Organisation Quality of Life, environment domain), gestation at MRI, the interaction term (where significant), birth weight z-score, birth head circumference z-score, sex, smoking in pregnancy, and breast milk at discharge. Corrected for false discovery rate.
BH = Benjamini-Hochberg correction.

**eTable 11: Cortical measures – relationship with gestational age and maternal final educational qualification (fully adjusted ridge regression model)**

| Cortical measure | Standardized β coefficient (95% confidence intervals) | Raw *p* value | BH corrected *p* value |
|---|---|---|---|
| *Gestation* | | | |
| Mean cortical curvature | -0.0001 (-0.10-0.10) | .63 | .84 |
| Mean cortical surface area | 0.01 (-0.11-0.12) | .48 | .84 |
| Mean cortical thickness | -0.002 (-0.10-0.09) | .61 | .84 |
| Mean gyrification index | 0.005 (-0.11-0.12) | .48 | .84 |
| Mean sulcal depth | 0.003 (-0.12-0.12) | .92 | .92 |
| *Maternal education* | | | |
| Mean cortical curvature | 0.0001 (-0.09-0.09) | .52 | .84 |
| Mean cortical surface area | -0.01 (-0.11-0.09) | .22 | .84 |
| Mean cortical thickness | 0.004 (-0.08-0.09) | .15 | .84 |
| Mean gyrification index | -0.002 (-0.10-0.10) | .77 | .85 |
| Mean sulcal depth | 0.01 (-0.11-0.13) | .67 | .84 |

Fully adjusted ridge regression model, including gestation at birth, maternal final educational qualification, gestation at MRI, the interaction term (where significant – none were, so not included), birth weight z-score, birth head circumference z-score, sex, smoking in pregnancy, and breast milk at discharge. Corrected for false discovery rate.
BH = Benjamini-Hochberg correction.

**eTable 12: Cortical measures – relationship with gestational age and paternal final educational qualification (fully adjusted ridge regression model)**

| Cortical measure | Standardized β coefficient (95% confidence intervals) | Raw *p* value | BH corrected *p* value |
|---|---|---|---|
| *Gestation* | | | |
| Mean cortical curvature | -0.01 (-0.13-0.10) | .67 | .74 |
| Mean cortical surface area | 0.01 (-0.12-0.13) | .63 | .74 |
| Mean cortical thickness | -0.01 (-0.12-0.10) | .38 | .74 |
| Mean gyrification index | 0.01 (-0.11-0.13) | .49 | .74 |
| Mean sulcal depth | 0.01 (-0.12-0.14) | .76 | .76 |
| *Paternal education* | | | |
| Mean cortical curvature | -0.04 (-0.14-0.06) | .18 | .74 |
| Mean cortical surface area | 0.01 (-0.09-0.11) | .60 | .74 |
| Mean cortical thickness | 0.01 (-0.08-0.10) | .48 | .74 |
| Mean gyrification index | 0.01 (-0.09-0.11) | .41 | .74 |
| Mean sulcal depth | -0.02 (-0.13-0.09) | .59 | .74 |

Fully adjusted ridge regression model, including gestation at birth, paternal final educational qualification, gestation at MRI, the interaction term (where significant – none were, so not included), birth weight z-score, birth head circumference z-score, sex, smoking in pregnancy, and breast milk at discharge. Corrected for false discovery rate.
BH = Benjamini-Hochberg correction.

#### eTable 13: Cortical measures – relationship with gestational age, maternal occupation, and interaction effect (fully adjusted ridge regression model)

| **Cortical measure** | **Standardized β coefficient (95% confidence intervals)** | **Raw *p* value** | **BH corrected *p* value** |
|---|---|---|---|
| *Gestation* | | | |
| Mean cortical curvature | -0.004 (-0.11-0.10) | .41 | .74 |
| Mean cortical surface area | 0.01 (-0.10-0.12) | .46 | .74 |
| Mean cortical thickness | -0.001 (-0.11-0.11) | .51 | .74 |
| Mean gyrification index | 0.01 (-0.10-0.12) | .54 | .74 |
| Mean sulcal depth | 0.005 (-0.11-0.12) | .88 | .88 |
| *Interaction* | | | |
| Mean gyrification index | 0.01 (-0.06-0.07) | .02 | .27 |
| *Maternal occupation* | | | |
| Mean cortical curvature | 0.001 (-0.06-0.06) | .81 | .88 |
| Mean cortical surface area | -0.01 (-0.07-0.06) | .44 | .74 |
| Mean cortical thickness | 0.0004 (-0.06-0.06) | .75 | .88 |
| Mean gyrification index | -0.01 (-0.08-0.06) | .40 | .74 |
| Mean sulcal depth | 0.03 (-0.05-0.10) | .20 | .74 |

Fully adjusted ridge regression model, including gestation at birth, maternal occupation, gestation at MRI, the interaction term (where significant), birth weight z-score, birth head circumference z-score, sex, smoking in pregnancy, and breast milk at discharge. Corrected for false discovery rate.
BH = Benjamini-Hochberg correction.

#### eTable 14: Cortical measures – relationship with gestational age and paternal occupation (fully adjusted ridge regression model)

| Cortical measure | Standardized β coefficient (95% confidence intervals) | Raw *p* value | BH corrected *p* value |
|---|---|---|---|
| *Gestation* | | | |
| Mean cortical curvature | -0.02 (-0.13-0.09) | .56 | .92 |
| Mean cortical surface area | 0.01 (-0.10-0.12) | .67 | .92 |
| Mean cortical thickness | -0.001 (-0.11-0.11) | .61 | .92 |
| Mean gyrification index | 0.01 (-0.10-0.12) | .53 | .92 |
| Mean sulcal depth | 0.01 (-0.11-0.13) | .79 | .92 |
| *Paternal occupation* | | | |
| Mean cortical curvature | -0.002 (-0.07-0.07) | .92 | .92 |
| Mean cortical surface area | 0.01 (-0.06-0.09) | .45 | .92 |
| Mean cortical thickness | -0.001 (-0.06-0.06) | .66 | .92 |
| Mean gyrification index | 0.01 (-0.06-0.09) | .46 | .92 |
| Mean sulcal depth | -0.01 (-0.09-0.08) | .83 | .92 |

Fully adjusted ridge regression model, including gestation at birth, paternal occupation, gestation at MRI, the interaction term (where significant – none were, so not included), birth weight z-score, birth head circumference z-score, sex, smoking in pregnancy, and breast milk at discharge. Corrected for false discovery rate.
BH = Benjamini-Hochberg correction.

#### eTable 15: Cortical measures – relationship with gestational age, subjective socioeconomic status, and interaction effect (fully adjusted ridge regression model)

| Cortical measure | Standardized β coefficient (95% confidence intervals) | Raw *p* value | BH corrected *p* value |
|---|---|---|---|
| *Gestation* | | | |
| Mean cortical curvature | 0.004 (-0.08-0.09) | .92 | .92 |
| Mean cortical surface area | 0.09 (0.003-0.17) | .03 | .11 |
| Mean cortical thickness | -0.01 (-0.12-0.09) | .75 | .92 |
| Mean gyrification index | 0.16 (0.07-0.26) | <.001 | .002 |
| Mean sulcal depth | 0.04 (-0.40-0.48) | .10 | .27 |
| *Interaction* | | | |
| Mean sulcal depth | 0.0005 (-0.01-0.01) | .02 | .11 |
| *Subjective socioeconomic status* | | | |
| Mean cortical curvature | -0.002 (-0.01-0.01) | .60 | .92 |
| Mean cortical surface area | 0.001 (-0.01-0.01) | .81 | .92 |
| Mean cortical thickness | -0.002 (-0.01-0.005) | .67 | .92 |
| Mean gyrification index | -0.001 (-0.01-0.01) | .86 | .92 |
| Mean sulcal depth | 0.004 (-0.01-0.02) | .15 | .33 |

Fully adjusted ridge regression model, including gestation at birth, subjective socioeconomic status (as World Health Organisation Quality of Life, environment domain), gestation at MRI, the interaction term (where significant), birth weight z-score, birth head circumference z-score, sex, smoking in pregnancy, and breast milk at discharge. Corrected for false discovery rate.
BH = Benjamini-Hochberg correction.
